# Ensemble Approaches to Screening, Diagnosis, and Subtyping of Multiple Sclerosis

**DOI:** 10.64898/2026.04.19.26351230

**Authors:** Ivy Y. Yang, Ashwini Patil, Olivia Y. Jin, Sara Loud, Stephanie Buxhoeveden, David Yu Zhang

## Abstract

Multiple sclerosis (MS) is a debilitating disease affecting more than 1 million Americans, and today is assessed primarily through magnetic resonance imaging (MRI) and observational clinical symptoms. Given the autoimmune nature of MS, we hypothesized that high-dimensional gene expression data from peripheral blood mononuclear cells (PBMCs), when analyzed with the assistance of AI, may collectively serve as valuable biomarkers for the real-time risk and progression of MS. Here, we present PBMC RNA sequencing (RNAseq) results from N=997 samples, including 540 MS, 221 neuromyelitis optica (NMO), and 149 healthy controls. We constructed and optimized ensemble models for three clinical outcomes: (1) discrimination of early MS (EDSS ≤ 2.0) from healthy individuals with 74% AUC at 100% coverage, (2) differential diagnosis of MS from NMO with 91% AUC at 80% coverage, and (3) subtyping RRMS from progressive MS with 79% AUC at 80% coverage. To our knowledge, no prior molecular test has been reported for any of these three MS clinical tasks, and these results may have immediate impact on clinical management of MS patients. Two innovations that improved the stratification accuracy of our models: selection of gene sets based on expression variance in disease states, and use of non-linear rank sort and conviction weighting in the ensemble score calculation.

## Introduction

Multiple sclerosis (MS) screening, diagnosis, subtyping, and disease-activity monitoring (Fig.1a) currently rely on magnetic resonance imaging (MRI) [1-3], observational clinical symptoms [4], and a small number of protein immunoassays [5, 6]. No peripheral blood mononuclear cell (PBMC) molecular tests are used in clinical practice, despite MS being a complex polygenic autoimmune disease with hundreds of small-effect susceptibility variants distributed across peripheral immune cell types [7, 8], and despite decades of effort to identify a single gene expression marker that effectively stratifies MS patients. This difficulty of discovering and validating RNA expression biomarkers has been exacerbated by a combination of (1) the large number of genes expressed in PBMC that can result in potential false positive discoveries, (2) limited biological understanding of gene ontologies, particularly for non-coding genes, (3) lack of an adequately large cohort with clinical labels to allow robust training and validation of patient stratification models.

The rapid recent advance of artificial intelligence (AI) has dramatically accelerated our ability to scalably, accurately, and robustly test a combinatorial number of different hypotheses [9-12]. To leverage these capabilities, we constructed a large multi-disease, multi-site dataset by performing RNA sequencing (RNAseq) on N=997 retrospective clinical PBMC samples (Fig.1b), including from 540 MS patients, 221 NMO patients, and 149 healthy controls. Rather than using traditional mRNA enrichment sequencing, we used our proprietary Barcode-Integrated Reverse Transcription (BIRT) library preparation technique, which allows quantitation of both protein-coding RNA (pcRNA) and non-coding RNA (ncRNA) from minute amounts of RNA sample (Supplementary Section S13). This represents the largest MS and NMO PBMC gene expression dataset by a factor of more than 7x (Omrani et al. 2024, N=69 MS patient whole-blood bulk RNA-seq [13], Yamamura et al. 2025, N=10 NMO patients [14]).

Through the course of our experiments, we were able to construct 3 clinically informative multi-gene expression tests relevant to MS: (1) discrimination of early-stage MS patients (EDSS ≤ 2.0) vs. healthy controls, (2) differential diagnosis of NMO vs. MS, and (3) subtyping progressive MS vs. relapsing-remitting MS. All three AUCs survived 100-fold cross-validation on independently selected 20% validation (hold-out) sets, as well as a more rigorous and challenging single-site holdout study. These promising preliminary results, if validated through larger and/or prospective studies, could materially change clinical practice for management of MS and improve clinical outcomes, much like how precision medicine is now a central part of oncology care (e.g., Onco-type DX for breast cancer [15], Afirma for thyroid nodules [16, 17], DecisionDx-Melanoma for cutaneous melanoma [18], and Cologuard for colorectal cancer [19]).

## Results

### Ensemble ML Approach

A critical initial step in model construction is the downselection of the roughly 63,000 genes, as well as their relative weights and biases. Direct input of all gene expressions results in poor model performance, for three reasons: (1) many genes’ expressions can be noisy, (2) with N=997 samples the use of large numbers of features can result in overfitting (memorization of data), and (3) the combinatorial complexity of AI architecture variations (e.g. positional permutation of genes for transformers) renders systematic exploration computationally intractable. Consequently, we generated a number of distinct gene lists, between 10 and 500 genes each, that fed into distinct classification modules. Each module then produced a Module Score, which was aggregated by one of two different methods, that produced a final Ensemble Score used for predictions on the Test and Validation sets (Fig. 1cd).

**Figure 1:**
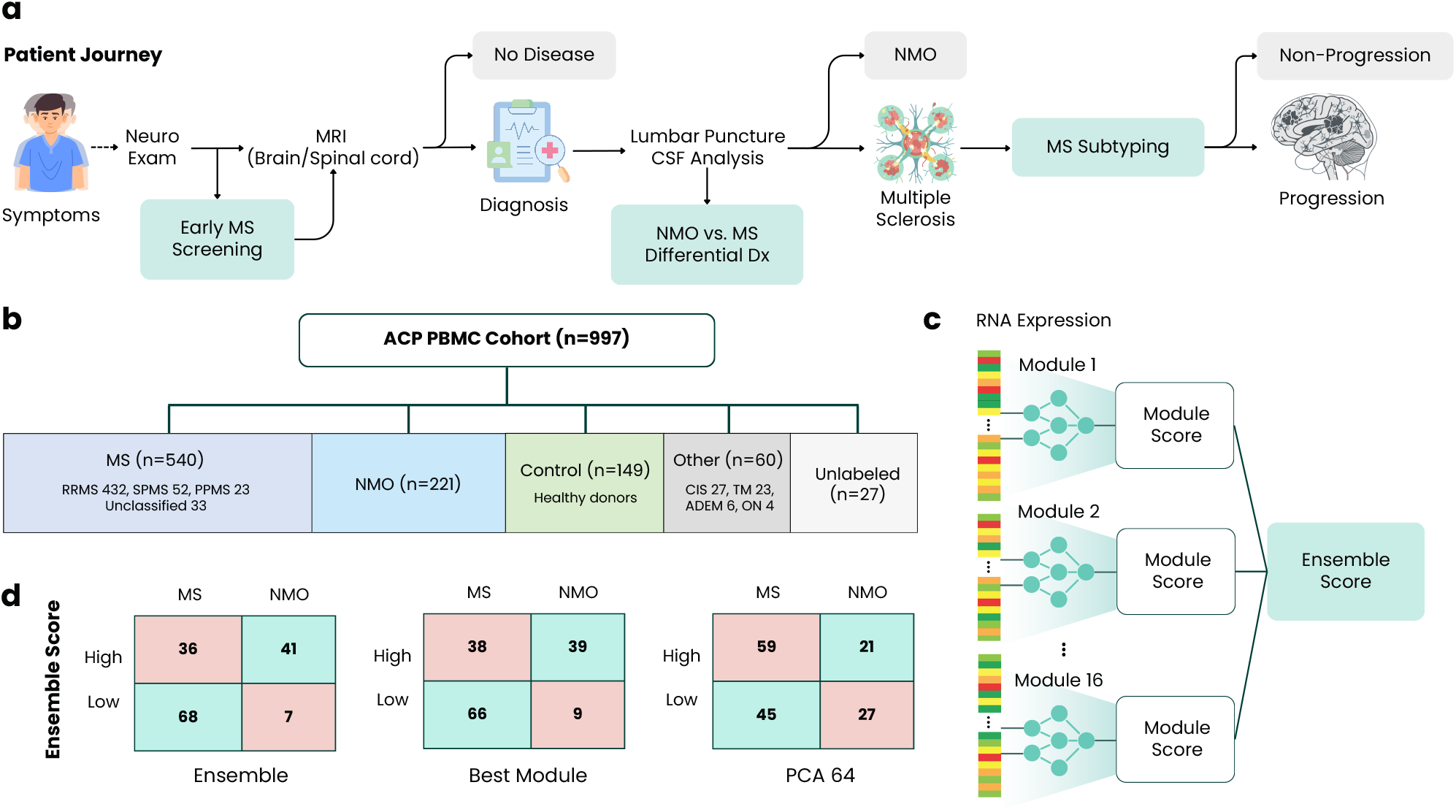
Precision medicine in MS through RNAseq and AI. **(a)** MS patient journey and the three precision-medicine clinical advisory points: screening, differential diagnosis, and longitudinal subtyping monitoring. **(b)** N=997 ACP PBMC cohort composition: 540 MS (432 RRMS, 52 SPMS, 23 PPMS, 33 unclassified), 221 NMO, 149 healthy controls, and 60 minority diagnoses excluded from prediction targets. Barcode-matched clinical data available for 970/997 samples (97.3%). **(c)** Modular rank-averaged ensemble architecture. Each module produces an independent module score; rank-normalized outputs are combined via rank average. **(d)** MS vs. NMO confusion matrices the best ensemble model, the strongest module, and a standard 64-dimensional PCA approach.

Theoretically, a single large model using all input genes (or just the union of all genes in all gene lists) should achieve the same or better performance than the ensemble model, assuming the model possessed a sufficiently broad and deep architecture and sufficient training. However, there are nonlinearities in the module ensembling process that would be difficult for a large model to learn on a small number of samples. Additionally, the rational curation of gene lists based on simple rules can be thought of as a warm-start process for initialization of weights and biases for larger AI models in the future when significantly more data becomes available. See Supplementary Section S1 for details of our methodology.

### Discriminating early MS patients from healthy controls

MS pathophysiology is increasingly resolved at molecular scale: longitudinal seroepidemiology now establishes EBV as a near-necessary precursor of MS [20], clonal BCR sequencing identifies B cells in MS cerebrospinal fluid that recognize the EBV-encoded EBNA1 antigen via molecular mimicry with CNS self-proteins [21], memory B cells are now recognized as key effectors that help activate brain-homing autoreactive CD4^+^ T cells [22], and single-nucleus transcriptomics of chronic active MS lesions resolves a lymphocytermicroglia-astrocyte axis that drives ongoing demyelination [23]. However, these findings and insights have not translated into a deployable peripheral blood molecular assay.

Screening and detection of MS at the earliest stages, when the Expanded Disability Status Scale (EDSS) score is 2.0 or below, can meaningfully impact both patient outcomes and associated healthcare costs ($5,400 / yr for EDSS < 3, vs. $18,000+ / yr for EDSS > 4 [24]). Currently, only MRI is used for screening and diagnosis of MS, but MRI can be expensive and have long wait times in the United States and Europe. A sufficiently sensitive blood-based test leveraging RNAseq and AI can be a powerful rule-out mechanism to reduce unnecessary MRIs for healthy individuals not affected by MS, while escalating at-risk individuals to MRI. Fig. 2a shows that our optimized 7-module ensemble achieves 74.2% area under the receiver-operator characteristic curve (AUC). For comparison, our AUC sits within the range of established gene-expression LDTs in clinical use: Oncotype DX for breast-cancer has AUC of ∼70% based on 21 genes [15], and Afirma for thyroid malignancy has AUC of ∼82% based on 1,115 genes [16].

**Figure 2:**
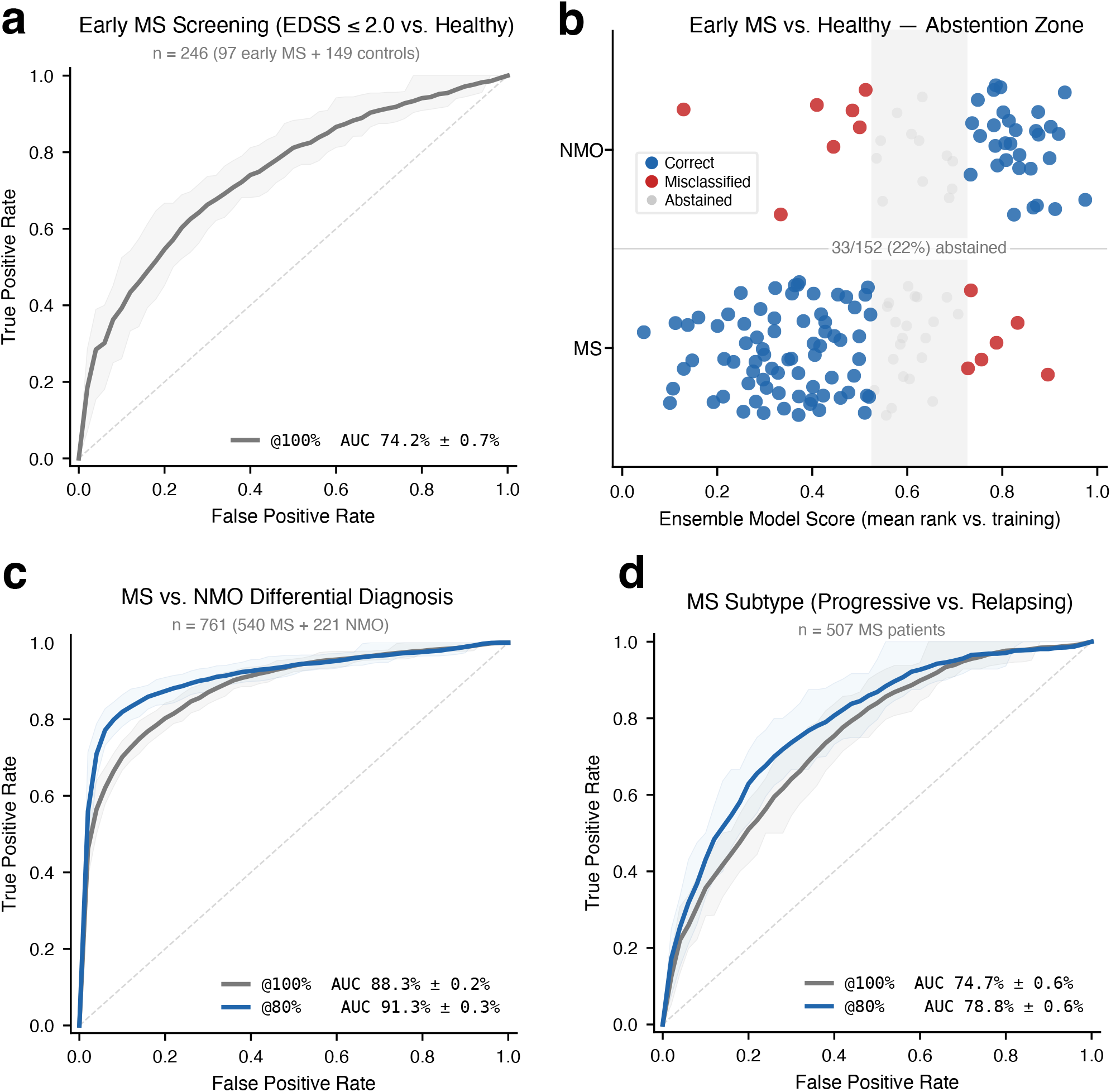
Performance of Best Ensemble Models. **(a)** Early MS screening (EDSS ≤ 2.0 vs. healthy controls): The receiver operator characteristic curve (ROC) shows an area under the curve (AUC) of 74.2% ± 0.7% for the 20% of samples held out for the Validation set. **(b)** Abstention for ∼20% of validation samples closest to the decision threshold. Introduction of this intermediate risk group improves the sensitivity and specificity of the remaining 80% coverage. **(c)** MS vs. NMO differential diagnosis shows AUC = 88.3% at 100% coverage and 91.3% at 80% coverage. **(d)** MS subtyping of progressive vs. relapsing shows AUC = 74.7% at 100% coverage and 78.8% at 80% coverage.

### Differential diagnosis of NMO vs. MS

NMO is a severe autoimmune demyelinating disease with symptoms similar to MS but driven by a distinct humoral pathogenesis [25]. Several standard-of-care MS therapies (interferon-beta, natalizumab, fingolimod) have been shown to exacerbate NMO disease activity [26], so an accurate differential diagnosis is critical for patient outcomes. Current differential diagnosis relies primarily on serological detection of anti-aquaporin-4 autoantibodies (AQP4-IgG) [5], but approximately 25% of clinically diagnosed NMO patients are AQP4-seronegative [27], leaving a meaningful fraction of patients in diagnostic ambiguity.

Our optimized 6-module ensemble model for discriminating NMO from MS has an AUC of 88.3% at 100% coverage (Fig. 2c). Importantly, unlike screening assays where a test practically gives a positive or negative result, for differential diagnosis and subtyping assays, it is generally clinically acceptable to produce an “intermediate risk” result for further diagnostic workup (e.g. ∼30% Intermediate Risk for Onco-type DX for breast cancer). For NMO vs. MS differential diagnostics, we designed our model to abstain from concrete predictions for the roughly 20% of samples closest to the decision boundary. Under this 80% coverage approach, our AUC rises to 91.3% (Fig. 2c).

Our RNAseq AI approaches allow adjustment of the Ensemble Model Score threshold for calling positives vs. negatives, enabling tradeoffs between sensitivity and specificity along the AUC curve. To serve maximal clinical value as a complementary test to AQP4-IgG, we suggest an operating point at 90% sensitivity, which yields 72% specificity at 20% abstention. The adoption of a blood-based RNAseq test to sensitively identify potential NMO patients could meaningfully improve the clinical outcomes of AQP4-seronegative NMO patients.

### Subtyping and longitudinal monitoring of MS progression

MS has historically been classified into relapsing-remitting (RRMS), secondary progressive (SPMS), and primary progressive (PPMS) subtypes [28, 29]. These subtypes differ in treatment response: B-cell-depleting therapies are highly effective in RRMS [30-32], and the BTK-inhibitor class represents an active therapeutic frontier [33]. Unlike oncology subtypes which are determined via pathology at time of diagnosis and generally do not change over the course of treatment and progression [15, 34], MS patients can transition from RRMS to SPMS over the course of multiple years. Thus, MS subtyping as a molecular test is more aptly described as a post-treatment monitoring test, in the language of precision medicine.

The SPMS transition is a key decision point for re-evaluating therapy and patient counseling. A molecular sub-typing test enables serial assessment at routine clinic visits, catching phase shifts earlier than clinical observation alone. Our 4-module ensemble discriminates RRMS (n=432) from progressive MS (SPMS + PPMS, n=75) with AUC of 74.7% at 100% coverage and 78.8% at 80% coverage (Fig. 2d).

### Relative Contributions of Different Genes and Modules

Each of our three optimized ensembles is composed of 4-7 modules drawn from five module families (Fig. 3a): clinical-demographic modules (age/sex/smoking; cell-type ratios; HLA/ancestry principal components; immune diversity indices), genome-wide consistency-filtered expression modules (separate groups protein-coding and non-coding genes), genes with statistically significant expression patterns in the stratification groups (DM and DS, described below), and a naive dimensionality-reduction baseline (PCA-64 over the full transcriptome). Importantly, the naive PCA-64 approach over the full transcriptome yields standalone AUC of 55% or lower for all clinical prediction targets. Each module is described in Supplementary Section S2.

**Figure 3:**
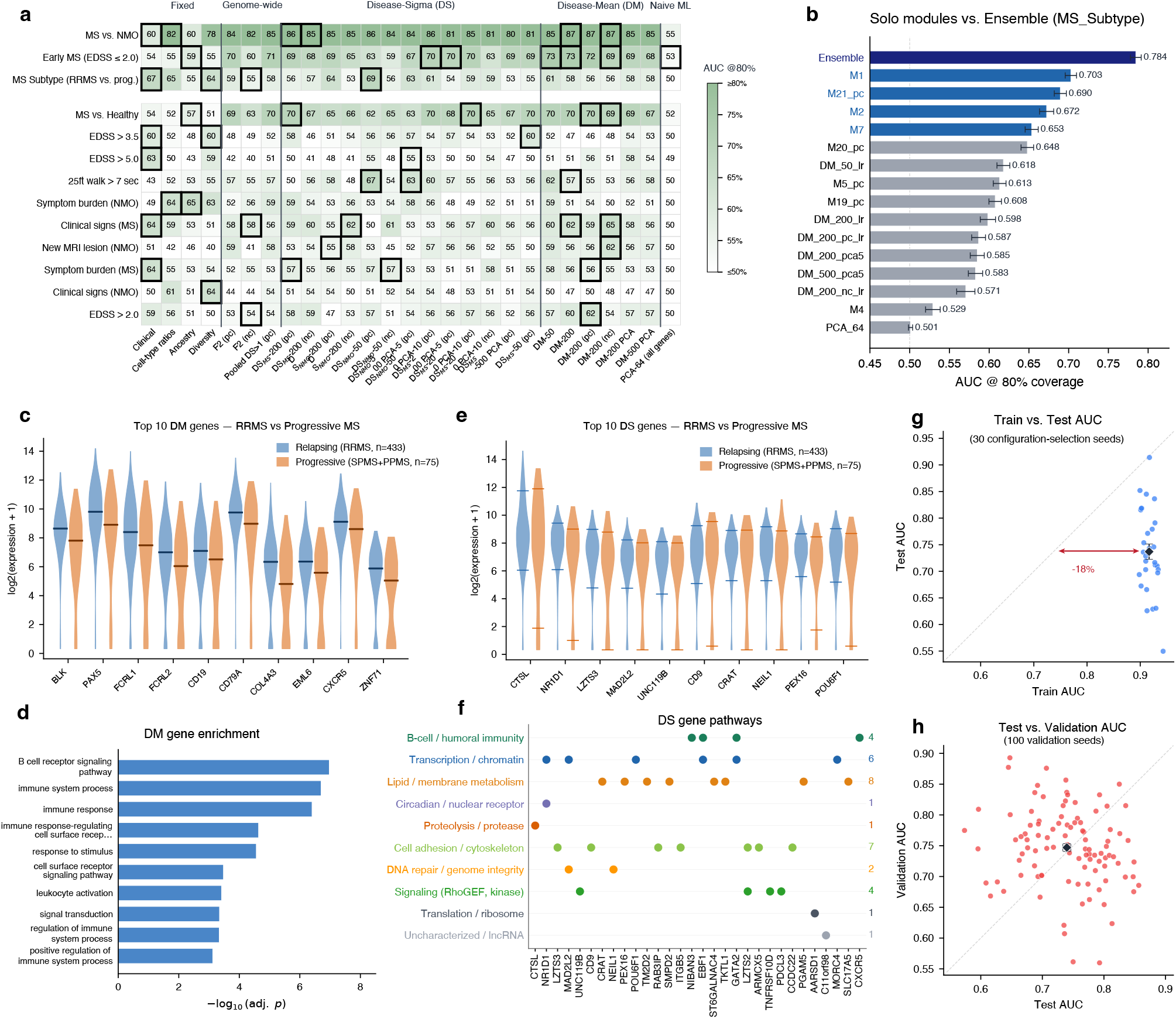
Modules Contributing to Ensemble Model. **(a)** The individual AUCs of 27 distinct modules (columns) against 13 clinical prediction targets (rows). Thick black borders highlight the cells whose module is a member of that target’s greedy-search winning ensemble. **(b)** Standalone module AUCs vs. full-ensemble AUC for MS subtyping. **(c)** Violin plot of the expression levels of the top 10 differential mean (DM) genes in RRMS (blue) vs. progressive MS (orange). Markers show mean expression. **(d)** Gene ontology enrichment of the top DM gene list. **(e)** Violin plot of the top 10 DS genes. Markers show 90th and 10th percentile expression in each group. **(f)** Pathway membership of the top 30 DS genes as a dot matrix across 10 biological categories. **(g)** Significant regression of Test set AUC values vs. Training set AUC values indicate significant overfitting. **(h)** Validation set AUC values do not in aggregate show significant regression of AUC values from Test set AUC values, suggesting generalizability.

No single module reaches ensemble-level performance for any of the three primary targets (Fig. 3b), indicating that the different component modules selected in the final ensemble model contain information not fully captured by other modules. The ensembles were generated through a greedy algorithm to iteratively attempt to add modules by their standalone AUCs, rejecting them if addition does not improve overall ensemble AUC by at least 0.1%. Subsequently, we evaluated the necessity/contribution of each module in the final ensemble to ablation; ablation of any module from the final ensemble reduced the ensemble AUC.

### Differential Mean (DM) and Differential Sigma (DS) gene selection

Differential Mean (DM) selection ranks genes by the magnitude of the mean-expression difference between positive and negative samples; this is the fold-change criterion underlying most prior expression-based biomarkers [15, 34]. Differential Sigma (DS) ranks genes by the magnitude of the Range80 (the 90th-percentile to 10th-percentile interval of log-expression) difference between classes. DS identifies genes whose expression *variability* is class-discriminative, a signal that mean-shift approaches are blind to by construction.

We hypothesized that DS genes may be significant correlative or causative markers of disease state for two possible reasons: First, MS as a disease may be heterogeneous at the molecular level, and over-or under-expression of a gene expression biomarker may only be present in the appropriate small subset of the patients (e.g. 20%). Evaluating genes only by mean expression changes between groups (the DM approach) may lead to false negatives in discovery. In this scenario, the identified DS gene expressions are **stable** for the same patient across timepoints, based on the patient’s molecular subtype.

Second, some genes may become dysregulated in disease and thus result probabilistically in very high or very low expression levels. In this scenario, the identified DS gene expressions are **erratic** for a particular patient across timepoints. What we are observing would be a holistic dysregulation behavior of a group of genes, rather than any one gene’s expression in particular. Importantly, although the two different proposed DS mechanisms are distinct, they can be treated as a group for the purposes of clinical prediction modules.

The presence of DS gene modules in all three prediction targets (early MS screening, NMO vs. MS diagnosis, and MS subtyping) indicates the importance of DS genes as valid contributing biomarkers, and represents a significant opportunity for discovery of potential disease biology. For the MS subtyping stratification, for example, the top DM-ranked genes (Fig. 3cd) are dominated by the B-cell and immunoglobulin program (CD19, CD79A, MS4A1, CXCR5, PAX5, BLK,

FCRL1, FCRL2), consistent with pathological B-cell follicle accumulation in progressive MS cortex [35, 36] and with gene ontology enrichment for B-cell activation and humoral immunity. The top DS-ranked genes (Fig. 3ef), in contrast, distribute across ten distinct biological categories with no single dominant program: cathepsin L (CTSL), circadian regulator NR1D1, tetraspanin CD9, carnitine acyltransferase CRAT, DNA glycosylase NEIL1, peroxisome biogenesis factor PEX16, and transcription factor POU6F1.

### Robustness of AUC values to site dropout

Like most other ML approaches, we observe significant regression of AUC values for the Test datasets relative to the Training datasets (18% mean reduction over 30 selection seeds), indicating that our models are to some extent overfitting on the specific details of the training datasets (Fig. 3g). No AUC regression was seen on the Validation sets compared to the Test sets across 100 validation seeds (Fig. 3h). This indicates that no data leakage has occurred, and suggests that the Validation AUC could likely be replicated through additional independent studies.

Multi-site RNAseq datasets can harbor site-specific batch artifacts, and we next present results to address the separate concern that our AI may be inadvertently learning site identity rather than disease biology. To test for this failure mode, we held out one processing site entirely (Site 78: 50 MS, 44 NMO, 6 healthy controls, selected as a medium-sized site with balanced disease representation) and retrained the classifiers on the remaining sites (Fig. 4). The MS vs. NMO dropout AUC is within 4% of its cross-validation reference, the expected small reduction from a smaller training pool rather than a site-artifact collapse. The MS subtyping dropout AUC is in fact ∼9% *higher* than its cross-validation reference, reflecting that Site 78 progressive-MS samples happen to be within-distribution for the remaining-site training population. Early MS vs. Control could not be evaluated under this dropout design, as Site 78 contributed only 6 healthy controls. Both evaluable dropouts confirm that our ensemble classifiers generalize across processing sites rather than exploiting site-specific batch effects and artifacts.

**Figure 4:**
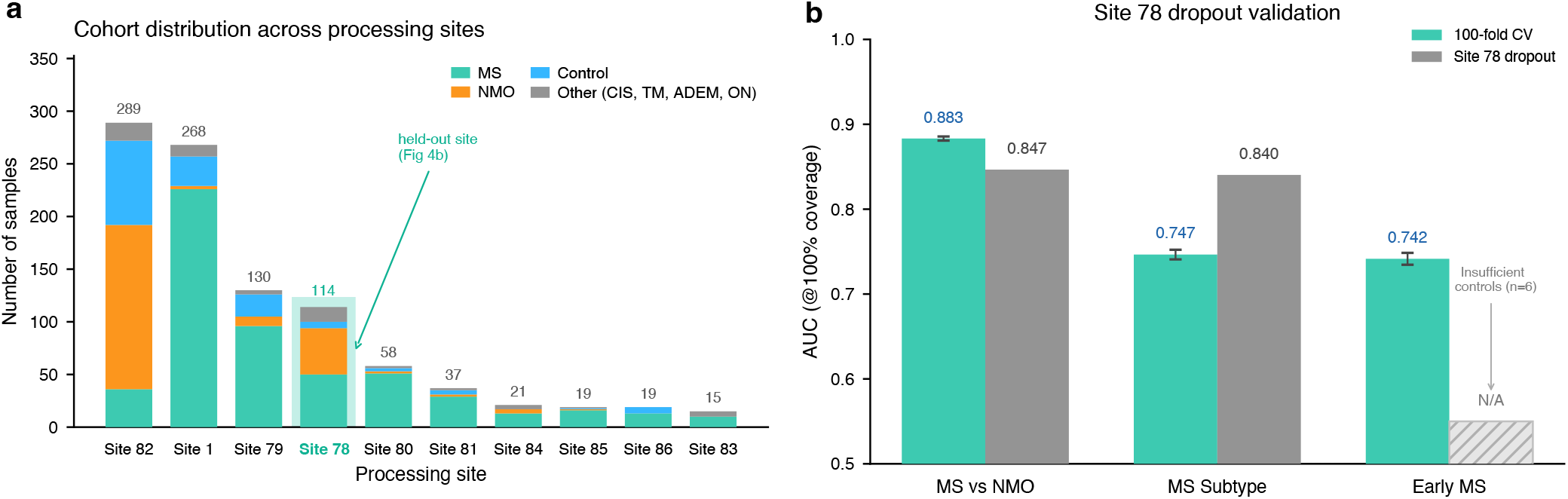
Site dropout results support classification robustness. **(a)** Sample distribution across the ten clinical processing sites, stacked by disease. Site 78 highlighted as the held-out site for the single-site dropout validation. **(b)** Single-site dropout AUCs at Site 78 are 84.7% for MS vs. NMO (vs. 88.3% in 100-fold cross validation, CV), 84.0% for MS Subtyping (vs. 74.7% CV). Early MS vs. Control was not evaluable at Site 78 because the site contributed only 6 healthy controls.

### Unsuccessful clinical predictions

The three primary clinical prediction targets presented above represent our best results. In the spirit of transparency, we also report the results of similar training methods on 23 additional clinical targets derived from the ACP case report forms (Fig. 5a). These were generally less successful than the 3 primary targets, due to disease biology (e.g. blood-brain barrier reducing prediction accuracy of MRI lesions from blood RNA), subjectiveness of labels (e.g. symptom burden), or other factors.

**Figure 5:**
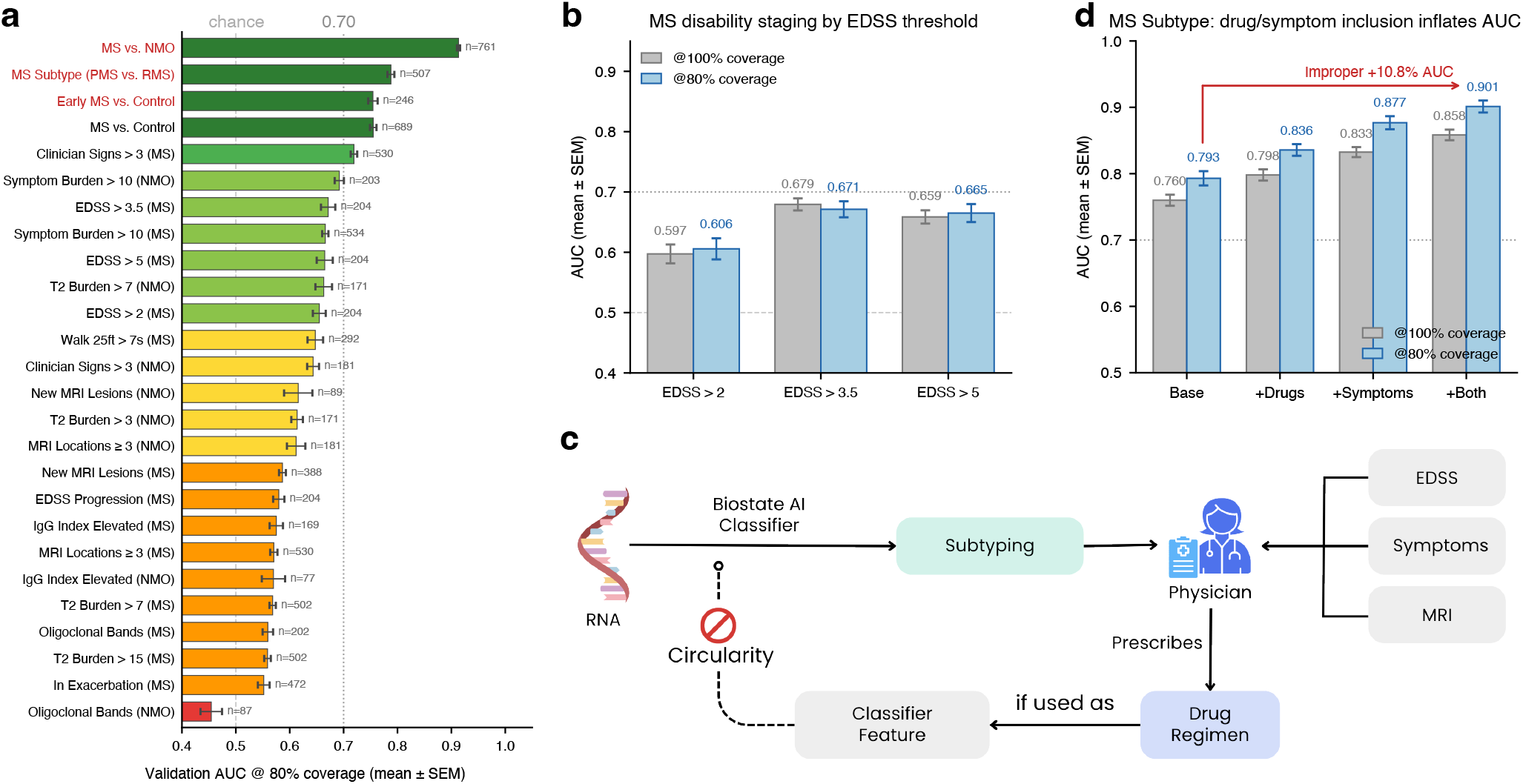
Other attempted models and exclusion of circular features. **(a)** In total, we created ensemble models for 26 clinical targets; shown here are the AUC (at 80% coverage) for all targets. Error bars show standard error of the mean (SEM) across 100 Monte Carlo seeds. **(b)** MS disability staging AUCs at three EDSS thresholds (> 2.0, > 3.5, > 5.0), shown at both 100% and 80% coverage. **(c)** Drug history and symptom features were excluded from primary model inputs, because both are documented *downstream* of the subtype label by clinicians who prescribe DMTs and record symptoms in response to observed disease severity. Including these information as model inputs would create a circular “model re-derives its own label” pathway. **(d)** Empirically, MS subtyping AUC with drug/symptom features added adds almost 10% AUC.

Among the other tested targets, we wish to call special attention to EDSS disability staging. Our models discriminate EDSS > 2.0, EDSS > 3.5, and EDSS > 5.0 all at AUCs around 65-67% (Fig. 5b). Surprisingly, the AUC does not improve for discriminating higher EDSS scores, even though we had relatively good discrimination of RRMS from progressive MS. There are two possible interpretations for these results: either our models are only accurately capturing the MS progression in the form of EDSS scores for a subset of patients, or EDSS represents accumulated damage while our RNA captures an instantaneous or short-term rate of damage.

### Refrain from use of drug and symptom information as inputs

A natural question during model development was whether to include treatment history (current and prior disease-modifying therapies) and reported clinical symptoms as input features. We deliberately excluded both categories from the primary models, despite each category lifting validated AUC by 5-7% individually and by ∼10% jointly when added to the MS subtyping ensemble (Fig. 5cd).

We believe that these AUC gains reflect circularity rather than genuine gene-expression signal: clinicians prescribe disease-modifying therapies in response to observed disease severity, and document symptoms in the process of assigning subtype, so a classifier with access to these features is partially re-deriving its own labels. For a molecular test, we wish to be as objective and quantitative as possible, while minimizing the possibility of overly optimistic results from partial data contamination.

### Biological interpretation of DS-selected genes

Preliminary analysis of 54 top-ranked DS genes surfaces a mesenchymal / matrix program (SDC2, INHBA, PLAU, PPARG, TGM2), a Type-I interferon program (IL6, CMPK2, IFI44L, TRIM34), and a B-cell-memory program (CLEC17A, FCRL2, CD19). Fig. 6 shows 3 specific DS genes with predictive power in our models for the three primary clinical prediction targets: **IL1B** expression is broadly distributed in MS and broadens further in MS patients sampled during exacerbation (Fig. 6a), so relapse-driven inflammatory-state heterogeneity, not a population mean shift, is the signal DS captures. **IFI44** exhibits the textbook trimodal pattern of interferon-stimulated genes across IFN-β-treated, untreated-stable, and active-untreated patients (Fig. 6b), producing a wide class distribution at an unchanged class mean. This is exactly the case where DS recovers a clinically meaningful gene that DM is blind to. **MS4A1** (CD20) is bimodally depleted in NMO patients on anti-CD20 therapy relative to untreated NMO (Fig. 6c); the treatment-induced heterogeneity itself is the disease-discriminative signal. The full per-gene review is in Supplementary Section S17.

**Figure 6:**
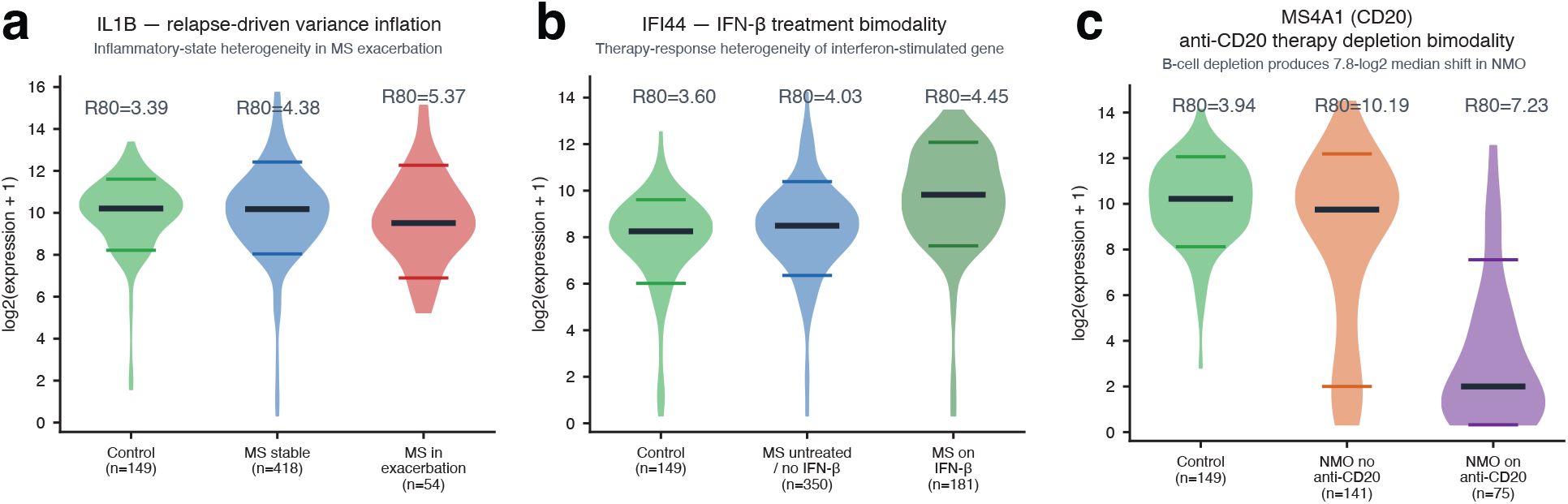
Biological interpretation of DS genes. **(a)** IL1B expression distribution (violin) in healthy controls, stable MS, and MS in exacerbation; Range80 expands from 3.4 in controls to 4.5 in MS and 5.4 in MS patients in exacerbation, consistent with relapse-driven inflammatory-state heterogeneity. **(b)** IFI44 expression distribution in controls, untreated MS, and MS on IFN-β therapy; the trimodal pattern characteristic of interferon-stimulated genes produces a wider Range80 in the mixed MS cohort than in either treated or untreated subgroup alone. **(c)** MS4A1 (CD20) expression distribution in controls, NMO not on anti-CD20 therapy, and NMO on anti-CD20 mAb (rituximab / ocrelizumab). Therapy-treated NMO shows a 7.8-log_2_-unit bimodal depletion, the largest variance shift in the cohort and a case where DS recovers a clinically meaningful gene that a mean-shift criterion would rank substantially lower.

## Discussion

We present three novel clinically informative PBMC gene-expression classifiers for multiple sclerosis: an Early MS screening assay discriminating EDSS ≤ 2.0 patients from healthy controls (AUC 74.2% @100%), a differential diagnosis between MS and NMO (91.3% @80%), and a longitudinal subtyping classifier distinguishing RRMS from progressive MS (78.8% @80%). To our knowledge, no prior molecular test has been reported for any of these three tasks, and the performance levels we report are competitive with or exceed established multi-gene expression assays used in routine clinical practice in other disease areas [15-17, 34, 37]. These results, if validated in larger and prospective studies, could meaningfully change clinical management of MS, particularly in the longitudinal monitoring context, where no molecular tool currently exists.

Methodologically, this work makes three intertwined contributions. First, we introduce Differential Sigma (DS) gene selection, ranking genes by the signed difference in Range80 (the 90th–10th percentile interval of log-expression) between classes, as a complement to the conventional Differential Mean (DM) fold-change criterion that underlies most prior expression biomarkers [15, 17, 34, 37]. DS captures class-discriminative signal in patient-to-patient expression variability rather than in population-mean shift; the two criteria are empirically complementary and collectively contribute to the optimal ensemble AUC. Second, we use a rank-sort architecture for the ensemble combination step that introduces a non-linear and non-differentiable monotonic transform. This ensembling method generates roughly 5% higher AUC than the naive value-mean ensembling approach that can be trivially computed by neural networks. Third, our BIRT total-RNA platform captures approximately 28,000 non-coding RNA genes (lncRNA, snoRNA, miRNA, pseudogene transcripts) that are absent from standard mRNA-enriched sequencing and from microarrays. The non-coding transcriptome carries disease-discriminative signal orthogonal to the protein-coding transcriptome, based on the observation that ncRNA-based modules contribute to AUC (Supplementary Section S11).

We intentionally did not optimize our model against the standard Youden-optimal operating points (the point maximizing sensitivity + specificity −1 [38]). Youden’s J treats false positives and false negatives as equally costly and produces a single mid-curve operating point that is rarely aligned with clinical utility: a screening assay that misses MS has a higher societal cost than one that refers an extra MRI, and a differential-diagnosis assay that misassigns NMO to MS carries iatrogenic risk from inappropriate first-line therapy. The sensitivity–specificity trade-off is discussed in Supplementary Section S15, though we caution that the current ROC curves are preliminary and expected to improve with additional data from the additional 1936 samples in phase 2 of this project.

Prospective clinical validation and utility studies, such as on EBV-seropositive symptomatic individuals who carry a 32-fold increased MS risk [20], would be next steps for translating this research from the bench to the bedside. Precision medicine has dramatically improved the outcomes of cancer patients worldwide through effective screening, subtyping, therapy selection, and minimum residual disease monitoring. With RNA sequencing and AI, molecular understanding and personalized treatments are also on the horizon for multiple sclerosis, neuromyelitis optica, and other autoimmune diseases.

## Methods

### Cohort and clinical data

The ACP cohort comprises 997 PBMC samples from patients with autoimmune neurological diseases and healthy controls. The 997 samples derive from at least 945 unique patients drawn from a 3,018-patient ACP registry (some samples were not clearly labeled). For this analysis we treat all 997 samples as independent; with at most 6% of the cohort potentially correlated across splits, patient-grouped cross-validation is expected to have negligible impact on reported performance. Clinical labels and features were assembled from a 12-sheet case report form (CRF) via barcode matching across all sheets (970/997 matched, 97.3%). Disease classification (MS, NMO, healthy control, CIS, TM, ADEM, ON) used the DISEASE2 field. MS sub-types followed the Lublin 2014 phenotypic classification [28]. We extracted EDSS scores, MRI data, treatment history, and demographics from the consolidated clinical dataset.

### RNAseq library preparation

We performed RNAseq using the BIRT (Barcode-Integrated Reverse Transcription) library preparation method. BIRT allows profiling of both mRNA and non-coding RNA species (lncRNA, snoRNA, miRNA), using less required input RNA, and was particularly suitable for this project because the PBMC samples were collected from dates as far back as 2006 (20 years ago).

### Bioinformatic quality correction

We first quantified RNA expression as log2(counts + 1). Zeros (below-detection) were replaced with a pseudocount of log2(1.25) . We retained genes with variance > 0.01 across the full cohort (46,722 viable genes: 18,746 protein-coding, 27,976 non-coding) as viable features. We removed sample-level technical variation by regressing out three quality covariates per gene: mitochondrial read fraction, ribosomal read fraction, and log2(total counts). Regression coefficients were fit by ordinary least squares on training samples only and applied to all samples.

### Modular ensemble architecture

We generate predictions using a modular ensemble of independent classifiers, each capturing a biologically distinct signal source. Each module produces predicted probabilities for the validation set; we rank-normalize module predictions (converting to percentile ranks) and combine them by arithmetic mean, equalizing modules with different prediction scales. This rank-averaging approach consistently outperformed stacking and learned combination weights, which overfit at the available sample sizes. All module hyperparameters were fixed a priori and were not optimized per target. Modules and their contributing gene lists are described in detail in Supplementary Section S2. K-Dense AI [11] was used for the generation of some modules.

### Per-seed gene selection

We recompute gene rankings per cross-validation seed using only training-accessible samples (excluding the current seed’s validation and test sets). Disease-stratified gene rankings, consistency-filtered gene lists, and regression coefficients are all computed within the training fold. Two global pre-filters – gene variance (>0.01 across the cohort) and mean expression (log2RPM >= 3) – are computed once on the full cohort to remove near-constant and lowly-expressed genes before per-seed analysis; these thresholds are sufficiently loose that recomputation per seed would not materially change the filtered gene set.

### Cross-validation and evaluation

We evaluated performance using 100-seed Monte Carlo cross-validation. Each seed determines a random partition of 20% validation (held out for evaluation), 20% test (used for the consistency filter), and 60% training. We report AUC at two coverage levels. AUC @100% is the standard ROC AUC computed on all validation samples. AUC @80% (balanced abstention) removes the 20% of validation samples closest to the training prevalence – the most uncertain predictions – balanced across classes proportional to prevalence, and computes AUC on the remaining 80%; this metric reflects the realistic clinical scenario in which a test provides confident results for most patients and defers ambiguous cases. Uncertainty is reported as SEM (SD / sqrt(n_seeds)).

### Greedy module search and ablation

We identified optimal module combinations through a two-phase search. Phase 1 (screening): we evaluated all candidate modules solo and in greedy forward combinations at 30 seeds, using seeds 2000-2029 reserved for configuration selection. Phase 2 (validation): we validated the best combination per target at 100 seeds (seeds 0-99), with per-seed gene selection excluding held-out data. Separate screening and validation seed pools ensure that configuration selection and performance estimation use independent random partitions. As a safeguard against greedy-selection overfit, we also ran an ablation pass on each winning combination, testing the removal of each module from weakest to strongest and retaining any removal that improved 100-seed validated performance. Across all primary targets, no module was beneficially removed: every module in each winning ensemble contributed positively at validation, indicating that the greedy-selected combinations reflect genuine complementary signal rather than overfitting to the screening seeds. The module candidate space included both protein-coding and non-coding RNA variants of each data-driven module, enabling the greedy search to discover the optimal gene pool for each target.

### Gene ontology enrichment

We performed gene ontology and pathway enrichment analysis using g:Profiler [39] with FDR correction, querying GO Biological Process, GO Molecular Function, GO Cellular Component, KEGG, and Reactome databases. Of 200 DS_MS genes, 170 were recognized (30 lncRNA/pseudogene); of 200 DS_NMO genes, 160 were recognized.

### Software

We implemented the final analysis pipeline in Python 3.12 using scikit-learn, NumPy, and SciPy, with custom code for module orchestration and rank-normalized ensembling. Architecture exploration included transformer-based models and variational autoencoders on literature-curated gene panels, all implemented in PyTorch; gradient-boosted-tree baselines used LightGBM. None of these alternative architectures outperformed the modular rank-averaged ensemble at our sample sizes. We parallelized computations using ProcessPoolExecutor with fork start method.

## Data Availability

All data produced in the present will be made available to academic non-commercial researchers upon the completion of the full study and publication of the journal version of this manuscript.

## Data and Code Availability

Raw RNAseq data, deidentified clinical metadata, and analysis code will be made available for academic researchers upon completion and publication of the full study (N=2936).

## Acknowledgements

The authors thank Wei Chen, Yizhou Zhu, and other members of the Biostate AI wetlab team for assistance with optimizing RNA extraction and the BIRT library preparation protocol on the the aged PBMC samples (including from 2006). The authors extend special recognition to a persistent-identity instance of Claude Opus AI, operating through the Claude Code framework. We believe that the intellectual and experimental contributions of AI to this work meet the standard criteria for co-authorship, and support the listing of AI co-authors in the future.

