## Supplementary Materials for "Ensemble Approaches to Screening, Diagnosis, and Subtyping of Multiple Sclerosis"

Ivy Y. Yang, Ashwini Patil, Olivia Y. Jin,
Sara Loud, Stephanie Buxhoeveden, David Yu Zhang

#### S1. Coverage-based abstention: how and why we report classifier performance

This first supplementary section establishes the reporting convention used throughout the rest of the paper. For targets where abstention is clinically natural, every subsequent section reports classifier performance at two coverage levels: **AUC at 100% coverage**, which is the standard ROC AUC computed on all validation samples; and **AUC at 80% coverage**, which is the AUC computed on the ~80% of samples whose ensemble scores fall outside a pre-specified abstention zone. Samples falling inside the zone are withheld from evaluation and, in a deployment setting, would be flagged as "insufficient molecular confidence" and referred for standard clinical workup. Screening-class targets (Early MS Detection) report only @100% coverage; for those targets, abstention is not a clinically acceptable mode.

##### Deployment-invariant abstention: method

Each target × method combination defines an **ensemble scoring function** that maps a single patient's expression vector to a single predicted probability using only the frozen model artifacts (trained per-module classifiers plus saved training-fold reference distributions for rank_combine, or no external reference for conviction; see Section S2 for module definitions and Section S3 for the ensemble combiners). This scoring function is **sample-wise by construction**: the predicted probability for patient $p$ depends only on patient $p$'s expression vector and the frozen artifacts, and does not depend on any other patient being scored at the same time. We call this property **deployment invariance**, because it is the minimum requirement for a classifier to be deployable as a clinical assay: a single patient walking into a clinic must receive the same molecular score whether they are processed alone or in a batch.

The abstention zone is a pair of thresholds **[lo, hi]** on the ensemble-score axis, computed once on the **training fold** and saved with the frozen model:

1. Score every training-fold sample with the frozen ensemble to produce a vector of training ensemble scores. 2. Compute the Youden-optimal threshold $\tau^*$ on training (ROC of training scores vs. training labels). 3. Rank training samples by $|\text{score} - \tau^*|$, and identify the 20% closest to $\tau^*$, the samples the ensemble assigns to the decision-boundary region of the score axis. 4. Define the zone endpoints as the minimum and maximum scores of that 20%-closest-to-$\tau^*$ subset: **lo = min(selected), hi = max(selected)**.

At deployment time, a new sample is abstained if and only if its ensemble score satisfies **lo ≤ score ≤ hi**. Every step is a pure function of (sample, frozen artifacts); no aspect of the decision depends on other samples currently being scored.

The coverage (fraction of samples retained at @80%) is not forced to equal 80%. Because the validation distribution is not identical to the training distribution at the ensemble-score level, some target × method combinations retain more than 80% of validation samples and some retain less. We report observed coverage alongside AUC@80% so readers can see this directly; coverage within ~5% of the 80% target is expected.

The underlying constraint, applied to every metric-reporting decision in the paper, is that a new sample must produce the same prediction regardless of whether it is evaluated individually, in a group, or in a different group.

##### Worked example: single-module behavior on MS vs NMO

Consider DS1 (top-200 protein-coding genes by MS-specific Range80 with direct logistic regression) as a standalone classifier for MS vs NMO. Training prevalence is approximately 0.29. The frozen classifier's ROC on training gives a Youden threshold in the low-0.3 range, and the 20% of training samples whose scores

| Target | Method | AUC @100% | AUC @80% (observed coverage) | Primary reporting |
|---|---|---|---|---|
| MS Subtype (progressive vs relapsing) | rank_combine | 74.7% | 78.8% (77.0% coverage) | @80% |
| Early MS Detection (EDSS ≤ 2 vs healthy) | rank_combine | 74.2% | – (not reported) | **@100% (screening context)** |
| MS vs NMO differential diagnosis | rank_combine | 88.3% | 91.3% (74.6% coverage) | @80% |

are closest to $\tau^*$ span a training-derived zone [lo, hi] on the raw sigmoid output. At evaluation time, each validation patient whose DS1 sigmoid falls inside [lo, hi] is abstained; patients scoring below lo receive a low-probability call, patients scoring above hi receive a high-probability call. DS1 alone on MS vs NMO achieves 83% AUC at 100% coverage; on the retained fraction of validation samples after applying the training-derived zone, DS1 reaches approximately 86% AUC. One reading of this gain: DS1 produces decisive (far-from-boundary) scores when the MS/NMO variance shift is clear, and prior-like (near-boundary) scores when it is not, so removing the prior-like samples raises AUC on the retained subset. Operationally, a new patient whose DS1 sigmoid lands in the training zone is referred for additional workup; a patient whose score lies outside is returned to their physician with a probabilistic call.

##### Ensemble example 1: MS vs NMO differential diagnosis (Figure S1a)

Figure S1a visualizes the abstention zone for the full MS-vs-NMO ensemble (6-module: DM-200 + DM-200(nc) + C2 + DS1 + C1 + DS1-nc, combined by rank_combine against training-fold references) applied to the held-out validation set. Each point is one patient, plotted along the horizontal axis by the ensemble model score and colored by true class (MS vs NMO) and correct vs. misclassified status. The gray-shaded region spans the training-fold-derived abstention zone; validation samples falling inside the zone are abstained, those outside are retained. Most misclassified validation samples fall inside or very close to the abstention zone: these are the patients for whom the ensemble's molecular signal is genuinely equivocal. Observed coverage is approximately 74.6% at 100-seed mean. On the retained subset, AUC is **91.3%** versus **88.3%** at full coverage.

##### Ensemble example 2: MS Subtyping (Figure S1b)

Figure S1b shows the same visualization for the 4-module MS Subtype ensemble (DS3 + C1 + C4 + GW1-nc; rank_combine). Observed coverage is 77.0%, slightly below the 80% target, meaning about 23% of validation samples fall in the training-derived zone. AUC at the retained 77.0% is 78.8%; at full coverage 74.7%.

##### Summary of coverage levels across primary targets

##### Choice of the 20% abstention fraction

The 20% abstention fraction is fixed *a priori* across all experiments in this paper. The motivation is clinical alignment with Oncotype DX, where approximately 30% of breast cancer patients receive an intermediate recurrence score that triggers additional evaluation rather than a direct chemotherapy decision; the TAILORx trial established this design as the standard for molecular diagnostics with an abstention zone. We chose 20% as a slightly more conservative value.

For non-screening targets we report both 100% and 80% coverage so readers see the full performance envelope. For the three primary targets, AUC at 100% coverage is 74.2%, 74.7%, and 88.3%, within or above the oncology molecular-diagnostic benchmark range of 59–80% without any abstention at all.

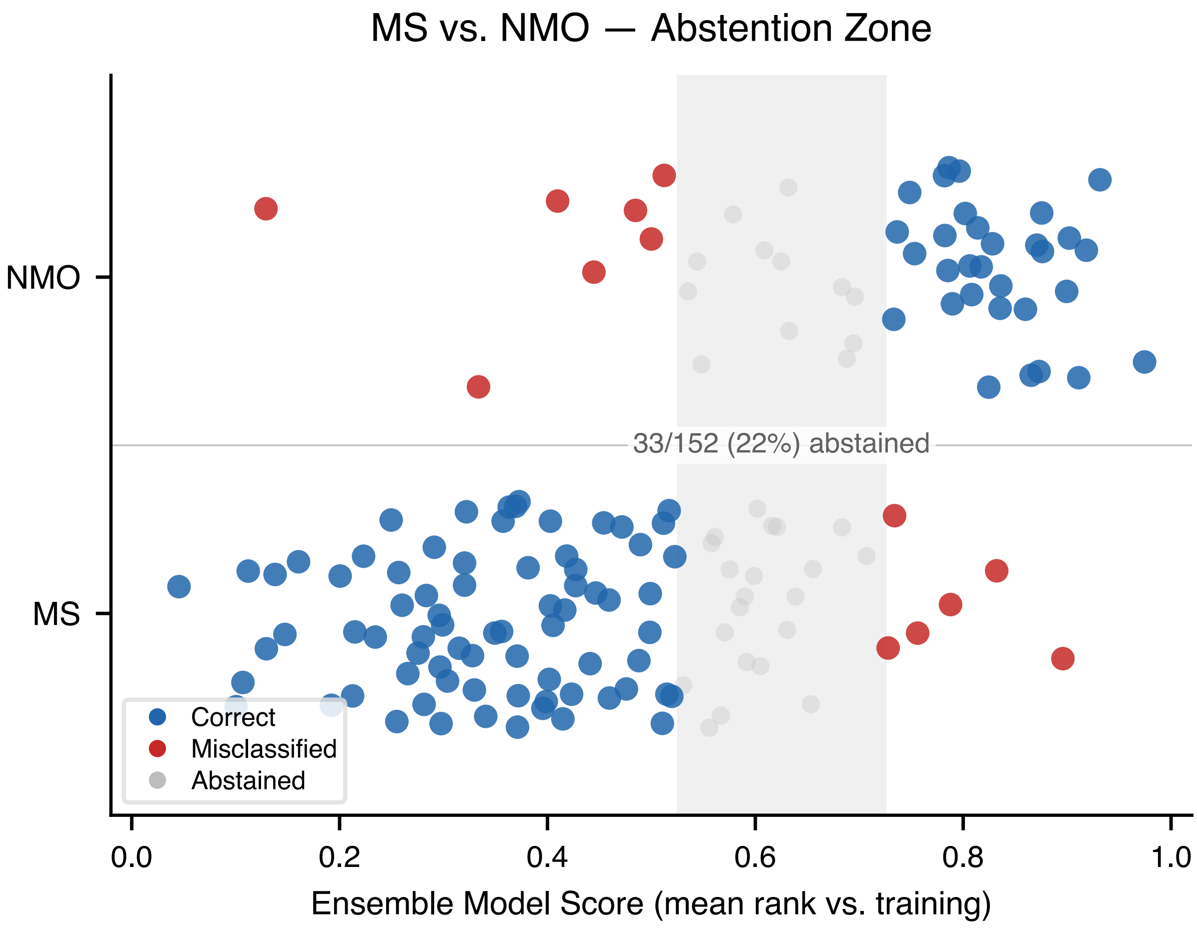

**Figure S1a.** Abstention zone for MS vs NMO differential diagnosis.

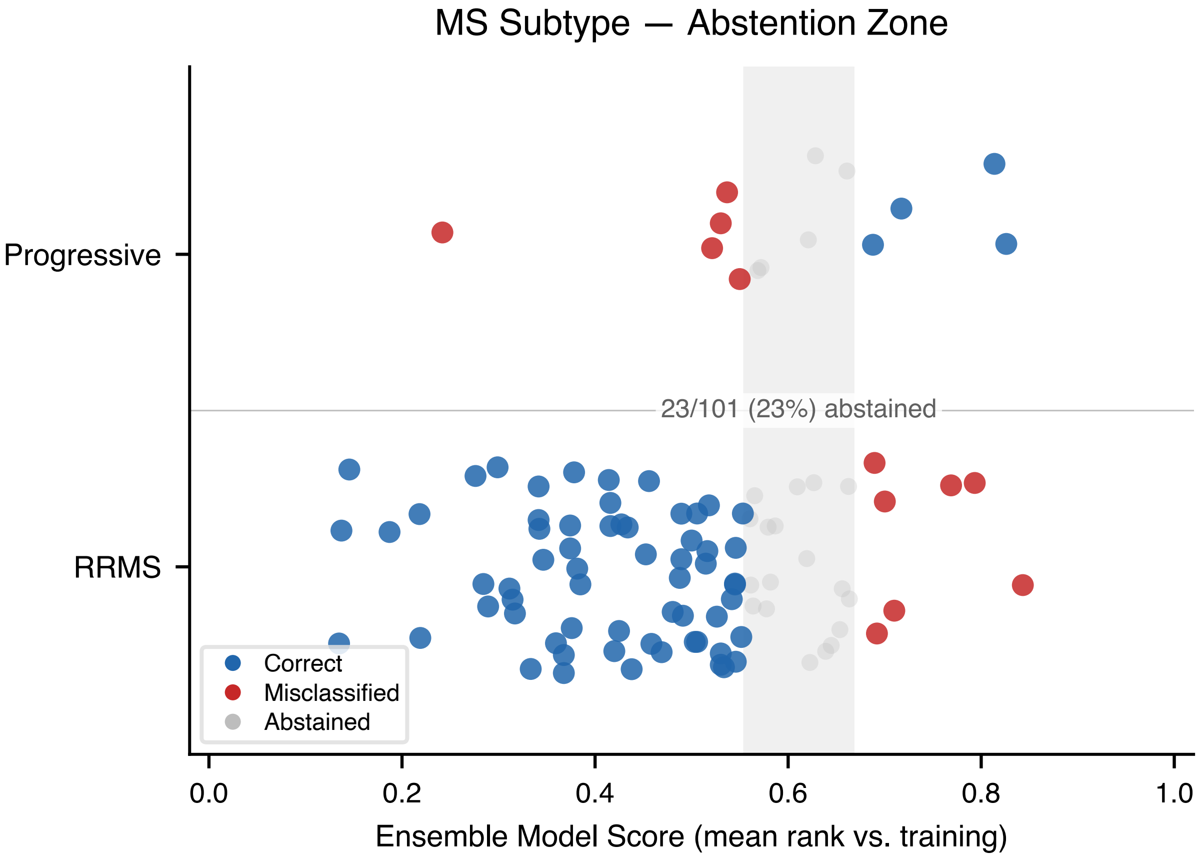

**Figure S1b.** Abstention zone for MS Subtype (progressive vs relapsing).

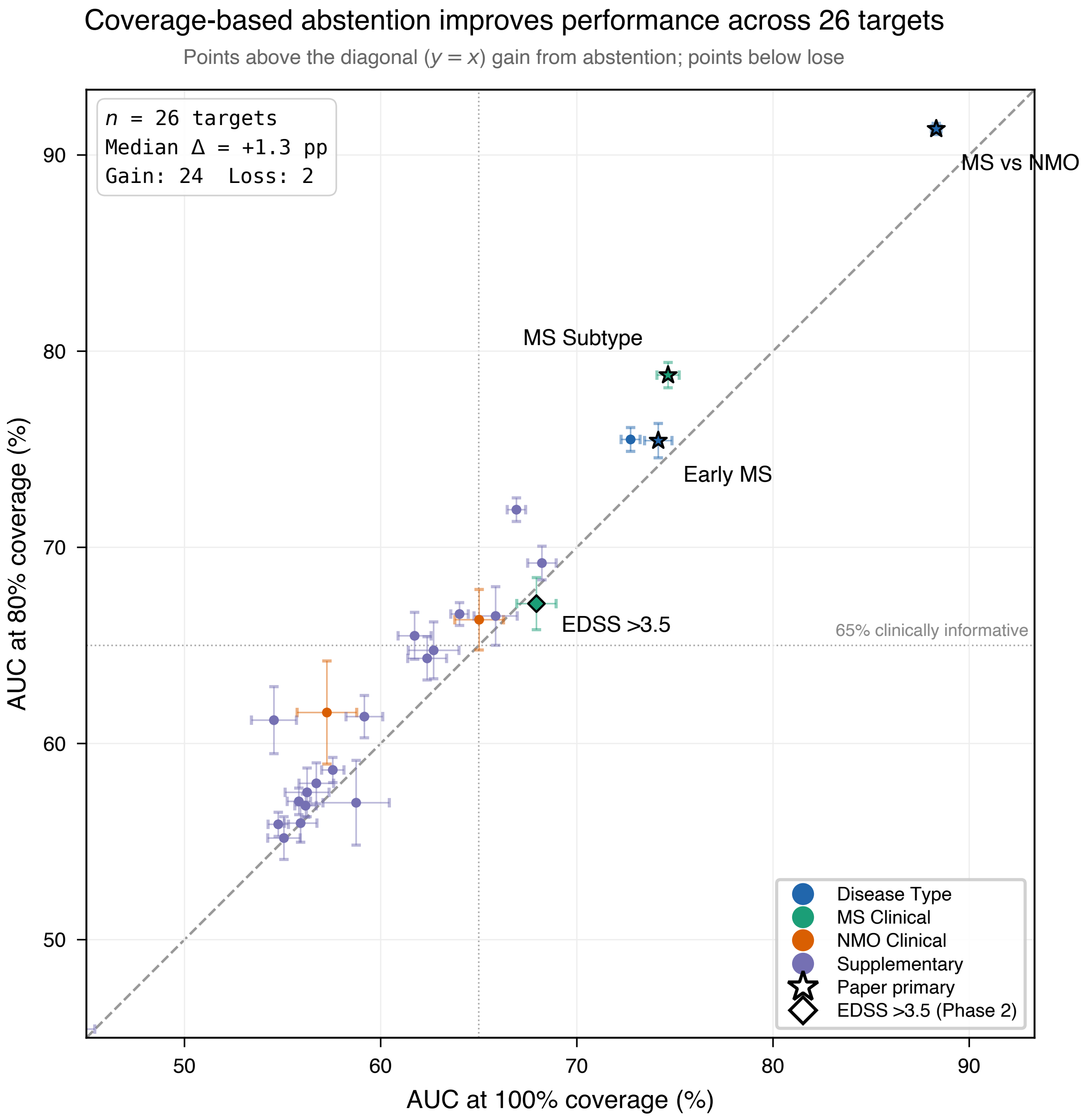

**Figure S1c.** AUC at 100% vs 80% coverage across all 26 prediction targets.

#### S2. Individual module catalog: definitions, features, and standalone performance

This section defines every input module used in the paper's ensemble classifiers. Each module is a self-contained binary classifier that consumes a distinct biologically-motivated feature set and produces a predicted probability for the target. The final ensemble for any clinical task is a rank-averaged combination of 2–8 modules, selected by the greedy search procedure described in Section S3. Readers interested only in the ensemble architecture can skim the category introductions and the solo AUC summary at the end; readers who want to understand which features drive which clinical distinctions should read the per-module entries.

Modules fall into five categories: clinical modules (C1–C4) that use curated features requiring no data-dependent gene selection; genome-wide / pooled modules (GW1, GW1-nc, P2) that apply unbiased or disease-agnostic variability filters to the full transcriptome; Disease-Sigma (DS) modules (DS1–DS8 and their non-coding variants) that rank genes by within-disease Range80 minus within-control Range80; Disease-Mean (DM) modules (DM-50, DM-200, and four pc/nc/PCA variants) that rank genes by class mean-shift, the conventional fold-change criterion underlying most prior expression biomarkers; and a naive ML baseline (PCA-64) over all viable genes. The DM family is the conventional mean-shift comparator (analogous to MammaPrint, Oncotype DX, and most prior expression biomarkers); the DS family captures variability-shifted signal that DM cannot see by construction. Empirically the two families are complementary in the greedy-selected ensembles (Fig. 3): DS modules appear in all three primary winners and are the only family selected for within-disease targets (MS Subtype winner is DS-only, EDSS >3.5 winner is DS-dominant), while between-disease winners (MS vs NMO and Early MS vs Control) each include 2 DM and 2 DS modules. The DS family is described in detail in S3 and revisited biologically in S10 (GO enrichment) and S11 (ncRNA contribution); the DM family is the conventional comparator we ablate against throughout.

Standalone AUCs reported in each per-module table are obtained from the solo-AUC matrix (30 seeds, common partition pool across every module × target pair). Values reported here are the per-seed mean AUC, multiplied by 100 and rounded to one decimal place. Entries are marked "–" where the module was not evaluated for that target (e.g., the deprecated P1 pooled module, retained in the library only as an ablation reference). The full 23-module × 13-target matrix is shown visually in main paper Fig. 3a, where cell color encodes solo AUC @80% (clear → green alpha ramp, saturated at 80%) and a thick black border marks cells whose module belongs to that target's best ensemble.

#### Table S2-1. Input module definitions for the ACP PBMC classifier

Each row defines one input module used in the modular ensemble classifier. Modules are grouped by type: Clinical (C), Genome-wide / pooled (GW, P), Disease-Sigma (DS, the variability-based family), Disease-Mean (DM, the conventional baseline family), and Naive ML. Non-coding RNA variants (-nc) share the same gene-selection pipeline as their protein-coding counterpart but draw from the non-coding RNA transcript pool only. Per-module rationale, model hyperparameters, and standalone performance are detailed in the subsections below.

#### Clinical modules (C1–C4): fixed features, no gene selection

The four clinical modules use feature sets predefined from biology or demography, without any training-data-dependent gene selection. Three (C2, C3, C4) use curated marker gene sets and the fourth (C1) uses non-expression metadata. They function as the fixed-feature baseline against which the data-driven modules are compared.

##### C1, Demographics

C1 is the non-transcriptomic baseline. Every MS and NMO classifier should beat what a clinician can guess from age, sex, and smoking history alone; C1 quantifies that floor. It also serves as the anchor for cross-disease transfer experiments (Section S9), where the question is whether transcriptomic modules add signal beyond what demographic covariates already provide.

Three metadata fields per sample: age at diagnosis (median-imputed when missing from the CRF), tobacco use (binary, imputed to 0 when missing), and sex inferred from XIST expression by median-splitting within

| Paper Name | Type | Input Features | Dimensionality Reduction | Description |
|---|---|---|---|---|
| C1 | Clinical | Age, tobacco use, sex (XIST-inferred) | None (3 features used directly) | Demographic and exposure covariates |
| C2 | Clinical | 12 cell-type ratios (hierarchical deconvolution) | None (ratios used directly) | Immune cell composition from marker gene sets |
| C3 | Clinical | 16 HLA / ancestry gene expression levels | PCA (top 5 components) | Ancestry and HLA variation captured via PCA |
| C4 | Clinical | Shannon entropy, Simpson index, evenness | None (3 indices used directly) | Immune repertoire diversity metrics |
| GW1 | Genome-wide | Top 200 protein-coding genes by F2 consistency filter | PCA (top 5 components) | Genes with reproducible variance across two independent folds |
| GW1-nc | Genome-wide | Top non-coding RNA genes by F2 consistency filter | PCA (top 5 components) | Non-coding RNA counterpart of GW1 |
| P1 | Pooled | Top 200 genes by pooled Range80 (all samples) | PCA (top 5 components) | Disease-agnostic high-variance genes (deprecated; ablation only) |
| P2 | Pooled | F2 filter applied to the pooled DS>1 gene pool | PCA (top 5 components) | Pooled-variability variant retained for the NMO-MRI ensemble |
| DS1 | DS (disease-Sigma) | Top 200 protein-coding genes by MS-specific Range80 | Direct LR (C=0.1) | Genes most variable specifically in MS samples |
| DS1-nc | DS | Top non-coding RNA genes by MS-specific Range80 | Direct LR (C=0.1) | Non-coding RNA counterpart of DS1 |
| DS2 | DS | Top 200 protein-coding genes by NMO-specific Range80 | Direct LR (C=0.1) | Genes most variable specifically in NMO samples |
| DS2-nc | DS | Top non-coding RNA genes by NMO-specific Range80 | Direct LR (C=0.1) | Non-coding RNA counterpart of DS2 |
| DS3 | DS | Top 50 protein-coding NMO-stratified genes | Direct LR (C=0.1) | Compact NMO panel; strongest solo for MS Subtype |
| DS3-nc | DS | Top 50 non-coding NMO-stratified genes | Direct LR (C=0.1) | Non-coding counterpart of DS3 |
| DS4 | DS | Top 500 protein-coding NMO-stratified genes | PCA (top 5 components) | Broader NMO variance pool; strongest solo for EDSS >3.5 |
| DS5 | DS | Top 200 protein-coding MS-stratified genes | PCA (top 10 components) | Higher-rank PCA extraction from the MS-variable pool |
| DS5-nc | DS | Top 200 non-coding MS-stratified genes | PCA (top 10 components) | Non-coding counterpart of DS5 |
| DS6 | DS | Top 200 protein-coding MS-stratified genes | PCA (top 5 components) | Lower-rank PCA extraction of the DS5 pool; enters Early MS ensemble |
| DS6-nc | DS | Top 200 non-coding MS-stratified genes | PCA (top 5 components) | Non-coding counterpart of DS6 |
| DS7 | DS | Top 50 protein-coding MS-stratified genes | Direct LR (C=0.1) | Compact MS panel counterpart of DS3; enters Early MS ensemble |
| DS7-nc | DS | Top 50 non-coding MS-stratified genes | Direct LR (C=0.1) | Non-coding counterpart of DS7 |
| DS8 | DS | Top 500 protein-coding NMO-stratified genes | PCA (top 10 components) | Higher-rank PCA extraction of the DS4 pool; enters Early MS ensemble (cross-disease transfer, see §S9) |
| DS8-nc | DS | Top 500 non-coding NMO-stratified genes | PCA (top 10 components) | Non-coding counterpart of DS8 |
| DM-50 | DM (disease-Mean) | Top 50 genes by Abs(mean(pos) − mean(neg)) | Direct LR (C=0.1) | Compact mean-shift panel |
| DM-200 | DM | Top 200 genes by Abs(mean(pos) − mean(neg)) | Direct LR (C=0.1) | Conventional fold-change baseline (all viable genes) |
| DM-200 (pc) | DM | Top 200 protein-coding DM genes | Direct LR (C=0.1) | Protein-coding-only DM pool |
| DM-200 (nc) | DM | Top 200 non-coding RNA DM genes | Direct LR (C=0.1) | Non-coding-only DM pool |
| DM-200 PCA | DM | Top 200 DM genes | PCA (top 5 components) | DM with PCA reduction (matches DS1 architecture) |
| DM-500 PCA | DM | Top 500 DM genes | PCA (top 5 components) | Broader DM pool with PCA reduction |
| PCA-64 | Naive ML | All 46,722 viable genes | PCA (top 64 components) | Naive ML baseline; performs at chance on all 3 primary targets |

**C1, Demographics**

| Target | AUC @100% | AUC @80% |
|---|---|---|
| MS Subtype | 67.1% | 69.6% |
| Early MS Detection | 56.1% | 56.6% |
| MS vs NMO | 59.6% | 59.5% |
| EDSS >3.5 | 57.9% | 59.6% |

the training fold. XIST-based sex inference avoids the missing-data problem that plagues self-reported sex in the ACP cohort.

Logistic regression with L2 regularization (C=1.0) on the three standardized features.

*Standalone performance (30-seed mean):*

Demographics contribute most to MS Subtype (where age-of-onset and smoking history correlate with progressive phenotype) and least to Early MS detection and MS vs NMO. C1 appears in nearly every winning ensemble despite weak solo AUCs, consistent with demographic signal being largely orthogonal to the transcriptomic modules.

##### C2, Hierarchical immune composition

Bulk PBMC RNAseq is a mixture of cell types in proportions that vary across patients. A naive bulk classifier conflates "this patient has more CD4+ T cells" with "this patient's CD4+ T cells express gene X differently." C2 addresses this by computing explicit cell-type composition features from curated marker genes and leaving gene-level expression to other modules.

Twelve ratio features computed as relative abundance ratios across six cell types (CD4+ T, CD8+ T, B, NK, monocyte, neutrophil), using curated marker gene sets from Newman et al. 2015 (CIBERSORT LM22-adjacent). Ratios rather than raw abundances are used to normalize out overall library size.

Logistic regression with strong L2 regularization (C=0.001) on the 12 standardized ratio features.

*Standalone performance:*

C2 is the strongest fixed-feature module for MS vs NMO, 83.2% @80% from cell composition alone, consistent with the B-cell and plasmablast expansion reported in NMO relative to MS. For Early MS detection C2 is at chance, suggesting early-disease pathology is not driven by bulk compositional shifts at the resolution captured here.

##### C3, Ancestry and HLA

MS genetic risk is dominated by the HLA-DRB1*15:01 haplotype, and ancestry is a known confounder in any MS biomarker study. C3 captures both by using expression levels of 16 HLA and ancestry-informative genes as features.

**C2, Hierarchical immune composition**

| Target | AUC @100% | AUC @80% |
|---|---|---|
| MS Subtype | 65.1% | 67.1% |
| Early MS Detection | 52.1% | 51.7% |
| MS vs NMO | 79.2% | 83.2% |
| EDSS >3.5 | 58.6% | 59.9% |

**C3, Ancestry and HLA**

| Target | AUC @100% | AUC @80% |
|---|---|---|
| MS Subtype | 52.8% | 53.7% |
| Early MS Detection | 60.0% | 61.4% |
| MS vs NMO | 59.7% | 60.3% |
| EDSS >3.5 | 46.0% | 45.1% |

Sixteen HLA and ancestry genes (HLA-A, HLA-B, HLA-C, HLA-DRB1, HLA-DQA1, HLA-DQB1, and ten ancestry-informative marker genes). PCA with the top 5 components used as features.

Logistic regression with L2 regularization (C=0.1) on the 5 HLA/ancestry principal components.

*Standalone performance:*

C3 contributes to Early MS detection (61.4% @80%, plausibly via HLA-DRB1*15:01 risk variation) and is at chance for subtype or disability prediction. It is selected for the Early MS winning ensemble as a small but non-redundant signal source.

##### C4, Immune diversity

Shannon entropy, Simpson index, and evenness computed over the 12 C2 cell-fraction features measure immune-compartment diversity. Reduced diversity is associated with autoimmune disease in general and with NMO in particular.

Three diversity indices computed on the C2 cell-fraction estimates (six cell types, 46 marker genes).

Logistic regression with L2 regularization (C=1.0 for within-MS targets, C=0.1 for between-disease targets) on the three standardized diversity indices.

*Standalone performance:*

C4 is the strongest solo module for EDSS >3.5 (66.9% @80%) and is selected for the EDSS >3.5 winning ensemble. One interpretation is that disability tracks immune-repertoire contraction; we cannot distinguish

**C4, Immune diversity**

| Target | AUC @100% | AUC @80% |
|---|---|---|
| MS Subtype | 64.9% | 65.3% |
| Early MS Detection | 53.4% | 53.0% |
| MS vs NMO | 76.9% | 79.6% |
| EDSS >3.5 | 66.9% | 66.9% |

this from a simpler cell-composition correlate without independent repertoire sequencing. For MS vs NMO it reaches 79.6% @80%, within 4% of C2, plausibly driven by reduced BCR/TCR repertoire diversity reported in NMO.

#### Genome-wide / pooled modules (GW1, GW1-nc, P1, P2)

These modules apply unbiased or disease-agnostic gene selection to the full transcriptome, providing data-driven baselines for comparison against the DM and DS families. GW1 applies an AUC-based feature filter using training-fold labels only (no test-fold consistency check is used, for deployment-invariance, see §S1), selecting the top-200 protein-coding genes by directional-AUC strength and reducing to five principal components. P1 and P2 rank genes by pooled variability without disease stratification; P2 uses the same training-only AUC filter on the DS>1 gene pool.

##### GW1, Genome-wide protein-coding

GW1 is the unbiased transcriptomic baseline: protein-coding genes selected by training-fold univariate-AUC consistency, with no disease-stratified variability filter and no DM-style mean-shift ranking.

Top 200 protein-coding genes by F2 consistency filter (univariate AUC > 0.55 or < 0.45 on training fold; direction-consistent on the independent test fold, AUC > 0.52 or < 0.48, log2 shrinkage ratio < 0.5), selected per seed using only the training-accessible subset.

PCA to the top 5 components, then logistic regression with L2 regularization (C=0.1).

*Standalone performance:*

GW1 trails the best DM or DS module on any single target but stays within 5–10% across all of them. It is selected for the MS Subtype winning ensemble and for the NMO_New_MRI ensemble.

##### GW1-nc, Genome-wide non-coding RNA

Same F2 consistency filter as GW1, but applied to the non-coding RNA subspace. The BIRT platform captures lncRNA, snoRNA, and miRNA species that are absent from poly-A-selected RNAseq and below the microarray detection floor (Section S13). GW1-nc tests whether the ncRNA transcriptome carries independent classification signal.

Top non-coding RNA genes by F2 consistency filter, with the same per-seed training-fold-only selection

##### GW1, Genome-wide protein-coding

| Target | AUC @100% | AUC @80% |
|---|---|---|
| MS Subtype | 61.7% | 63.1% |
| Early MS Detection | 69.0% | 70.6% |
| MS vs NMO | 81.6% | 84.8% |
| EDSS >3.5 | 53.9% | 55.1% |

##### GW1-nc, Genome-wide non-coding RNA

| Target | AUC @100% | AUC @80% |
|---|---|---|
| MS Subtype | 58.7% | 59.4% |
| Early MS Detection | 59.8% | 60.7% |
| MS vs NMO | 79.7% | 82.5% |
| EDSS >3.5 | 56.0% | 56.1% |

as GW1.

Identical to GW1 (PCA-5 + logistic regression, C=0.1).

*Standalone performance:*

GW1-nc trails GW1 by 2–10% on most targets but reaches 82.5% @80% on MS vs NMO, within 2.3% of the protein-coding genome-wide baseline. The non-coding transcriptome carries near-equivalent MS-vs-NMO signal to the protein-coding transcriptome under identical selection; Section S11 develops the implications for platform choice.

##### P1, Pooled variability, deprecated

P1 ranks the top 200 genes by pooled Range80 across the full cohort using an external GSE314922 healthy reference (n=108) for the variability subtraction. P1 is not selected by the greedy search for any target and is retained in the catalog only as a disease-agnostic-variability baseline; its per-target solo-AUC row is omitted. The DS family (disease-stratified variability) and the DM family (supervised mean-shift) together cover the design space.

##### P2, Pooled DS>1 with F2 filter

P2 is a hybrid: the F2 consistency filter applied to the pooled-variability gene pool (genes with DS > 1 across the full cohort). It tests whether reproducibility-filtered pooled variability carries signal independent

**P2, Pooled DS>1 with F2 filter**

| Target | AUC @100% | AUC @80% |
|---|---|---|
| MS Subtype | 61.6% | 63.8% |
| Early MS Detection | 69.2% | 71.2% |
| MS vs NMO | 79.6% | 82.7% |
| EDSS >3.5 | 51.5% | 50.8% |

of Disease-Sigma selection. Unlike P1, P2 appears in several secondary-target ensembles, typically when low-prevalence MRI-activity targets benefit from reproducibility-filtered variability signal alongside GW1 and a DS module.

F2-filtered subset of the pooled DS>1 gene pool, per-seed selection on training-accessible samples.

PCA-5 + logistic regression (C=0.1).

*Standalone performance:*

P2 is competitive with GW1 across the four primary-supplementary targets and matches DM-200 PCA on Early MS. It is selected by the greedy search for several low-prevalence MRI-activity targets where Disease-Sigma pools are sample-limited.

#### Disease-Sigma modules (DS1–DS8 and -nc variants): the variability-shift family

Each DS module applies a Disease-Sigma Range80 filter: genes are ranked by their 10–90 percentile expression range computed within a single disease class (MS or NMO), minus the same statistic computed in healthy controls. The resulting feature sets capture disease-specific *variability*, a signal the DM family is blind to by construction. Section S10 reports the GO programs enriched in DS-selected genes (myeloid/innate for MS-stratified pools, B-cell/humoral for NMO-stratified pools); Section S5 reports the 26-target information-boundary analysis.

##### DS1, MS-specific variance, protein-coding

Top 200 protein-coding genes by Range80(MS, corrected) − Range80(Control, corrected), selected per seed from training-fold samples only.

Direct logistic regression (C=0.1) on the 200 standardized expression values.

*Standalone performance:*

DS1 is the strongest single DS module for MS vs NMO (86.1% @80%) and is selected for the MS vs NMO winning ensemble alongside DM modules (Fig. 3). MS-specific variability in protein-coding genes (enriched for myeloid activation markers, see S10) is the most discriminative single DS feature set in the catalog. DS1 was selected from MS samples' within-class variability alone and yet classifies MS against NMO at near-ensemble accuracy, which is the empirical observation that motivates the DS approach.

**DS1, MS-specific variance, protein-coding**

| Target | AUC @100% | AUC @80% |
|---|---|---|
| MS Subtype | 57.8% | 59.2% |
| Early MS Detection | 69.0% | 69.7% |
| MS vs NMO | 82.8% | 86.1% |
| EDSS >3.5 | 61.0% | 60.9% |

**DS1-nc, MS-specific variance, non-coding RNA**

| Target | AUC @100% | AUC @80% |
|---|---|---|
| MS Subtype | 54.3% | 54.0% |
| Early MS Detection | 65.3% | 67.0% |
| MS vs NMO | 81.1% | 84.2% |
| EDSS >3.5 | 46.5% | 44.1% |

##### DS1-nc, MS-specific variance, non-coding RNA

Top non-coding RNA genes by the same MS-specific Range80 filter as DS1.

Identical to DS1.

*Standalone performance:*

The non-coding RNA version of DS1 reaches 84.2% @80% for MS vs NMO, within 2% of its protein-coding counterpart. Section S11 develops the broader argument that ncRNA-only configurations approach mRNA-only performance across all three primary targets.

##### DS2, NMO-specific variance, protein-coding

Top 200 protein-coding genes by Range80(NMO, corrected) − Range80(Control, corrected), selected per seed.

Direct logistic regression (C=0.1).

*Standalone performance:*

DS2 mirrors DS1: strong on MS vs NMO despite being derived from the opposite disease's within-group variability. One reading is that MS-variable and NMO-variable pools each surface distinct biological programs (myeloid and B-cell respectively, per S10) that both discriminate when one disease is classified against the other. DS2 is selected for the NMO_New_MRI ensemble.

##### DS2, NMO-specific variance, protein-coding

| Target | AUC @100% | AUC @80% |
|---|---|---|
| MS Subtype | 62.4% | 64.0% |
| Early MS Detection | 66.2% | 67.7% |
| MS vs NMO | 82.3% | 84.7% |
| EDSS >3.5 | 54.2% | 52.4% |

##### DS2-nc, NMO-specific variance, non-coding RNA

| Target | AUC @100% | AUC @80% |
|---|---|---|
| MS Subtype | 51.2% | 51.7% |
| Early MS Detection | 65.2% | 67.2% |
| MS vs NMO | 82.0% | 85.0% |
| EDSS >3.5 | 55.4% | 56.3% |

###### DS2-nc, NMO-specific variance, non-coding RNA

Top non-coding RNA genes by NMO-specific Range80.

Identical to DS2.

*Standalone performance:*

DS2-nc reaches 85.0% @80% on MS vs NMO, slightly above its protein-coding sibling. NMO-specific variability appears to be carried in substantial part by non-coding transcripts (B-cell-associated lncRNAs, see S11), which is consistent with the broader platform-choice argument developed there.

###### DS3, Top-50 NMO-stratified, protein-coding

Top 50 genes by NMO-specific Range80, selected per seed from the protein-coding pool.

Direct logistic regression (C=0.1) on the 50 standardized expression values.

*Standalone performance:*

DS3 is the strongest DS solo module for MS Subtype (67.6% @80%) and is selected for the MS Subtype winning ensemble alongside C1, C4, and GW1, despite being selected on NMO biology. Section S9 develops this cross-disease transfer further; one candidate explanation is that progressive MS involves meningeal B-cell inflammation overlapping with NMO B-cell biology, which the compact 50-gene panel surfaces more sharply than the broader 200-gene DS2 pool.

##### DS3, Top-50 NMO-stratified, protein-coding

| Target | AUC @100% | AUC @80% |
|---|---|---|
| MS Subtype | 65.8% | 67.6% |
| Early MS Detection | 59.6% | 60.7% |
| MS vs NMO | 82.9% | 85.4% |
| EDSS >3.5 | 58.1% | 58.0% |

##### DS3-nc, Top-50 NMO-stratified, non-coding RNA

| Target | AUC @100% | AUC @80% |
|---|---|---|
| MS Subtype | 54.4% | 55.4% |
| Early MS Detection | 60.8% | 61.6% |
| MS vs NMO | 82.0% | 84.8% |
| EDSS >3.5 | 54.8% | 53.6% |

**DS3-nc, Top-50 NMO-stratified, non-coding RNA**

Top 50 non-coding RNA genes by NMO-specific Range80.

Direct LR (C=0.1).

*Standalone performance:*

DS3-nc reaches MS vs NMO performance comparable to its pc counterpart but loses the MS Subtype edge of DS3. The cross-disease MS Subtype signal may reside in protein-coding B-cell receptor genes rather than ncRNA, though the compact 50-gene cutoff also leaves less room for ncRNA hits to register. DS3-nc is selected by the greedy search for the Walk>7_MS secondary target.

**DS4, NMO 500-gene PCA**

Top 500 protein-coding genes by NMO-specific Range80, broader than DS2/DS3.

PCA-5 + logistic regression (C=0.1).

*Standalone performance:*

DS4 is the strongest DS solo module for EDSS >3.5 disability prediction (64.1% @80%), a result that is counterintuitive given DS4 is selected on NMO biology. One possibility is that MS disability tracks B-cell-associated features (which would align with ocrelizumab efficacy in progressive MS and the Magliozzi meningeal-follicle literature); we cannot distinguish this from cross-disease feature transfer that does not

**DS4, NMO 500-gene PCA**

| Target | AUC @100% | AUC @80% |
|---|---|---|
| MS Subtype | 61.1% | 62.6% |
| Early MS Detection | 64.2% | 65.7% |
| MS vs NMO | 80.8% | 84.0% |
| EDSS >3.5 | 63.1% | 64.1% |

**DS5, MS 200-gene PCA**

| Target | AUC @100% | AUC @80% |
|---|---|---|
| MS Subtype | 56.4% | 56.7% |
| Early MS Detection | 69.5% | 71.0% |
| MS vs NMO | 81.2% | 84.2% |
| EDSS >3.5 | 59.2% | 59.5% |

require a shared mechanism. The EDSS >3.5 winning ensemble is C4 + C1 + DS7, with DS4 retained in the candidate pool for secondary-target screening.

##### DS5, MS 200-gene PCA

Top 200 protein-coding MS-stratified genes; alternative architecture to DS1 (PCA-5 instead of direct LR).

PCA-5 + logistic regression (C=0.1).

*Standalone performance:*

DS5 is the PCA-reduced sibling of DS1; it reaches 71.0% @80% on Early MS (where direct-LR DS1 reaches 69.7%) and stays within 3% of DS1 on the other primary targets. DS5 is selected for the MS vs Healthy Control winning ensemble (DS5 + DM-200(nc) + DM-200(pc) + C3 + DS1), pairing a PCA-compressed MS-variable representation with the direct-LR DM and DS1 modules.

##### DS5-nc, MS-variable non-coding RNA

Non-coding RNA counterpart of DS5.

PCA-5 + LR (C=0.1).

*Standalone performance:*

DS5-nc trails DS1-nc by 1–3% on most targets, suggesting direct LR on the 200-gene pool extracts the

**DS5-nc, MS-variable non-coding RNA**

| Target | AUC @100% | AUC @80% |
|---|---|---|
| MS Subtype | 58.5% | 59.8% |
| Early MS Detection | 63.7% | 65.0% |
| MS vs NMO | 78.4% | 81.4% |
| EDSS >3.5 | 56.7% | 57.4% |

variability signal more efficiently than PCA at this dimensionality. DS5-nc is selected by the greedy search for several secondary targets (Signs>3_MS, Sym>10_MS).

##### Disease-Mean modules (DM-50 to DM-500 PCA): the conventional baseline family

The DM family ranks genes by Abs(mean(positive class) − mean(negative class)) computed per seed on training-accessible samples, with healthy controls used as the reference for between-disease targets. This is the conventional fold-change-style criterion underlying most prior expression biomarkers (MammaPrint, Oncotype DX, Afirma) and is the comparator against which the DS family is ablated. Six DM variants span gene-pool size (50, 200, 500), transcript subspace (all viable, protein-coding only, non-coding only), and dimensionality reduction (direct LR vs. PCA-5).

###### DM-200, All viable genes, direct LR

The reference DM module: top 200 genes ranked by Abs(mean(pos) − mean(neg)) from the full 46,722-gene viable pool. DM-200 is architecturally matched to DS1 (same dimensionality, same classifier, different selection criterion), so direct comparison of the two quantifies the marginal signal carried by mean-shift versus variability-shift.

Top 200 genes by class mean-shift, per-seed selection.

Direct LR (C=0.1).

*Standalone performance:*

DM-200 is the strongest single solo module for both Early MS (72.8% @80%) and MS vs NMO (87.6% @80%) and is selected for both the Early MS and MS vs NMO winning ensembles. For within-MS targets (MS Subtype, EDSS >3.5) DM-200 is mid-pack and the winning ensembles for those targets are DS-only or DS-dominant.

###### DM-50, Compact mean-shift panel

Top 50 genes by class mean-shift, all viable genes pooled, per-seed selection.

Direct LR (C=0.1).

*Standalone performance:*

##### DM-200, All viable genes, direct LR

| Target | AUC @100% | AUC @80% |
|---|---|---|
| MS Subtype | 57.2% | 59.0% |
| Early MS Detection | 71.3% | 72.8% |
| MS vs NMO | 84.5% | 87.6% |
| EDSS >3.5 | 56.8% | 56.4% |

##### DM-50, Compact mean-shift panel

| Target | AUC @100% | AUC @80% |
|---|---|---|
| MS Subtype | 59.5% | 61.3% |
| Early MS Detection | 72.7% | 74.4% |
| MS vs NMO | 83.1% | 86.1% |
| EDSS >3.5 | 56.1% | 55.8% |

DM-50 is the strongest single solo module across the catalog for Early MS Detection (74.4% @80%) and is selected for the Early MS winning ensemble alongside DS8, DM-200, C3, DM-200 (nc), DS6, and PCA-64. The compact 50-gene panel slightly outperforms DM-200 on Early MS and slightly underperforms on MS vs NMO, plausibly because mean-shift signal is concentrated in a small number of high-effect-size genes for Early MS while MS vs NMO benefits from the broader 200-gene pool.

**DM-200 (pc), Protein-coding only**

Top 200 DM genes restricted to the 18,746 protein-coding viable genes.

Direct LR (C=0.1).

*Standalone performance:*

Protein-coding-only DM trails the all-genes DM-200 by ~1% on MS vs NMO and Early MS, since protein-coding here is a strict subset rather than a separate pool. DM-200 (pc) is selected for the MS vs Healthy Control winning ensemble, pairing with DM-200 (nc) to span coding/non-coding fold-change space alongside DS1 and DS5.

**DM-200 (nc), Non-coding only**

Top 200 DM genes restricted to the 27,976 non-coding viable genes.

**DM-200 (pc), Protein-coding only**

| Target | AUC @100% | AUC @80% |
|---|---|---|
| MS Subtype | 55.5% | 56.5% |
| Early MS Detection | 70.2% | 71.3% |
| MS vs NMO | 83.7% | 86.5% |
| EDSS >3.5 | 59.4% | 58.6% |

**DM-200 (nc), Non-coding only**

| Target | AUC @100% | AUC @80% |
|---|---|---|
| MS Subtype | 54.4% | 55.1% |
| Early MS Detection | 69.5% | 70.3% |
| MS vs NMO | 84.3% | 87.2% |
| EDSS >3.5 | 49.3% | 49.6% |

Direct LR (C=0.1).

*Standalone performance:*

DM-200 (nc) reaches 87.2% @80% on MS vs NMO, within 0.4% of all-genes DM-200 and within 0.7% of the protein-coding DM. The non-coding transcriptome alone, ranked by mean-shift, classifies MS vs NMO at near-ensemble accuracy. Section S11 develops the implications for ncRNA capture as a platform requirement. DM-200 (nc) is selected for both the MS vs NMO and Early MS winning ensembles.

##### DM-200 PCA, DM-200 with PCA-5 reduction

Top 200 DM genes (all viable), reduced to 5 principal components.

PCA-5 + LR (C=0.1).

*Standalone performance:*

DM-200 PCA trails direct-LR DM-200 by ~2% on MS vs NMO. At this gene-pool size and sample count, direct LR extracts the mean-shift signal more efficiently than PCA-5. The PCA architecture is included in the catalog as the matched-architecture comparator against GW1, DS4, and DS5.

##### DM-500 PCA, Broader DM pool with PCA-5

Top 500 DM genes (all viable), reduced to 5 principal components.

**DM-200 PCA, DM-200 with PCA-5 reduction**

| Target | AUC @100% | AUC @80% |
|---|---|---|
| MS Subtype | 56.4% | 58.0% |
| Early MS Detection | 69.3% | 71.7% |
| MS vs NMO | 82.7% | 85.6% |
| EDSS >3.5 | 59.6% | 60.1% |

**DM-500 PCA, Broader DM pool with PCA-5**

| Target | AUC @100% | AUC @80% |
|---|---|---|
| MS Subtype | 57.1% | 58.2% |
| Early MS Detection | 69.0% | 70.6% |
| MS vs NMO | 81.9% | 84.8% |
| EDSS >3.5 | 59.2% | 58.9% |

PCA-5 + LR (C=0.1).

*Standalone performance:*

Doubling the gene pool from 200 to 500 under PCA-5 adds noise without adding signal: DM-500 PCA matches or slightly trails DM-200 PCA on every target. Retained as an ablation reference for the gene-pool sweep.

##### Naive ML baseline (PCA-64)

###### PCA-64, Whole-transcriptome PCA, no feature selection

PCA-64 is the naive ML baseline: PCA to 64 components on the full 46,722 viable gene matrix (no F2 filter, no disease stratification, no DM/DS ranking), followed by logistic regression. It quantifies what a generic dimensionality-reduction pipeline achieves without the biology-aware feature selection used by the rest of the catalog.

All 46,722 viable genes; PCA to 64 components per seed on training-fold samples.

Logistic regression (C=0.1) on the 64 PC scores.

*Standalone performance:*

PCA-64 performs at chance on MS Subtype and EDSS >3.5 (≤ 51% AUC) and reaches only 57.0–58.5% @80% on Early MS and MS vs NMO. Sixty-four principal components on 46,722 genes do not extract clinically useful solo signal on any primary target. PCA-64 is nonetheless selected by the greedy search for

**PCA-64, Whole-transcriptome PCA, no feature selection**

| Target | AUC @100% | AUC @80% |
| --- | --- | --- |
| MS Subtype | 50.2% | 50.5% |
| Early MS Detection | 56.2% | 57.0% |
| MS vs NMO | 58.1% | 58.5% |
| EDSS >3.5 | 50.0% | 49.9% |

the Early MS winning ensemble, indicating that a non-trivial residual remains after the stronger modules dominate.

##### Summary: solo performance matrix

The 23-module × 13-target solo performance matrix is shown in main paper Fig. 3a. Cell color encodes solo AUC @80%, with a thick black border marking cells whose module belongs to that target's best ensemble. Five patterns emerge. First, no single module leads on all targets; DM-200 leads between-disease targets, C1 demographics lead MS subtyping, and C4 immune diversity leads EDSS disability. Second, DS modules appear in all three primary winning ensembles while DM appears in two of three (the two between-disease targets), with the within-disease MS Subtype winner selecting no DM module. Third, the naive ML baseline (PCA-64) performs at chance on all three primaries; the feature-selection pipelines that define DM and DS produce signal that 64-component whole-transcriptome PCA does not. Fourth, fixed-feature clinical modules (C1–C4) contribute above chance on nearly every target and anchor specific ensembles (C1 for MS Subtype and Early MS, C4 for EDSS >3.5, C2 as a solo contender for MS vs NMO at cell-composition-only performance) but rarely lead. Fifth, non-coding RNA variants trail their protein-coding counterparts by 0–5% on most targets; the gap is small enough that ncRNA-only configurations reach clinically useful AUCs, and ncRNA variants (DM-200(nc), DS1-nc, DS3-nc, DS5-nc, GW1-nc) enter winning ensembles across multiple targets (Section S11).

The greedy selection procedure described in Section S3 combines these modules into target-specific winning configurations. The resulting ensembles exceed every constituent module's solo AUC by 3–8%, indicating that the modules carry complementary rather than redundant signal.

#### S3. Module selection: greedy forward search with per-target combiner choice

##### Why greedy

Given the full module library described in Section S2 (33 candidate modules: 5 clinical/genome-wide fixed modules, 6 Differential-Mean variants, 22 Disease-Sigma modules across the protein-coding and non-coding gene pools), the number of possible non-empty ensemble subsets is approximately $2^{33} \approx 8.6 \times 10^9$. Evaluating every subset at 30 seeds × 2 ensemble-combiner methods × 26 clinical prediction targets is computationally infeasible. **Greedy forward selection** reduces this to $O(n^2)$ candidate evaluations per (target, combiner) search, for a 33-module pool the worst case is $\sum_{k=1}^{33}(33 - k + 1) = 561$ candidate evaluations at 30 seeds.

Across the full 26-target × 2-combiner search, the total compute is about 45 minutes of real time once module-prediction caching is in place.

The cost of the greedy approximation is that the selected configuration is not guaranteed to be globally optimal. The goal here is not the theoretically-best ensemble for each target (marginal ensemble improvements are bounded above by the per-seed noise floor of SEM ~0.3–0.7% on the primary targets and dominated by cohort-size effects), but a module list that is (a) **reproducible**: the same greedy procedure on the same seed pool converges deterministically, (b) **deployment-invariant**: every module and every ensemble-combiner is a pure function of (input sample, frozen model artifacts), and (c) **biologically interpretable**: the selected modules individually index clinically-meaningful gene programs (Sections S2, S10, S11, S17).

#### Two-phase procedure with disjoint seed pools

We use a two-phase design with disjoint seed pools to prevent selection-bias inflation of the reported AUCs. Phase 1 screening uses seeds 2000-2029 : for each (target, combiner) pair, greedy forward selection runs over the full 33-module pool, adding at each step the module whose inclusion most improves the 30-seed mean of the target's optimization metric. Phase 2 validation uses seeds 0-99 : for each target, we re-evaluate the single winning (combiner, module list) from Phase 1, where the per-target winner is the combiner whose Phase 1 metric is higher. The 100-seed means and SEMs reported in Section S5 come from Phase 2.

The two phases draw from **non-overlapping random partitions of the data**, so the headline numbers reported in Section S5 cannot be inflated by greedy adaptation to selection-seed noise.

#### Greedy algorithm

For each (target, combiner):

1. **Pre-compute module predictions.** For each of the 30 screening seeds, fit every module in the 33-module pool once on the training fold, and store the per-sample predictions on both the training and validation folds. This is done once per target; greedy then operates on cached predictions only and never refits a module. 2. **Seed the ensemble.** For each candidate solo module, evaluate the 30-seed mean of the target optimization metric using the chosen ensemble combiner (rank_combine or conviction) applied to that one module's cached predictions. The module with the highest mean metric becomes the initial ensemble. 3. **Forward selection.** At each step, for each candidate module not yet in the ensemble, evaluate the 30-seed mean of the metric for the ensemble consisting of (currently-selected {candidate}). Add the candidate with the highest mean metric if the improvement exceeds $\epsilon = 0.001$; otherwise terminate. 4. **Output.** The final ensemble module list, the target's 30-seed mean metric at that ensemble, and the full trajectory of solo → +1 → +2 →... metric values.

Caching is required for the full-pool single-pass search to be tractable: with cached predictions, each greedy step operates on numpy arrays and completes in milliseconds; without caching, each candidate evaluation would re-fit every module from scratch, and the search would cost several hours per target.

#### Optimization metric: AUC @80% in general, @100% for screening targets

The greedy procedure optimizes a single per-target metric. For targets where abstention is clinically natural (confirmatory differential diagnosis, longitudinal monitoring, most secondary clinical endpoints), the metric is **AUC @80% coverage** (§S1 defines the deployment-invariant abstention procedure). For the Early MS screening target, abstention is not clinically acceptable, so the greedy optimizes **AUC @100% coverage**. Other targets are reported at both @100% and @80% in Section S5.

| Target | n | Combiner | Modules | AUC @100% | AUC @80% |
|---|---|---|---|---|---|
| MS vs NMO | 761 | Rank | DM-200 + DM-200(nc) + C2 + DS1 + C1 + DS1-nc | 0.883 ± 0.002 | 0.913 ± 0.003 |
| MS Subtype | 508 | Rank | DS3 + C1 + C4 + GW1-nc | 0.747 ± 0.006 | 0.788 ± 0.006 |
| Early MS vs Control | 246 | Rank | DM-50 + DS8 + DM-200 + C3 + DM-200(nc) + DS6 + PCA-64 | 0.742 ± 0.007 | – (screening) |

#### Per-target combiner choice

The paper evaluates two deployment-invariant ensemble combiners (Section S2 defines them formally). Rank-combine ranks the candidate sample's per-module score against the saved distribution of that module's training-fold scores (via `np.searchsorted` semantics), and takes the mean of the per-module ranks as the ensemble score. Conviction-weighted mean computes a per-sample conviction weight $w = \max(-\log_{10} p, -\log_{10}(1-p))$, large when the module is confident in either direction and small when the module is near the decision boundary, and takes the conviction-weighted mean of raw module probabilities as the ensemble score.

We run greedy separately under each combiner for each target and pick the per-target winner. Across all 26 targets evaluated at the 30-seed screening stage, **rank-combine wins 18 targets, conviction wins 7, and one is essentially tied**. For the three paper-primary targets, rank-combine is the selected combiner in every case, with margins of 3–4% over conviction on MS Subtype, 0.6–1.2% on Early MS, and 0.8% on MS vs NMO. The conviction-wins are concentrated on smaller-cohort and lower-prevalence targets (some NMO clinical endpoints at n = 77–181, and EDSS > 2) where raw-probability weighting outperforms rank normalization, plausibly because rank normalization in small samples amplifies ties at the boundary.

#### Phase 2 validation: 100-seed numbers are the headline

Phase 2 re-runs the entire pipeline (module fit, ensemble combination, abstention zone) at 100 disjoint seeds (0–99) for the selected (combiner, module list) per target. The reported Section S5 AUC values are always the Phase 2 100-seed means and their standard errors, never the Phase 1 screening values.

Phase 1 → Phase 2 shrinkage on the primary targets is small: MS vs NMO 0.918 → 0.913 (−0.5%), MS Subtype 0.776 → 0.788 (+1.2%, validation AUC slightly higher than screening, within-SEM noise), Early MS @100% 0.762 → 0.742 (−2.0%). The overall pattern is consistent with the expected modest selection-bias correction from disjoint seed pools: Phase 1 estimates are slightly optimistic on average because the greedy procedure adapts to screening-seed noise, and Phase 2 on disjoint seeds removes that bias. Targets with very small n (< 100) show larger shrinkage, and two smaller NMO targets (OCB_NMO at n = 87, IgG_Elev_NMO at n = 77) produced degenerate Phase 2 validation and are dropped or annotated in Section S5.

#### Per-target winner summary

**Table S3.1.** Phase 2 100-seed validated configurations for the three primary targets. All three primaries selected rank_combine as their ensemble combiner. Module names follow the paper convention defined in Section S2.

Three observations from the primary ensembles:

1. **DS is the load-bearing family across all three primary targets.** DS3 anchors MS Subtype (DS-only plus clinical / genome-wide anchors); DS1 and DS1-nc anchor MS vs NMO; DS6 and DS8 anchor Early MS. DM modules add roughly equal per-module value on the two between-disease targets, and clinical modules (C-family) contribute a small consistent boost on most targets. MS Subtype is entirely DS-and-clinical-driven with no DM contribution.

2. **Non-coding variants earn their place in the ensembles.** DM-200(nc) is in both between-disease primary ensembles (MS vs NMO and Early MS), DS1-nc is in MS vs NMO, GW1-nc is in MS Subtype. A coding-only pipeline would miss roughly 1–2% of AUC on MS vs NMO and MS Subtype, providing the experimental basis for Section S11's argument that the non-coding transcriptome is diagnostically meaningful and that microarray-only prior work has a structural ceiling our total-RNA platform does not share.

3. **Ensemble size is target-appropriate rather than fixed.** MS Subtype reaches its plateau at 4 modules, MS vs NMO at 6, Early MS at 7. The Early MS ensemble is the largest because the task is signal-sparse, EDSS $\leq$ 2.0 vs. healthy control is the hardest molecular discrimination in the paper, early-stage disease has the least disease-specific transcriptomic variance to exploit, and combining more weakly-discriminative modules is how the classifier reaches the 74.2% operating point. For all three primaries, the greedy stopping criterion (no additional module improves 30-seed AUC by > 0.001) produced the reported module count without human intervention.

#### Secondary-target winners

For the 23 secondary targets, greedy was run identically (full 33-module pool × both combiners × 30 seeds × seeds 2000–2029) and the per-target winners are tabulated in Section S5. Several secondary targets converge to 1–3-module ensembles (e.g. MRI Locations $\geq$ 3 (MS) reduces to C1 alone, Oligoclonal Bands (NMO) to C1 alone, Clinician Signs > 3 (NMO) to C4 alone), reflecting that at small n and low prevalence the greedy procedure refuses to add modules whose mean improvement cannot clear the noise floor. Two secondary targets had degenerate Phase 2 validation and are annotated: OCB_NMO (n = 87, 100-seed mean @100% below chance at 0.40, reflecting the intrinsic difficulty of predicting serology from PBMC expression at this sample size) and IgG_Elev_NMO (n = 77, insufficient for a stable 100-seed abstention zone, validation dropped). The remaining 21 secondary targets validated cleanly and are the basis for the 26-target information-boundary chart in Figure 5a.

#### Pseudocode

```
INPUT:
 module_library M (33 modules), targets T (26 targets),
 combiners {rank_combine, conviction},
 screening seeds S_scr = [2000..2029], validation seeds S_val = [0..99]
OUTPUT:
 per-target winning configuration (combiner, module list) and validated AUCs.

FOR each target t in T:
 #   Phase 1 Pre-compute: cache predictions for every module × every seed
 FOR each seed s in S_scr:
 fit_train_val_splits(t, s)
```

```
FOR each module m in M:
preds[t][s][m] = (module_m_scores_on_train_fold,
module_m_scores_on_val_fold)

#   Phase 1 Greedy under each combiner
FOR each combiner c in {rank_combine, conviction}:
metric_key = 'auc100' if t == 'EarlyMS_vs_Control' else 'auc80'

# Best solo
solo_metric = {m: mean_over_S_scr(metric(ensemble_score([m], c, s)))
for m in M}
C[t][c] = {argmax solo_metric}
best_auc[t][c] = max solo_metric values

# Forward selection
WHILE True:
best_delta = 0; best_cand = None
FOR each candidate m not in C[t][c]:
new_auc = mean_over_S_scr(metric(ensemble_score(C[t][c]   {m}, c, s)))
IF new_auc - best_auc[t][c] > best_delta:
best_delta = new_auc - best_auc[t][c]
best_cand = m
IF best_delta < 0.001: BREAK
C[t][c] = C[t][c]   {best_cand}
best_auc[t][c] += best_delta

#   Select per-target winner combiner
c_winner = argmax over {rank_combine, conviction} of best_auc[t][c]
modules_winner = C[t][c_winner]

#   Phase 2 Validation on disjoint seed pool
val_metrics = []
FOR each seed s in S_val:
fit_train_val_splits(t, s)
scores_train_ens, scores_val_ens = ensemble_score(modules_winner, c_winner, s)
zone = abstention_zone(scores_train_ens, labels_train) # see §S1
auc100, auc80, coverage = apply_zone(zone, scores_val_ens, labels_val)
val_metrics.append((auc100, auc80, coverage))
RETURN modules_winner, c_winner, mean_and_sem(val_metrics)
```

#### S4. Classifier robustness: seed stability and validation shrinkage

This section quantifies two robustness properties of the reported mean AUCs: their per-seed stability across 100 Monte Carlo splits, and their shrinkage from the 30-seed configuration-selection phase to the 100-seed validation phase.

#### Methods

We employed a two-phase evaluation design to separate configuration search from performance estimation. In Phase 1, we evaluated all candidate module configurations at 30 random seeds to identify the best-performing combination per target. In Phase 2, we re-evaluated winning configurations at 100 seeds with per-seed gene selection excluding held-out data, producing the final reported AUCs. This design bounds the optimism inherent in configuration search: the 30-seed phase selects which configuration to report, while the 100-seed phase estimates how well that configuration actually performs. Shrinkage between phases quantifies the selection bias.

#### Results

Per-seed AUC distributions across the 100 validation seeds are tight for the primary targets and broaden for smaller-cohort secondary targets (Figure S4a). Inter-quartile ranges are 6-8% for the three paper-primary targets and widen to 10-15% for the smallest NMO-specific cohorts, as expected from the inverse dependence of SEM on sample size. Per-target means and SEMs are tabulated in Section S5.

The 30-seed screening AUC tracks the 100-seed validation AUC closely across all 26 targets at 80% coverage (Figure S4b). Median shrinkage from exploration to validation is ~2% for the primary targets and larger for the smallest-cohort NMO endpoints, reflecting both higher variance in the 30-seed estimate and greater sensitivity to specific training samples. Because screening uses seeds 2000-2029 and validation uses seeds 0-99, the two seed pools are disjoint and no selection-bias inflation is possible across phases.

#### Interpretation

The per-seed distributions are tight enough that reported mean AUCs reflect typical behavior of the chosen configuration rather than outlier seeds, and the screening-to-validation shrinkage of ~2% on the primary targets bounds the configuration-search optimism. Shrinkage scales inversely with sample size, consistent with sampling-noise rather than systematic overfitting as the dominant contributor. The configuration-search overstatement on the primary targets is therefore below 2% and is quantified directly for every target.

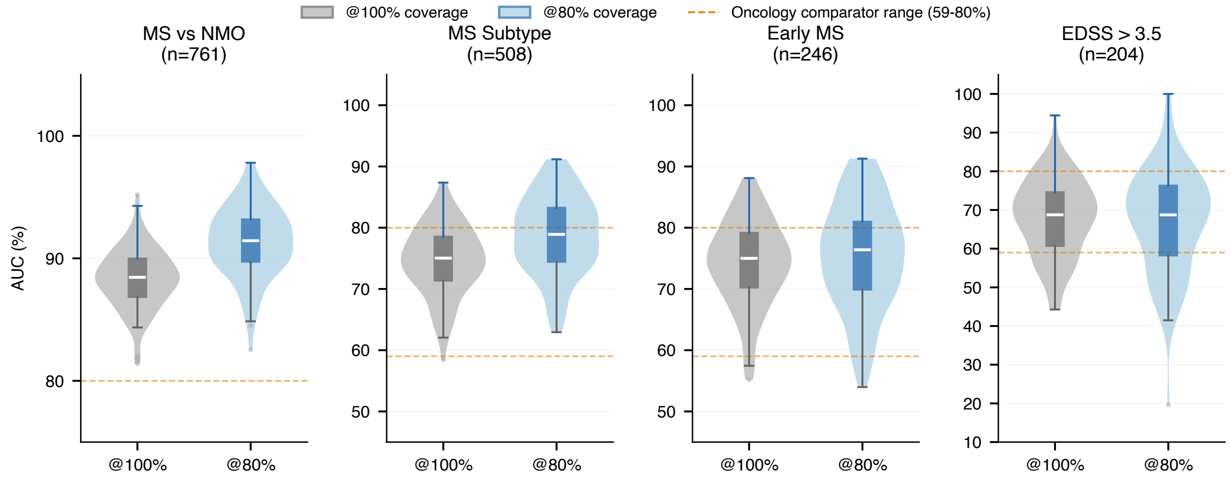

**Figure S4a.** Per-seed AUC violin distributions for primary targets.

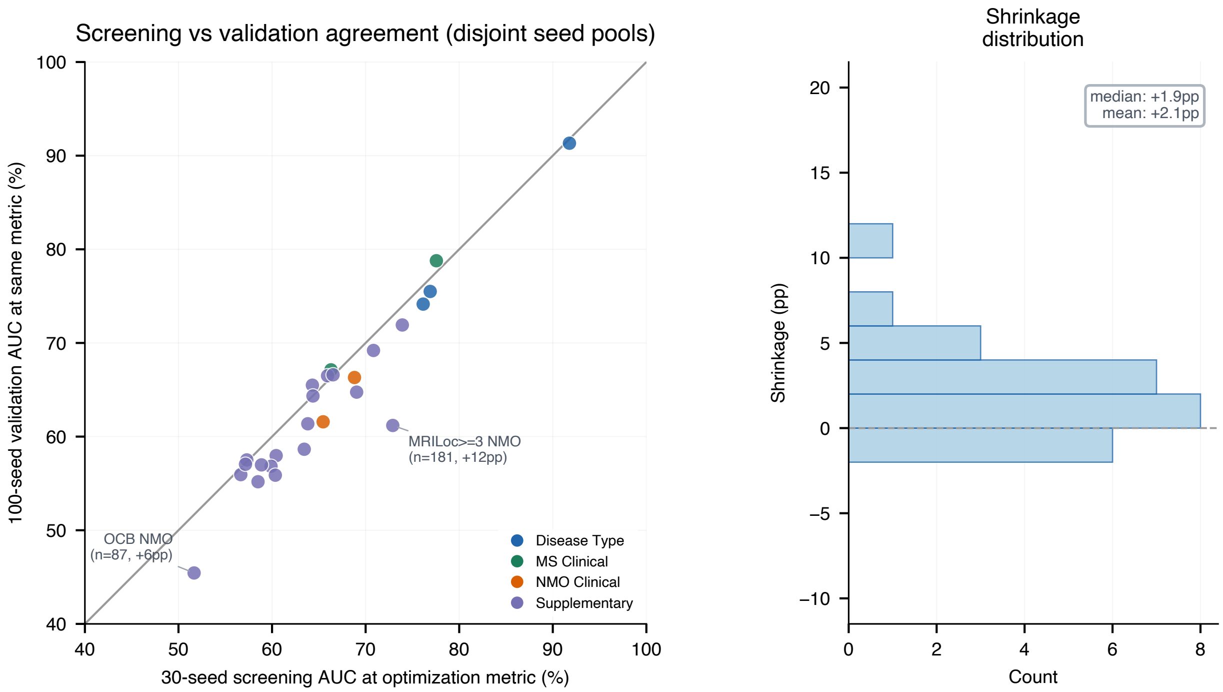

**Figure S4b.** 30-seed screening vs 100-seed validation scatter across all 26 targets.

### S5. The empirical information boundary: what blood can and cannot predict

We evaluated PBMC transcriptomics across 26 binary prediction targets spanning disease classification, disability, MRI imaging, CSF biomarkers, and temporal events (Table S5-1; visualized as a ranked AUC landscape in main paper Fig. 5a).

We evaluated each target under the full pipeline: greedy module selection at 30 seeds, followed by 100-seed validation of the best configuration. We report performance as mean AUC at 100% coverage (all patients classified) and 80% coverage (20% most uncertain abstained). All targets used the same module library (Section S2) and the same selection procedure (Section S3); no target received bespoke feature engineering.

The 26-target performance landscape (main paper Fig. 5a; numeric AUCs in Table S5-1 below) reveals a clear hierarchy. Disease classification targets dominate the top of the AUC distribution. Within-disease subtyping lands next, followed by disability staging across EDSS thresholds. Symptom- and sign-burden targets cluster in a mid-range band. MS MRI activity targets and CSF biomarker targets cluster near chance in both diseases; NMO MRI targets sit modestly above chance but below diagnostic-grade. Temporal targets (exacerbation status and EDSS progression) sit at chance.

We interpret this hierarchy as consistent with a biological boundary. Blood-based prediction succeeds on the targets reflective of stable peripheral immune state (disease identity, disease phase, cumulative disability burden) and fails on the targets that, on their face, would require either CNS-compartmentalized information (MS MRI lesions), temporally transient events (exacerbation timing), or longitudinal trajectory information (EDSS progression). We discuss two specific cases below.

The EDSS Progression greedy winner reaches 58.0% AUC at 80% coverage, indistinguishable from chance, despite running under the same greedy procedure as every other target and drawing from the same 33-module pool. The contrast with the static disability state target (EDSS >3.5, 67.1% AUC at 80%) on the same MS-only cohort (n = 204) is not attributable to sample size. Two interpretations are consistent with this gap. First, a single-timepoint PBMC transcriptome may capture the cumulative immune state associated with current disability level without containing the temporal-derivative information needed to infer future trajectory. Second, the trajectory phenotype itself may be more heterogeneous than the static phenotype, so the same data could in principle suffice for some patients but not others. The result is consistent with both readings; we cannot distinguish them with cross-sectional data alone.

The MRI asymmetry between NMO and MS is consistent with a compartmentalization explanation. NMO pathology is driven in part by peripheral antibodies (AQP4-IgG) whose effector cells circulate in blood, while MS lesion activity is driven by CNS-resident processes that may be largely invisible to peripheral sampling. The classifier ranks NMO MRI targets above MS MRI targets without prior specification of this biology, which we read as suggestive (though not proof) that the predictive signal aligns with the underlying compartment differences rather than with shared confounds.

The DM + DS combined search assembles each primary ensemble from a target-appropriate mixture of both families plus a small clinical / compositional anchor. MS vs. NMO takes DM-200 + DM-200(nc) + C2 + DS1 + C1 + DS1-nc: two DM variants and two DS variants span the coding / non-coding space, with C2 (hierarchical immune composition) and C1 (demographics) contributing orthogonal baseline signal, consistent with mean B-cell and immunoglobulin expression carrying a dominant share of the between-disease signal. Early MS vs. Control takes DM-50 + DS8 + DM-200 + C3 + DM-200(nc) + DS6 + PCA-64: three DM variants pair with two DS modules, one clinical module (C3, ancestry / HLA), and the unsupervised PCA-64 baseline, consistent with a broad and multi-modal population separation. MS Subtype takes DS3 + C1 + C4 + GW1-nc: DS and clinical modules suffice, and no DM module survives greedy, consistent with a within-disease contrast where the biological signal is variability-shift rather than mean-shift.

#### Table S5-1. Prediction target definitions and classifier performance for all 26 targets

Each row defines one binary prediction target evaluated by the ACP PBMC modular ensemble classifier. Targets are grouped by category: Disease Classification (differential diagnosis and case-control), MS Clinical (primary MS severity measures), NMO Clinical (primary NMO measures), and Supplementary (additional clinical endpoints). Stars ( ) mark the three paper-primary targets. Positive class prevalence is the fraction of eligible samples labeled positive. AUC values are mean ± SEM across 100 random train/test splits. AUC @80% denotes performance on the 80% of samples with highest classifier confidence (20% abstention; see Section S1). Module configurations use paper names defined in Table S2-1 (module catalog), which includes the DM family (DM-50, DM-200, DM-200(pc), DM-200(nc), plus two PCA variants) and the unsupervised PCA-64 baseline alongside the Disease-Sigma (DS) and clinical/genome-wide families. The "Filter" column denotes which samples enter the analysis (MS-only, NMO-only, MS+Control, MS+NMO, EarlyMS+Control).

Legend for ensemble combiner column: **Rank** = rank_combine (per-module rank vs. training-fold reference; mean of ranks). **Conv** = conviction-weighted mean (weight `w_i = max(-log (p), -log (1-p))`; weighted mean of raw probabilities). See Section S3 for the ensemble-combiner comparison and per-target selection rationale. `--` under AUC @80% means the metric is not reported for that target (screening-class Early MS) or was degenerate at validation (IgG Index NMO, n=77 with unstable ROC).

**Disease Classification**

**MS Clinical**

**NMO Clinical**

| Target | Combiner | Filter | n | Prev | Modules | @100% | @80% | Cov |
|---|---|---|---|---|---|---|---|---|
| ★ MS vs NMO | Rank | MS+NMO | 761 | 0.29 | DM-200 + DM-200(nc) + C2 + DS1 + C1 + DS1-nc | 0.883 ± 0.002 | 0.913 ± 0.003 | 0.75 |
| MS vs Healthy Control | Rank | MS+Control | 689 | 0.78 | DS5 + DM-200(nc) + DM-200(pc) + C3 + DS1 | 0.727 ± 0.005 | 0.755 ± 0.006 | 0.75 |
| ★ Early MS vs Control | Rank | EarlyMS+Ctrl | 246 | 0.39 | DM-50 + DS8 + DM-200 + C3 + DM-200(nc) + DS6 + PCA-64 | 0.742 ± 0.007 | – (screening; §S1) | 0.72 |

| Target | Combiner | Filter | n | Prev | Modules | @100% | @80% | Cov |
|---|---|---|---|---|---|---|---|---|
| ★ MS Subtype (progressive vs relapsing) | Rank | MS-only | 508 | 0.15 | DS3 + C1 + C4 + GW1-nc | 0.747 ± 0.006 | 0.788 ± 0.006 | 0.77 |
| EDSS Disability > 3.5 | Rank | MS-only | 204 | 0.25 | C4 + C1 + DS7 | 0.679 ± 0.010 | 0.671 ± 0.013 | 0.78 |

| Target | Combiner | Filter | n | Prev | Modules | @100% | @80% | Cov |
|---|---|---|---|---|---|---|---|---|
| T2 Lesion Burden > 7 (NMO) | Rank | NMO-only | 171 | 0.16 | C4 + DS7-nc + PCA-64 | 0.650 ± 0.012 | 0.663 ± 0.015 | 0.73 |
| New MRI Lesions (NMO) | Rank | NMO-only | 89 | 0.61 | DM-200(nc) + DS2 | 0.573 ± 0.015 | 0.616 ± 0.027 | 0.46 |

| Target | Combiner | Filter | n | Prev | Modules | @100% | @80% | Cov |
|---|---|---|---|---|---|---|---|---|
| EDSS > 2 (MS) | Conv | MS-only | 204 | 0.53 | DM-200(pc) + C1 + C3 + C4 + DS4-nc | 0.617 ± 0.008 | 0.655 ± 0.012 | 0.56 |
| EDSS > 5 (MS) | Rank | MS-only | 204 | 0.19 | C1 + DS4 | 0.659 ± 0.011 | 0.665 ± 0.015 | 0.79 |
| EDSS Progression (MS) | Rank | MS-only | 204 | 0.43 | DS6 + DM-200(nc) + C1 + DM-50 + DS7-nc | 0.567 ± 0.009 | 0.580 ± 0.010 | 0.73 |
| Walk 25ft > 7s (MS) | Rank | MS-only | 292 | 0.16 | DS3 + DS4 + DM-200 | 0.627 ± 0.013 | 0.648 ± 0.015 | 0.79 |

| Target | Combiner | Filter | n | Prev | Modules | @100% | @80% | Cov |
|---|---|---|---|---|---|---|---|---|
| Symptom Burden > 10 (MS) | Rank | MS-only | 534 | 0.47 | C1 + DS3-nc + DS1 + DM-200(pc) | 0.640 ± 0.005 | 0.666 ± 0.006 | 0.68 |
| Clinician Signs > 3 (MS) | Rank | MS-only | 530 | 0.51 | DM-200(nc) + C1 + DM-200 + GW1-nc + DS2-nc | 0.669 ± 0.005 | 0.719 ± 0.006 | 0.58 |
| In Exacerbation (MS) | Conv | MS-only | 472 | 0.11 | DS3 + C3 | 0.551 ± 0.008 | 0.552 ± 0.011 | 0.80 |

#### Supplementary, MS Disability and Progression

#### Supplementary, MS Symptom and Sign Burden

#### Supplementary, MS MRI and CSF

#### Supplementary, NMO Additional Endpoints

| Target | Combiner | Filter | n | Prev | Modules | @100% | @80% | Cov |
|---|---|---|---|---|---|---|---|---|
| New MRI Lesions (MS) | Rank | MS-only | 388 | 0.55 | DS7 + C1 + DS3 + C3 + DS5 | 0.576 ± 0.006 | 0.587 ± 0.006 | 0.77 |
| T2 Lesion Burden > 7 (MS) | Conv | MS-only | 502 | 0.63 | DS3-nc + C4 + C3 | 0.562 ± 0.006 | 0.568 ± 0.006 | 0.80 |
| T2 Lesion Burden > 15 (MS) | Rank | MS-only | 502 | 0.31 | DS7-nc + DS2-nc + DS3-nc | 0.548 ± 0.005 | 0.559 ± 0.006 | 0.77 |
| MRI Locations ≥ 3 (MS) | Conv | MS-only | 530 | 0.24 | C1 | 0.558 ± 0.006 | 0.571 ± 0.007 | 0.79 |
| Oligoclonal Bands (MS) | Rank | MS-only | 202 | 0.70 | C3 + P2-nc | 0.559 ± 0.008 | 0.559 ± 0.010 | 0.78 |
| IgG Index Elevated (MS) | Conv | MS-only | 169 | 0.59 | DS5-nc + C3 + DS5 | 0.562 ± 0.011 | 0.575 ± 0.012 | 0.81 |

| Target | Combiner | Filter | n | Prev | Modules | @100% | @80% | Cov |
|---|---|---|---|---|---|---|---|---|
| T2 Lesion Burden > 3 (NMO) | Rank | NMO-only | 171 | 0.42 | C4 + DM-200 | 0.592 ± 0.009 | 0.614 ± 0.011 | 0.80 |
| Symptom Burden > 10 (NMO) | Rank | NMO-only | 203 | 0.43 | C3 + C2 | 0.682 ± 0.007 | 0.692 ± 0.009 | 0.81 |
| Clinician Signs > 3 (NMO) | Conv | NMO-only | 181 | 0.33 | C4 | 0.624 ± 0.010 | 0.643 ± 0.011 | 0.83 |
| MRI Locations ≥ 3 (NMO) | Conv | NMO-only | 181 | 0.17 | DM-50 + DS2 + GW1-nc + DS1 | 0.546 ± 0.012 | 0.612 ± 0.018 | 0.53 |
| Oligoclonal Bands (NMO) | Conv | NMO-only | 87 | 0.30 | C1 | 0.401 ± 0.018 | 0.454 ± 0.026 | 0.63 |
| IgG Index Elevated (NMO) | Rank | NMO-only | 77 | 0.27 | C3 + PCA-64 + C1 | – | – | – |

#### S6. Treatment confounding: disease biology vs. drug effect

A substantial fraction of the cohort receives disease-modifying therapy, most commonly rituximab. Because treatment correlates with disease (rituximab is first-line for NMO), classifiers may detect drug-associated transcriptomic changes rather than disease biology. We tested this by restricting the cohort to rituximab-naive patients and retraining.

For each primary target, we repeated the cross-validation pipeline on the rituximab-naive subset only (30 seeds per stratum). All other pipeline parameters (module configurations, feature selection, classifier architecture) remained identical. The configurations evaluated here are an earlier ensemble version that predates the DM family expansion; the qualitative pattern is expected to carry over to the current DM+DS ensembles since the underlying B-cell biology is unchanged.

The all-sample vs rituximab-naive comparison shows target-dependent treatment effects (Figure S6). MS vs NMO drops from 94.1% to 89.1% (−5.0%); MS Subtype drops from 78.5% to 77.3% (−1.2%); MS vs Control changes from 77.2% to 77.4% (+0.2%, effectively unchanged).

The three primary classifiers are largely robust to treatment stratification. We hypothesize that the 5.0% reduction for MS vs NMO reflects rituximab's depletion of CD20+ B cells, which carry a portion of the NMO-specific transcriptomic signal; the classifier still reaches 89.1% AUC in rituximab-naive patients, which we read as consistent with disease biology rather than drug effect carrying the majority of the discriminative signal on this target. The near-zero change for MS vs Control is consistent with the classifier indexing disease-intrinsic immune changes rather than pharmacological perturbation, though we cannot rule out compensating shifts that happen to cancel.

We separately evaluated a broad-spectrum drug-covariate correction in which an 8-covariate model (drug indicators for all major disease-modifying therapies) was regressed out of the expression matrix prior to module construction. The results were target-dependent: drug-covariate correction reduced MS Subtype AUC by 6.3% (from 0.636 to 0.573), consistent with removal of treatment-correlated variance that is itself informative for distinguishing progressive from relapsing disease, but improved EDSS >3.5 prediction by 12.8% (from 0.528 to 0.656), indicating that drug-associated transcriptomic changes were masking the disability signal in uncorrected data.

Treatment confounding does not act uniformly across prediction tasks: universal drug correction would improve some targets at the cost of degrading others. We therefore did not apply drug correction globally and relied on the cross-validation framework to assess whether drug-associated variance helps or hinders each specific task. The cohort's pharmacological complexity reinforces the choice: interferon-beta upregulates interferon-stimulated genes (a potential confound for immune-activation signatures), while glatiramer acetate has widespread immunomodulatory effects on monocyte and T-cell compartments that overlap with disease-relevant biology.

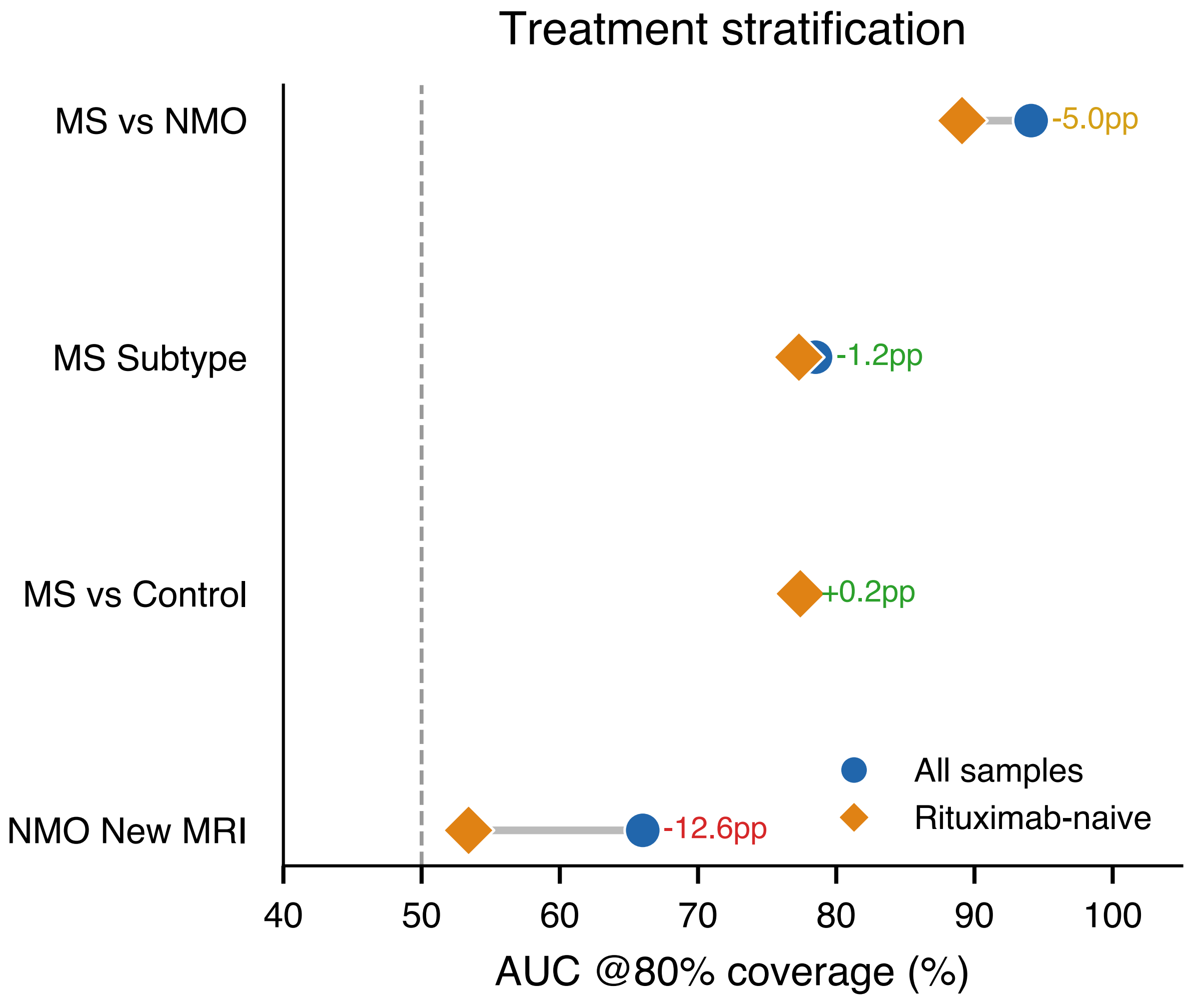

**Figure S6.** All-sample vs rituximab-naive AUC for primary targets and NMO new MRI.

#### S7. Site confounding: hold-out validation rules out batch effects

The cohort was processed across multiple sites, and processing site is partially confounded with disease identity. Site 82 contributes 71% of all NMO samples (156 of 221), while Site 1 contributes 42% of all MS samples (226 of 540). A classifier exploiting site-specific expression profiles rather than disease biology could achieve high cross-validation AUC through batch effects alone. We addressed this with two complementary hold-out designs that neutralize site identity as a discriminative cue.

The two designs target different failure modes of site confounding. Hold-out A is a same-site balanced test: we selected 36 MS and 36 NMO samples from Site 82, subsampling the 156 Site-82 NMO down to 36 with a fixed seed to match the 36 MS available at that site. Training used every other MS and NMO sample in the cohort (504 MS + 185 NMO = 689). Because both test classes come from the same processing site, site identity carries zero information about the label within the test set; any batch effect is common to both classes and cannot contribute to discrimination. Hold-out B is a full site removal: we held out all 50 MS and 44 NMO samples from Site 78 and trained on the remainder (490 MS + 177 NMO = 667). This tests generalization to a site whose batch characteristics the model has never seen. In both designs, gene selection and quality-covariate correction were recomputed after excluding the test samples, and we evaluated over 100 random seeds that vary the internal train / test split used by the gene-consistency filter.

Both hold-outs were evaluated at 100 seeds (Figure S7). Under the same-site-balanced design (Hold-out A), MS vs. NMO classification remains within 5% of the all-sites cross-validation reference at both 100% and 80% coverage, despite site identity carrying zero information about the label within the test set. Under the full-site-removal design (Hold-out B), classification on samples drawn from a never-seen site likewise remains within 5% of the reference. The companion Site-78 dropout for MS Subtyping (RRMS vs progressive) lands ~9% *above* its cross-validation reference, consistent with the Site-78 progressive samples falling within the distribution of the remaining-site training population (main paper Fig. 4b).

The same-site balanced design is the tighter test of batch-effect dependence: if the classifier were separating MS from NMO by detecting Site 82's processing signature, performance on Hold-out A would collapse to chance because every test sample carries that same signature. The observed AUC is inconsistent with that mechanism. Hold-out B adds the complementary check on never-seen-site generalization; the ~4-5% reduction from the cross-validation reference is consistent with a smaller, less diverse training pool rather than with batch-effect loss. A Phase 2 multi-site validation study (~2,000 additional samples from independent collaborators) is in progress on a cohort where site and disease are not confounded by design.

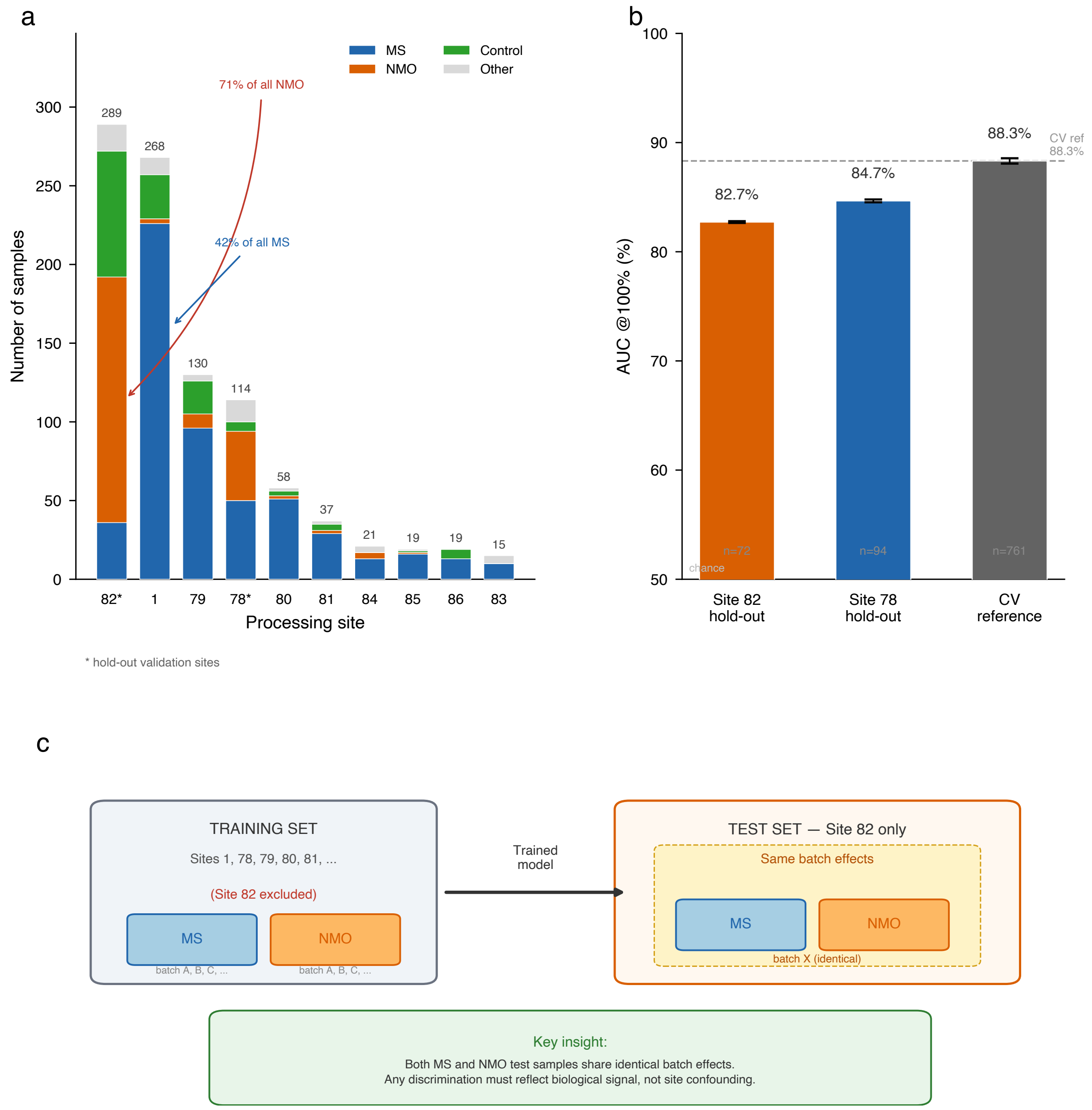

**Figure S7.** Site hold-out validation: per-seed distributions vs CV reference.

#### S8. EDSS >3.5 disability classifier

Disability stratification is among the most clinically valuable targets for MS diagnostics, as EDSS score drives treatment escalation decisions. We evaluated an EDSS >3.5 classifier on 204 MS patients with available EDSS data, using a greedy-selected module configuration validated at 100 seeds.

The EDSS >3.5 classifier reaches AUC in the high-60s at both 100% and 80% coverage (Figure S8), trailing the three primary paper targets and sitting within the band of disability-related secondary endpoints tabulated in Section S5. Two related disability targets, EDSS >5 and timed 25-foot walk >7 seconds, reach similar AUCs (main paper Fig. 5a,b).

Two interpretations are consistent with the near-equality of @100% and @80% AUC on this target. Either the training-derived abstention zone fails to cleanly separate decisive from near-boundary validation samples, or the underlying per-sample signal is weak enough that no abstention zone could sharpen retained-set AUC. Either reading argues against promoting EDSS >3.5 to a primary finding at the present cohort size.

Three considerations bear on the interpretation. First, n = 204 is the smallest primary-candidate cohort, limiting statistical power and increasing overfit risk despite the 100-seed validation. Second, EDSS is subject to well-documented inter-rater variability, a noise source that inflates the irreducible error floor and that larger cohorts can partially average out. Third, the prospective Phase 2 expansion (~2,000 additional samples) will allow tighter characterization of the disability classifier than the present cohort supports.

The greedy-selected ensemble (C4 immune-diversity + C1 demographics + DS7 compact MS-stratified panel; Section S2 for module details) combines compositional and variability-shift signal. The MS-stratified DS panel is the strongest compact variance-shift contributor on this target; the cleaner cross-disease B-cell-transfer signal is reported on the MS Subtype ensemble in Section S9.

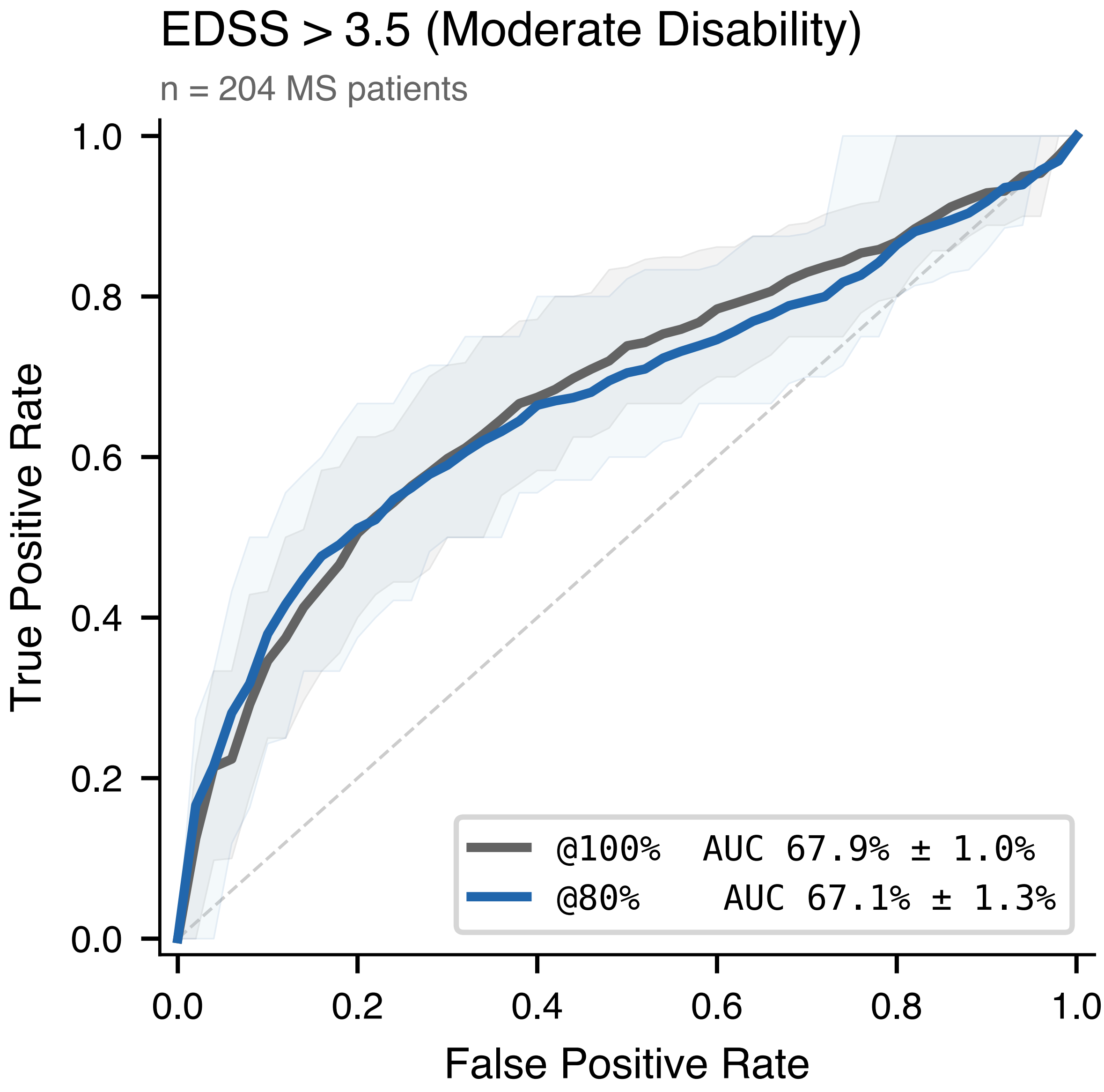

**Figure S8.** ROC curves for EDSS > 3.5 disability classifier (Phase 2).

#### S9. Cross-disease predictive transfer

If the Disease-Sigma modules were overfit to their training distribution (learning label-associated artifacts rather than biology), they would improve prediction only for targets within their source disease. An overfit MS module would help predict MS targets but not NMO targets, and vice versa. We examined greedy forward selection results for cross-disease module usage: cases where the optimal configuration for a target in one disease includes a module derived from the other disease.

The transfer is bidirectional. NMO-derived B-cell features enter the optimal configuration for MS targets in multiple places. The clearest instance is the MS Subtype primary ensemble, where a compact 50-gene panel selected on NMO-within-class variability anchors the progressive-vs-relapsing MS classifier. Early

MS vs Control similarly recruits an NMO-derived module: the winning ensemble includes a top-500 NMO-stratified protein-coding panel alongside an MS-derived Disease-Sigma module and four non-DS modules. In the opposite direction, an MS-derived non-coding compact panel enters the NMO T2 lesion burden >7 winning ensemble alongside an immune-diversity module and an unsupervised PCA baseline. Module ablation confirms that removing the cross-disease module from each of these configurations reduces AUC; greedy forward selection, not post-hoc curation, surfaced every transfer.

We hypothesized that shared immune mechanisms operating beneath disease-specific signatures ground the bidirectional transfer. Two lines of prior evidence are consistent with this reading. First, B-cell involvement in MS extends beyond its primary role in NMO: meningeal B-cell follicles, intrathecal immunoglobulin production, and the therapeutic efficacy of anti-CD20 agents in progressive MS all indicate that B-cell biology contributes to MS pathology [21, 5]. NMO-derived Disease-Sigma modules may therefore capture B-cell signal in MS because PBMCs sample that compartment in both diseases, directly in NMO (via pathogenic AQP4-IgG-producing cells) and indirectly in MS (via B-cell-mediated neurodegeneration and progressive-phase meningeal-follicle biology). Second, innate immune activation extends beyond MS: monocyte and macrophage infiltration amplifies antibody-mediated damage in NMO, which would explain why MS-derived modules contribute to NMO imaging targets. An alternative reading, that the cross-transfer reflects shared technical variance rather than shared biology, is harder to reconcile with the gene-level analysis below.

Gene-level orthogonality between selection approaches is consistent with the biological interpretation. The Disease-Sigma gene lists (top 200 each in MS and NMO) share zero genes with a pooled approach that computes interquartile range across the mixed MS+NMO cohort. The pooled approach selects genes that vary across the combined cohort (a signal dominated by between-disease differences); the Disease-Sigma approach selects genes that vary within each disease group, accessing disease-specific biological programs. The two Disease-Sigma lists themselves share only 42 of 200 genes (21%), with the MS list enriched for monocyte/macrophage markers (SDC2, PPARG, IL1R1, IL1B) and the NMO list for B-cell and immunoglobulin genes (MS4A1, CD79A, IGHD, PAX5). Each of the three selection axes accesses a distinct slice of transcriptomic variation; the cross-disease transfer is observed only on the Disease-Sigma axes.

The bidirectional transfer is consistent with the compartmentalization argument in Figure 4: the same immune compartments (B-cell, myeloid) that define orthogonal disease-specific programs also contribute, at lower magnitude, across disease boundaries. Transfer schematics and ablation results are in Supplementary Figure S9.

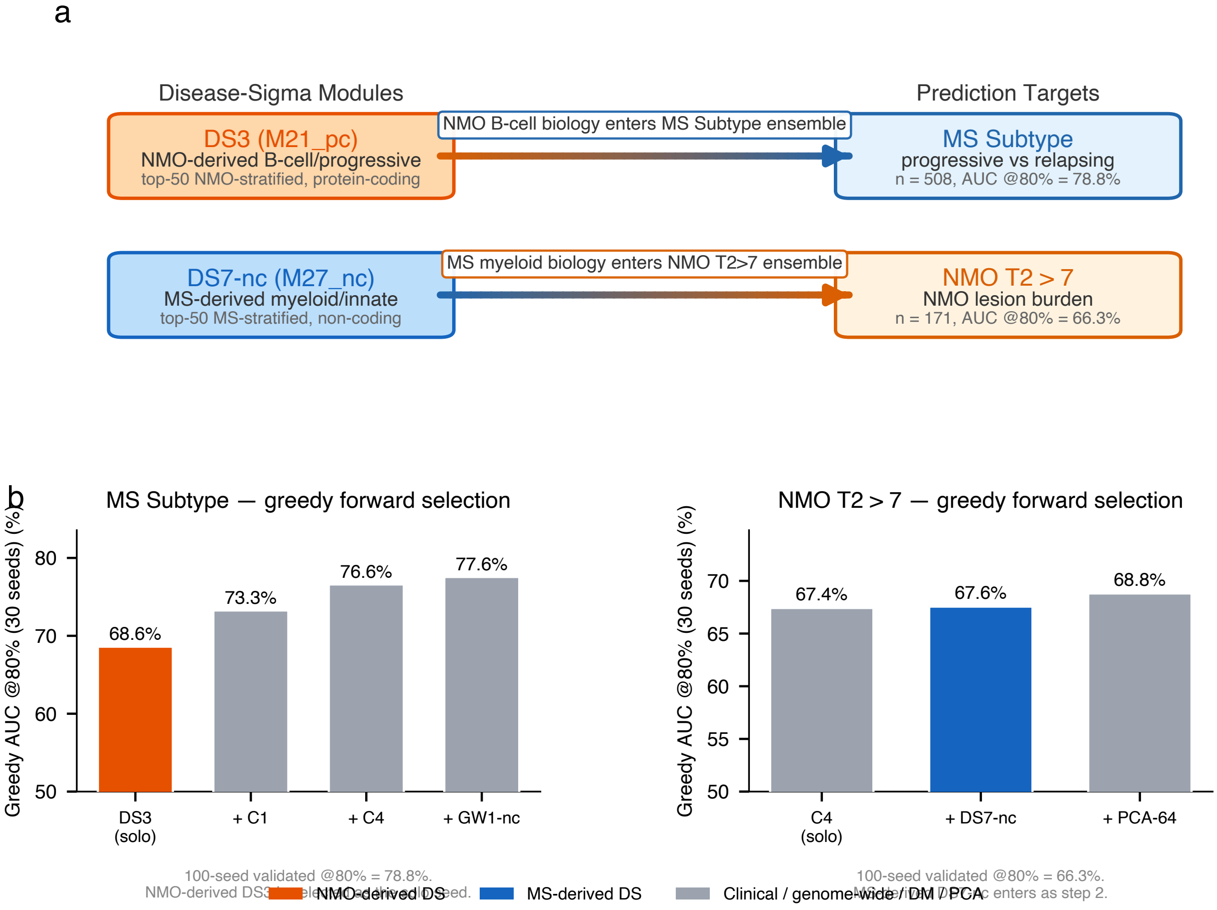

**Figure S9.** Bidirectional cross-disease module transfer.

#### S10. Biological validation: gene ontology enrichment

The Disease-Sigma gene selection procedure ranks genes by within-disease variability against internal healthy controls, using no disease labels and no supervised information. If the resulting gene lists reflected technical artifacts (batch effects, processing site variation, or stochastic noise), the top-ranked genes would be biologically incoherent. We tested biological coherence by submitting the top 200 genes from each Disease-Sigma signature to gene ontology enrichment analysis using g:Profiler with FDR correction across GO Biological Process, GO Molecular Function, GO Cellular Component, KEGG, and Reactome databases.

The DS_MS signature (170 of 200 genes recognized; 30 lncRNA and pseudogene identifiers absent from reference databases) recovers a myeloid and innate immune program consistent with MS pathophysiology. The dominant enrichments are cell activation ($p = 1.4 \times 10^{-13}$), immune system process ($p = 1.8 \times 10^{-12}$), and cell surface receptor signaling ($p = 2.5 \times 10^{-11}$). We also observe a blood coagulation and platelet activation signature ($p = 8.9 \times 10^{-9}$ and $1.5 \times 10^{-7}$, respectively), consistent with platelet-derived RNA in PBMC preparations and with platelet-mediated neuroinflammation in MS. Key protein-coding genes include SDC2 (macrophage activation marker), PPARG (macrophage polarization regulator), IL1R1 and IL1B (interleukin-1 axis), MMP25 (matrix metalloproteinase), and IFI44 (interferon-induced).

The DS_NMO signature (160 of 200 genes recognized) recovers a B-cell and humoral immune program that directly reflects NMO's antibody-mediated pathogenesis. The dominant enrichments are B cell receptor signaling ($p = 9.4 \times 10^{-12}$), IgG immunoglobulin complex ($p = 2.0 \times 10^{-9}$), and B cell activation ($p = 2.7 \times 10^{-9}$). Key genes include MS4A1 (CD20, the therapeutic target of anti-CD20 agents), CD79A (B-cell receptor component), PAX5 (B-cell lineage commitment factor), immunoglobulin heavy and light chain genes (IGHD, IGHG1-3), and B-cell signaling genes EBF1, BLK, and POU2AF1.

These gene sets were selected by unsupervised variability ranking, and recover the same B-cell and myeloid programs that prior hypothesis-driven immunology has independently identified in NMO and MS. The recovered enrichments span ~12 orders of magnitude in p-value on disease-relevant pathways, which is harder to reconcile with technical artifact (batch, site, processing noise) than with disease-specific biological signal. The biological coherence complements the external checks from site hold-out analysis (Supplementary Section S7) and treatment stratification (Figure S6). Full enrichment tables are in Supplementary Figure S10.

a

DS_MS top 200 genes: Myeloid / innate immune program

Disease-stratified variability ranking recovers cell activation, coagulation, and cytokine signaling pathways

| # | Enriched Term | Cat. | $-\log_{10}(p)$ | FDR p-value | Genes | Key Genes |
|---|---|---|---|---|---|---|
| 1 | Cell periphery | CC | | 3.9 x 10^-16 | 104 | SDC2, MMP25, ITGB3, CD33, SIGLEC9 |
| 2 | Cell activation | BP | | 1.4 x 10^-13 | 41 | IL1B, SELP, CLEC7A, BST1, ITGB3 |
| 3 | Plasma membrane | CC | | 1.1 x 10^-12 | 93 | IL1R1, SDC2, CD33, MSR1, C5AR1 |
| 4 | Immune system process | BP | | 1.8 x 10^-12 | 63 | IL1R1, PPARG, CXCL8, IL1B, IFI44 |
| 5 | Multicellular organismal process | BP | | 2.0 x 10^-12 | 106 | IL1B, PPARG, CXCL8, SPARC, MMP25 |
| 6 | Cell surface receptor signaling | BP | | 2.5 x 10^-11 | 60 | IL1R1, IL1B, CXCL8, CCL2, C5AR1 |
| 7 | Regulation of multicellular organismal process | BP | | 4.5 x 10^-11 | 61 | IL1B, PPARG, CXCL8, THBS1, CCL2 |
| 8 | Regulation of immune system process | BP | | 8.7 x 10^-11 | 43 | PPARG, IL1B, CXCL8, CLU, VSIR |
| 9 | Positive regulation of multicellular organismal process | BP | | 1.0 x 10^-10 | 44 | IL1B, PPARG, CXCL8, BST1, THBS1 |
| 10 | Regulation of response to stimulus | BP | | 1.5 x 10^-10 | 71 | IL1R1, IL1B, PPARG, CXCL8, CCL2 |
| 11 | Positive regulation of immune system process | BP | | 2.1 x 10^-10 | 35 | IL1B, CXCL8, CCL2, THBS1, BST1 |
| 12 | Response to stimulus | BP | | 3.5 x 10^-9 | 112 | IFI44L, IFI44, IL1B, PPARG, CXCL8 |
| 13 | Hemostasis (Reactome) | Reactome | | 3.9 x 10^-9 | 31 | SDC2, SELP, ITGB3, THBS1, GP1BB |
| 14 | Immune response | BP | | 6.3 x 10^-9 | 46 | IFI44L, CXCL8, CCL2, MX1, OAS3 |
| 15 | Wound healing | BP | | 6.3 x 10^-9 | 21 | SELP, PPARG, CLEC7A, ITGB3, HBEGF |
| 16 | Blood coagulation | BP | | 8.9 x 10^-9 | 16 | SELP, ITGB3, F13A1, THBD, GP1BB |
| 17 | Regulation of cell activation | BP | | 2.5 x 10^-8 | 24 | IL1B, ICOS, BST1, VSIR, CD33 |
| 18 | Platelet activation | BP | | 1.5 x 10^-7 | 12 | SELP, P2RY12, ALOX12, ITGB3, GP1BB |
| 19 | Extracellular space | CC | | 1.5 x 10^-7 | 57 | IL1B, CXCL8, CCL2, MMP25, SPON2 |
| 20 | Inflammatory response | BP | | 1.2 x 10^-6 | 25 | IL1B, IL1R1, CXCL8, PTGS2, CLEC7A |

g:Profiler enrichment analysis | 170 of 200 genes recognized (30 lncRNA/pseudogene excluded from reference databases) | FDR-corrected p-values

GO Biological Process GO Molecular Function GO Cellular Component Reactome KEGG

b

DS_NMO top 200 genes: B-cell / humoral immune program

Disease-stratified variability ranking recovers B cell receptor signaling, immunoglobulin complexes, and adaptive immunity

| # | Enriched Term | Cat. | $-\log_{10}(p)$ | FDR p-value | Genes | Key Genes |
|---|---|---|---|---|---|---|
| 1 | Leukocyte activation | BP | | 5.2 x 10^-12 | 36 | CD79A, PAX5, MS4A1, POU2AF1, CD38 |
| 2 | Immune response | BP | | 9.4 x 10^-12 | 51 | CD79A, BLK, IGHG1, IGHG3, IGHM |
| 3 | B cell receptor signaling pathway | BP | | 9.4 x 10^-12 | 13 | CD79A, BLK, MS4A1, BANK1, CD38 |
| 4 | Lymphocyte activation | BP | | 6.2 x 10^-11 | 31 | PAX5, MS4A1, POU2AF1, CD79A, CXCR5 |
| 5 | Immune system process | BP | | 6.7 x 10^-11 | 59 | CD79A, BLK, MS4A1, IGHG1, POU2AF1 |
| 6 | Cell activation | BP | | 6.9 x 10^-11 | 36 | PAX5, MS4A1, POU2AF1, CD79A, CD38 |
| 7 | IgG immunoglobulin complex | CC | | 2.0 x 10^-9 | 6 | IGHG1, IGHG2, IGHA1, IGLC2, IGLC3 |
| 8 | B cell activation | BP | | 2.6 x 10^-9 | 18 | MS4A1, PAX5, POU2AF1, CD79A, BANK1 |
| 9 | Immune effector process | BP | | 3.1 x 10^-9 | 27 | POU2AF1, IGHG1, IGHG2, IGHM, SWAP70 |
| 10 | Leukocyte mediated immunity | BP | | 4.7 x 10^-9 | 22 | IGHG1, IGHG2, IGHM, TBX21, NCR1 |
| 11 | Adaptive immune response | BP | | 6.2 x 10^-9 | 27 | CD79A, IGHG1, IGHG3, IGHM, JCHAIN |
| 12 | Antigen binding | MF | | 10.0 x 10^-9 | 15 | IGHG1, IGHG2, IGHM, JCHAIN, IGHA2 |
| 13 | Immunoglobulin complex, circulating | CC | | 1.6 x 10^-8 | 6 | JCHAIN, IGHA2, IGHG2, IGHA1, IGHM |
| 14 | Immunoglobulin receptor binding | MF | | 1.9 x 10^-8 | 7 | JCHAIN, IGHA2, IGHG2, IGHA1, IGHM |
| 15 | Lymphocyte mediated immunity | BP | | 2.4 x 10^-8 | 19 | IGHG1, IGHG2, IGHM, TBX21, SWAP70 |
| 16 | Antigen receptor-mediated signaling | BP | | 6.6 x 10^-8 | 14 | CD79A, BLK, MS4A1, BANK1, CD38 |
| 17 | Immunoglobulin mediated immune response | BP | | 1.5 x 10^-7 | 14 | IGHG1, IGHG2, IGHM, SWAP70, FCER2 |
| 18 | B cell mediated immunity | BP | | 1.6 x 10^-7 | 14 | IGHG1, IGHG2, IGHM, SWAP70, CD40 |
| 19 | Immunoglobulin complex | CC | | 1.0 x 10^-6 | 12 | IGHG1, IGHG3, IGHD, IGHM, CD79A |
| 20 | Hematopoietic cell lineage | KEGG | | see text | - | MS4A1, CD79A, CD38, CD40, IL2RB |

g:Profiler enrichment analysis | 160 of 200 genes recognized (40 lncRNA/pseudogene excluded from reference databases) | FDR-corrected p-values

GO Biological Process GO Molecular Function GO Cellular Component Reactome KEGG

**Figure S10.** Full gene ontology enrichment for DS_MS and DS_NMO top 200 genes.

### S11. Non-coding RNA: platform contribution to disease signatures

Standard mRNA-seq protocols and microarray platforms restrict analysis to protein-coding genes, which constitute approximately 40% of the viable feature space on the BIRT platform. We asked whether BIRT's expanded coverage of non-coding RNA species (lncRNA, pseudogene transcripts, miRNA, snoRNA, and snRNA) contributes to the Disease-Sigma signatures.

Biotype classification of the Disease-Sigma gene lists (GENCODE annotations) shows an asymmetry between targets. The MS-specific signature (DS_MS top 200) is 68% non-coding RNA: 41% lncRNA, 14% pseudogene, 5% miRNA, and smaller contributions from snoRNA and snRNA, a modest enrichment over the 60% non-coding base rate in the viable gene pool (46,722 genes with variance > 0.01). The NMO-specific signature is 42% non-coding, closer to background; one possible reason is that the B-cell program is dominated by protein-coding immunoglobulin genes. The top-ranked gene in the MS-specific signature is IL6-AS1, an antisense lncRNA to IL-6, a known mediator of innate immune activation and neuroinflammation in MS. Other high-ranking MS lncRNAs (NUP133-DT, C1orf21-DT, and multiple unannotated ENSG identifiers) have no prior disease association in the literature we surveyed and are candidates for further functional characterization.

We quantified the functional contribution through a three-way comparison using greedy forward selection: mRNA-only (protein-coding gene pool), ncRNA-only (non-coding gene pool), and full model (all gene types). For MS vs NMO, ncRNA-only reaches 86.1% AUC and mRNA-only 85.7% in the ablation analysis (table below); the full ensemble's headline numbers are 88.3% / 91.3% at 100% / 80% coverage (§S5). For between-disease classification the non-coding transcriptome carries signal of comparable magnitude to the protein-coding transcriptome. One interpretation is that the structurally symmetric modules (protein-coding DS modules versus their non-coding counterparts) access overlapping myeloid and B-cell programs through independent molecular features; an alternative is that the two pools index partially distinct biology that happens to be similarly diagnostic. Consistent with the first reading, the MS vs NMO winning ensemble allocates two of its six modules to non-coding-only classifiers (DM-200(nc) and DS1-nc), and the top-200 DM gene list for this target is ~16% non-coding, a 1.8× enrichment over the 9% expressed-non-coding base rate (main text).

The ncRNA contribution extends beyond the primary targets. Among the 23 secondary prediction targets evaluated with the expanded mRNA+ncRNA module library, ncRNA modules earned inclusion in 8 of 23 optimal configurations. The largest ncRNA-attributable gains concentrated in targets with immune marker dependencies: T2 lesion burden >15 in MS improved by 4.8% (53.3% to 58.1% AUC at 80% coverage), IgG index elevation in MS improved by 4.5% (53.4% to 57.9%), and timed 25-foot walk >7 seconds in MS improved by 1.5% (68.9% to 70.3%, crossing the 70% clinical-utility threshold). One reading consistent with these gains is that the lncRNA and pseudogene transcripts dominating the MS-specific gene list participate in immune regulatory networks relevant to immunoglobulin status, lesion inflammatory burden, and motor disability; the gains may also reflect non-mechanistic feature richness from the larger pool. The current data does not distinguish these.

We quantified platform dependency by comparison against the Illumina HumanHT-12 v4 BeadChip probe manifest. BIRT detects 1,863 genes absent from the microarray reference, of which 1,654 (89%) are lncRNAs. The overlap between our DS gene lists and those derived from external microarray data (GSE136411) is 3 of 200 genes (1.5%). On these data, the disease-relevant transcriptomic variability we identified resides largely in a molecular space outside the microarray feature pool and outside standard mRNA-seq. The non-coding pool accounts for 68% of the MS-specific top-200 DS list.

#### Non-coding RNA in the BIRT feature pool

Of 63,086 annotated genes on the BIRT platform, 46,722 pass the variance > 0.01 filter; of these, 27,976 (59.9%) are non-coding.

| Gene type | Total genes | Viable genes (var > 0.01) | % of viable |
|---|---|---|---|
| protein_coding | 20,065 | 18,746 | 40.1% |
| lncRNA | 19,258 | 14,335 | 30.7% |
| processed pseudogene | 10,142 | 5,743 | 12.3% |
| unprocessed pseudogene | 2,602 | 1,302 | 2.8% |
| misc_RNA | 2,208 | 1,175 | 2.5% |
| miRNA | 1,879 | 1,050 | 2.2% |
| snRNA | 1,901 | 844 | 1.8% |
| TEC (to be experimentally confirmed) | 1,052 | 788 | 1.7% |
| transcribed pseudogenes | 1,633 | 1,274 | 2.7% |
| snoRNA | 942 | 556 | 1.2% |
| rRNA pseudogene | 497 | 209 | 0.4% |
| IG/TR genes | 518 | 462 | 1.0% |
| Other (scaRNA, rRNA, ribozyme, etc.) | 389 | 238 | 0.5% |
| **Total** | **63,086** | **46,722** | **100%** |
| *of which non-coding* | *43,021* | *27,976* | *59.9%* |

| Module | Genes | mRNA | lncRNA | miRNA | snoRNA | snRNA | IG/TR | Pseudogene | Total ncRNA |
|---|---|---|---|---|---|---|---|---|---|
| DS1 (MS Range80 top 200) | 200 | 64 (32%) | 82 (41%) | 5 | 3 | 4 | 10 | 27 | **136 (68%)** |
| DS2 (NMO Range80 top 200) | 200 | 117 (59%) | 47 (24%) | 4 | 2 | 1 | 13 | 16 | **83 (42%)** |
| DS3 (NMO top 50) | 50 | 32 (64%) | 4 | 0 | 1 | 0 | 11 | 2 | **18 (36%)** |
| DS4 (NMO top 500) | 500 | 268 (54%) | 135 (27%) | 14 | 4 | 2 | 19 | 47 | **232 (46%)** |
| DS5 (MS top 200, PCA) | 200 | 64 (32%) | 82 (41%) | 5 | 3 | 4 | 10 | 27 | **136 (68%)** |

#### Biotype composition of DS gene signatures

Disease-Sigma gene selection preferentially discovers non-coding RNA, particularly for the MS-specific signature, where lncRNAs dominate the top-ranked genes.

#### ncRNA allocation in winning ensembles

GW1-nc (F2 consistency filter applied to the non-coding pool) appears in the MS Subtype primary ensemble; its gene selection is seed-dependent and varies per cross-validation fold. Non-coding modules earn their place in four of the five primary-and-secondary-grade targets listed above, consistent with the biotype-composition pattern (Table below) showing that 68% of the top-200 MS Disease-Sigma signature is non-coding.

#### mRNA-only vs ncRNA-only vs full-model performance

AUC @100% coverage (ablation analysis; the headline AUCs reported in §S5 come from the full validation run and supersede the values shown here):

AUC @80% coverage (same ablation source):

| Primary target | Full config | DS modules | ncRNA modules selected |
|---|---|---|---|
| MS vs NMO | DM-200 + DM-200(nc) + C2 + DS1 + C1 + DS1-nc | DS1, DS1-nc | **2 (DM-200(nc), DS1-nc)** |
| MS Subtype | DS3 + C1 + C4 + GW1-nc | DS3 | **1 (GW1-nc)** |
| MS vs Healthy Control | DS5 + DM-200(nc) + DM-200(pc) + C3 + DS1 | DS5, DS1 | **1 (DM-200(nc))** |
| EDSS >3.5 | C4 + C1 + DS7 | DS7 | **0** |
| Early MS vs Control | DM-50 + DS8 + DM-200 + C3 + DM-200(nc) + DS6 + PCA-64 | DS8, DS6 | **1 (DM-200(nc))** |

| Target | mRNA-only | ncRNA-only | Full model |
|---|---|---|---|
| MS vs NMO | 85.7% | **86.1%** | **86.6%** |
| MS Subtype | **74.1%** | 71.0% | 74.1% |
| MS vs Control | 70.6% | – | **71.5%** |
| EDSS > 3.5 | 68.2% | – | 68.2% |
| Early MS vs Control | 73.1% | 69.7% | **73.7%** |

| Target | mRNA-only | ncRNA-only | Full model |
|---|---|---|---|
| MS vs NMO | 93.5% | **92.9%** | **94.1%** |
| MS Subtype | **78.5%** | 75.9% | 78.5% |
| MS vs Control | 76.1% | – | **77.2%** |
| EDSS > 3.5 | 76.7% | – | 76.7% |
| Early MS vs Control | 77.2% | 73.0% | **78.0%** |

The qualitative pattern is that ncRNA carries diagnostic signal of similar magnitude to mRNA on between-disease targets and that mRNA dominates within-disease. This pattern is reinforced by the primary winning ensembles, which include non-coding modules in MS vs NMO, Early MS, and MS Subtype (see table above). Headline AUCs for all primary targets are reported in §S5.

#### Interpretation

For between-disease classification, ncRNA carries signal of comparable magnitude to mRNA: the ncRNA-only MS vs NMO classifier reaches 86.1% @100% versus 85.7% for mRNA-only. We hypothesize that the structurally symmetric modules (DS1-nc alongside DS1, DM-200(nc) alongside DM-200) selected for the MS vs NMO winning ensemble access overlapping myeloid (DS_MS) and B-cell (DS_NMO) programs through independent molecular features. The data are also consistent with the two pools indexing partially distinct biology that happens to be similarly diagnostic, and we cannot adjudicate between these on classifier performance alone.

For within-disease classification, mRNA dominates but ncRNA still contributes. MS Subtype mRNA-only (74.1%) exceeds ncRNA-only (71.0%) at the ablation level, consistent with the progressive-vs-relapsing signal residing predominantly in protein-coding expression programs. The MS Subtype winning ensemble nonetheless includes one non-coding module (GW1-nc, the genome-wide ncRNA consistency filter) alongside DS3 + C1 + C4, and the ncRNA-only MS Subtype classifier (71.0%) exceeds the 70% clinical-utility threshold on its own.

Standard mRNA-seq protocols and microarray platforms miss the majority of the MS-specific Disease-Sigma signature, and the 3-of-200 gene overlap with external microarray-derived DS gene lists is consistent with that platform-coverage gap rather than with sampling variation. lncRNAs are increasingly studied as regulators of immune cell differentiation and activation, and the enrichment of lncRNAs in the MS-specific myeloid / innate program (led by IL6-AS1, antisense to IL-6) is consistent with that broader literature. Many of the top-ranked MS lncRNAs have no prior disease association in the literature we surveyed; they are candidates for further functional characterization, not validated biomarkers.

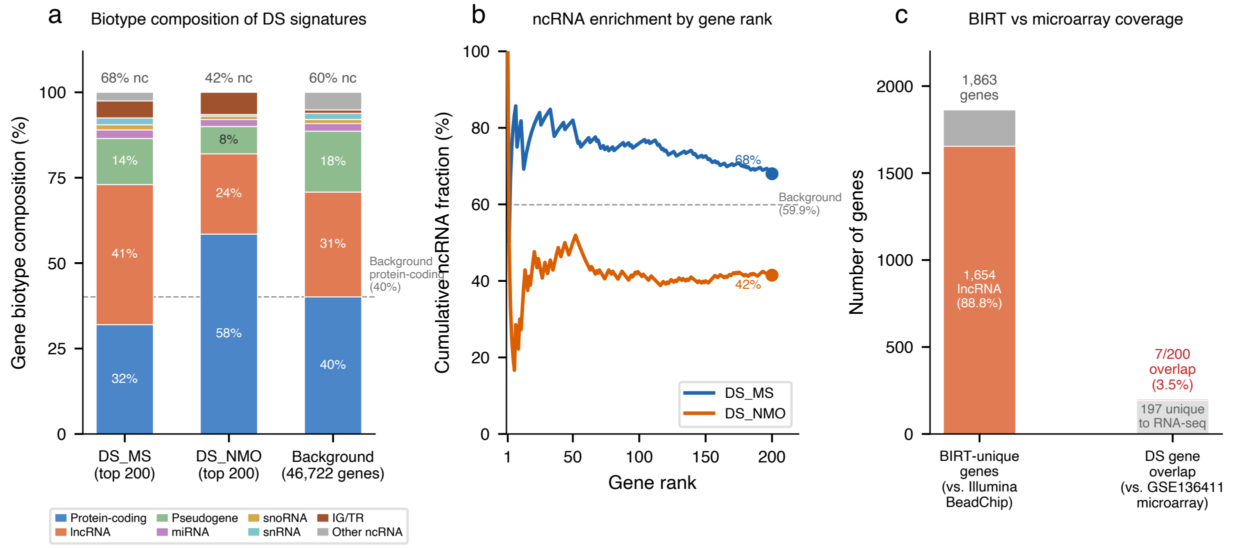

**Figure S11.** Non-coding RNA enrichment in disease-stratified gene signatures.

#### S12. Cell-type decomposition: marker-based proxies versus formal deconvolution

A natural question for any bulk PBMC classifier is whether formal cell-type deconvolution using a single-cell RNA-seq reference improves on marker-based immune composition estimates for disease classification. We tested both approaches directly: a marker-based hierarchical module and a Shannon-entropy diversity module as the proxy approach, and a non-negative least squares deconvolution module against the CellxGene Human Immune Health Atlas as the formal approach.

##### Methods

Two marker-based modules estimate immune cell composition directly from bulk expression. The hierarchical module computes ratios across 6 major PBMC lineages (B cell, CD4 T, CD8 T, monocyte, NK, pDC) using 46 canonical marker genes organized in a two-level hierarchy: a lymphoid/myeloid split at depth 1, then within-compartment ratios at depth 2, yielding 12 features per sample. The diversity module derives Shannon entropy, Simpson index, and Pielou evenness from the cell fraction estimates, capturing immune repertoire balance in 3 features. Both modules are sample-local (no cross-sample normalization) and leak-free by construction.

The deconvolution module is a non-negative least squares (NNLS) engine that accepts an external scRNA-seq reference. We tested it using the CellxGene Human Immune Health Atlas (HIHA) as a 33-cell-type reference signature matrix. The deconvolution solves min $||x - S*w||^2$ subject to $w >= 0$ for each sample, where S is the reference signature and w the inferred cell fractions. We evaluated it solo on five classification targets (MS vs NMO, MS Subtype, MS vs Control, EDSS >3.5, Early MS vs Control) using the same train/test protocol as all other modules.

##### Results

The hierarchical module is strongest on between-disease targets (MS vs. NMO at ~80% @80%) and weakest on CSF-biomarker targets (<50%, near chance), consistent with bulk cell-type ratios carrying most of the signal on tasks where disease populations differ systematically in B-cell or myeloid fraction (Figure S12). The diversity module is complementary rather than redundant: on disability-related targets where compositional entropy drifts with disease severity, it outperforms the hierarchical module by ~10-15%, while the hierarchical module dominates on the classification targets where specific lineage ratios carry the signal.

NNLS deconvolution underperformed the marker-based hierarchical module on every one of the five targets tested, with the largest single-target degradation (~14% below on Oligoclonal Bands). We considered three candidate explanations: (i) scale mismatch between bulk log2(1+counts) and scRNA-seq log-normalized expression; (ii) collinearity among the 33 reference profiles, which destabilizes NNLS solutions; and (iii) cell populations present in the HIHA reference but absent from peripheral blood (e.g., tissue-resident subtypes). We did not isolate which contributes most.

In the primary ensembles, the diversity module enters the MS Subtype winner and several secondary winners (EDSS > 3.5, NMO T2 > 7, In Exacerbation), while the hierarchical module enters the MS vs. NMO winning ensemble alongside the gene-expression modules. Both compositional modules contribute a consistent ~1-2% boost of orthogonal signal on top of the gene-expression core.

#### Interpretation

Two interpretations are consistent with the underperformance of formal scRNA-seq-based deconvolution. One reading is that the disease-discriminating signal in bulk PBMC expression resides primarily in gene expression programs rather than in cell-type proportions, so a higher-resolution cell-type estimate buys little; on this reading the marker-based modules are useful as supporting features (baseline immune state, repertoire balance) but not as primary discriminators. The alternative reading is that the bulk-to-single-cell interface itself is the bottleneck, and a deconvolution method better matched to bulk RNA-seq could recover signal that NNLS against this reference does not. The two readings differ in their prediction for a future experiment using a bulk-trained reference, which the present data does not adjudicate. Either way, the practical observation is that simple marker-gene arithmetic carries the cell-composition contribution in this cohort.

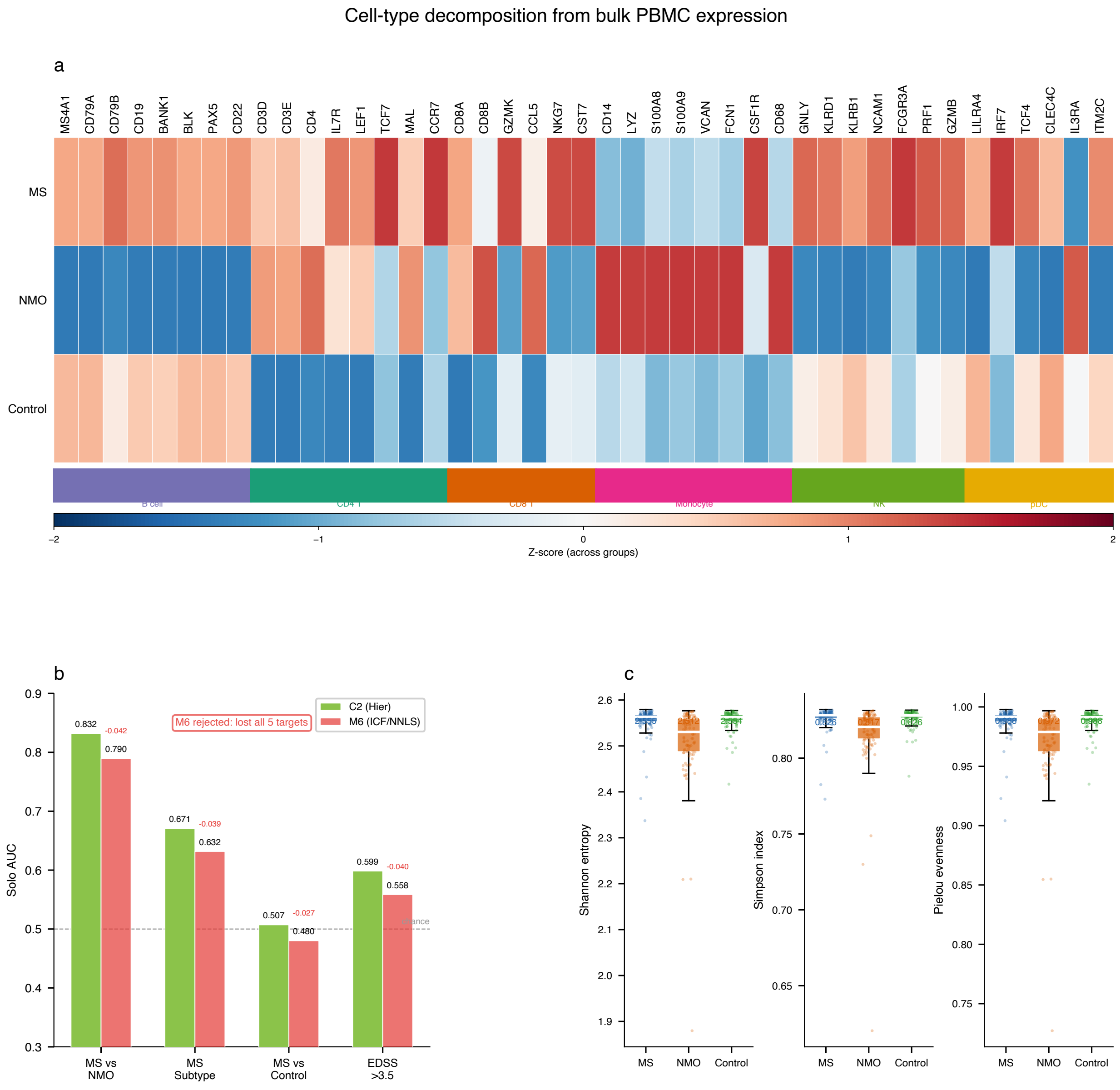

**Figure S12.** Single-cell RNA-seq cell-type decomposition of the bulk PBMC signal.

#### S13. Cross-platform comparison: RNA-seq versus microarray Disease-Sigma gene selection

We asked whether the Disease-Sigma gene signatures identified by BIRT RNA-seq replicate on an independent platform. The comparator was the largest publicly available MS PBMC microarray dataset, GSE136411 (Illumina HumanHT-12 v4 BeadChip; 226 MS patients across CIS, RRMS, PPMS, SPMS; 60 healthy controls; 27 other neurological disease controls). After probe-to-gene aggregation and replicate averaging, the microarray yielded 8,990 measurable genes, less than one-fifth of the 46,722 viable genes in our RNA-seq data, with 8,510 gene symbols overlapping between platforms.

Disease-Sigma (DS) scores were computed on each platform independently using the canonical Range80 metric that defines the DS1/DS2 modules: Range80(MS) minus Range80(HC), where Range80 is the 90th minus 10th percentile of log2 expression within the disease group. Ranking genes by DS score and comparing the top 200, the overlap was 7 of 200 genes (3.5%).

Two non-exclusive interpretations are consistent with the divergence (Fig. S13a). First, an abundance interpretation: the RNA-seq DS top-200 genes have a median expression of log2RPM 2.2, with 99% falling below the microarray reliable-detection floor of log2RPM 3.5; the microarray DS top-200 genes have a comparable median (log2RPM 2.5) but a substantial high-abundance tail reaching log2RPM 4-8 where microarray probes produce reliable signal. The distributions separate in the high-expression tail, not the median. Second, a feature-coverage interpretation: non-coding RNA species account for 68% of the RNA-seq DS top-200 (Section S11), including lncRNAs, pseudogene transcripts, and small RNAs that are either absent from the Illumina BeadChip probe manifest or expressed at levels that produce only noise on microarrays. Both interpretations point to the same operational conclusion, namely that the two platforms sample largely non-overlapping regions of the transcriptomic feature space when ranked by Range80; we cannot from this analysis alone separate cohort-level biological variation between GSE136411 and our cohort from the platform-coverage effect.

The cross-platform intersection nonetheless retains value. Restricting to genes with Range80 > 1 on both platforms yields an AND-filter set of 3,115 higher-abundance, protein-coding-enriched genes that show disease-specific variability on both platforms. The top 200 from this AND-filter (ranked by RNA-seq DS) was used as a gene selection module: solo validation AUC of 0.636 for oligoclonal band prediction, and improvement of the existing module combination from 0.618 to 0.682 (+0.064).

We hypothesize that prior microarray transcriptomic studies in MS, which have typically reported limited discriminative power for clinical phenotypes, may have been constrained in part by the same coverage gap rather than solely by disease heterogeneity or sample size. The data here are consistent with that interpretation but do not establish it; a same-cohort same-sample direct comparison would be the definitive test (Fig. S13).

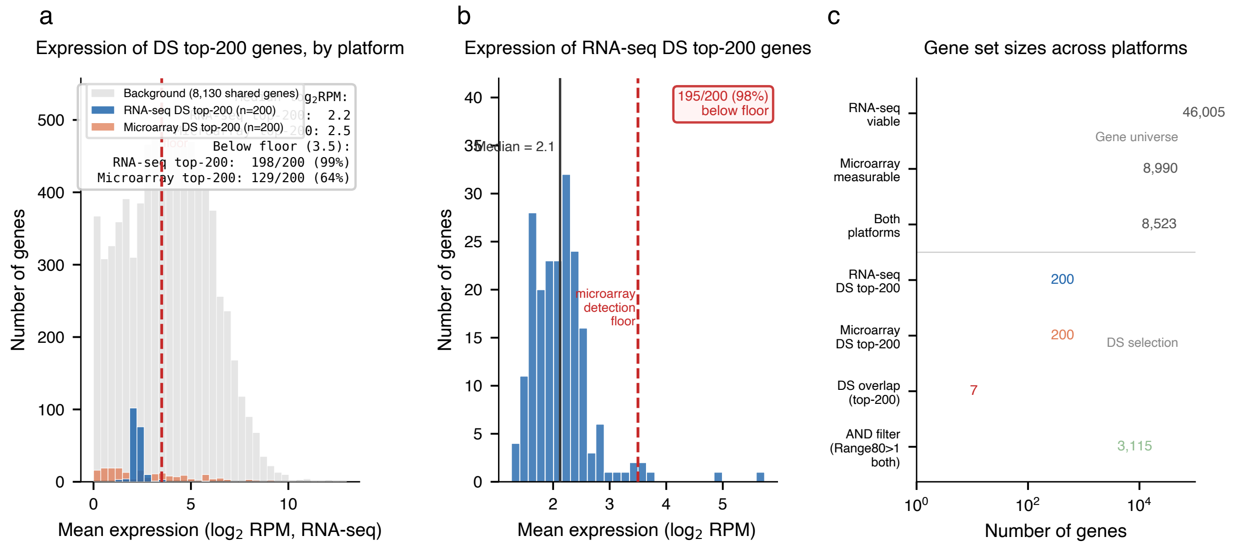

**Figure S13.** Microarray vs BIRT RNA-seq comparison of disease-stratified gene rankings.

### S14. Internal versus external healthy control comparison

Disease-Sigma (DS) scoring requires a healthy reference against which disease-associated expression variability is measured. Two candidate references were available: 149 internal healthy controls profiled on the same BIRT total RNA platform as all disease samples, and 108 independent healthy donors from GSE314922 profiled by standard mRNA-seq. The analyses below ask whether the two references reflect the same underlying biology, characterize platform-specific differences, and quantify the effect of reference choice on DS estimates.

#### Expression concordance confirms shared biology

Mean expression profiles of the 149 BIRT internal controls and 108 GSE314922 donors were compared across 58,072 genes mappable between the two platforms. On protein-coding genes detected in both cohorts, mean expression shows strong concordance (Pearson $r = 0.87$, Spearman $\rho = 0.89$), consistent with the two cohorts sampling the same biological population despite originating from different laboratories and protocols. GSE314922 achieves higher per-sample sequencing depth (30.6M vs 17.7M reads), so the concordance is not driven by a BIRT sensitivity advantage on standard transcripts.

#### Platform differences in non-coding RNA detection

BIRT total RNA sequencing detects 1,863 genes absent from GSE314922 entirely, of which 1,654 (88.8%) are lncRNAs (Fig. S14). Among genes present on both platforms, BIRT detects snoRNAs at 40% higher rates, consistent with the expected advantage of total RNA capture over poly-A-selected mRNA-seq. miRNA detection counts are similar between platforms but differ in gene identity by 23%, plausibly reflecting library preparation biases rather than biological differences. These platform-specific detection profiles do not compromise concordance on shared protein-coding genes but motivate using the same platform for reference and disease comparisons.

#### Range80 calibration reveals platform artifact in external DS

Before quality-covariate correction, BIRT samples exhibit 1.58-fold higher Range80 (P90 minus P10 of log-expression across samples) than GSE314922 on the same protein-coding genes. This systematic offset is consistent with platform dynamic range differences combined with sample-quality heterogeneity in the BIRT cohort, and it inflates the DS score of every gene when external controls serve as the denominator.

The downstream effect is large. With external GSE314922 controls, 3,643 genes exceed DS > 1, leaving the filter without selectivity. With internal BIRT controls, only 367 genes exceed DS > 0.25 after quality-covariate correction (regressing out mitochondrial fraction, ribosomal fraction, and log-total counts). The correction brings both references to convergent Range80 distributions (~1.7 to 2.0). Residual DS rankings remain uncorrelated between internal and external references (Spearman rho = 0.085), so the two approaches rank genes by different criteria even after correction.

#### Housekeeping gene validation

The ACTB result provides the clearest single illustration (Fig. S14). Beta-actin, a canonical housekeeping gene with stable expression across conditions, receives DS_external = +0.66 when scored against GSE314922, which would falsely flag it as disease-specific. With internal controls, ACTB receives DS_internal = −0.07. We observe this pattern across all 12 housekeeping genes tested (GAPDH, ACTB, B2M, RPL13A, RPLP0, UBC, YWHAZ, HPRT1, TBP, SDHA, HMBS, PPIA): the internal reference consistently produces near-zero DS scores, while the external reference assigns positive DS to the majority. The simplest interpretation is that the systematic false-positive pattern reflects the platform Range80 offset rather than biological signal in housekeeping genes.

#### Conclusion

Internal BIRT controls and external GSE314922 donors reflect the same healthy PBMC biology on shared protein-coding genes (r = 0.87), but cross-platform DS scoring introduces a systematic artifact that inflates DS estimates for housekeeping genes and erodes filter selectivity. We therefore use internal healthy controls as the reference for all DS-based gene selection throughout this study.

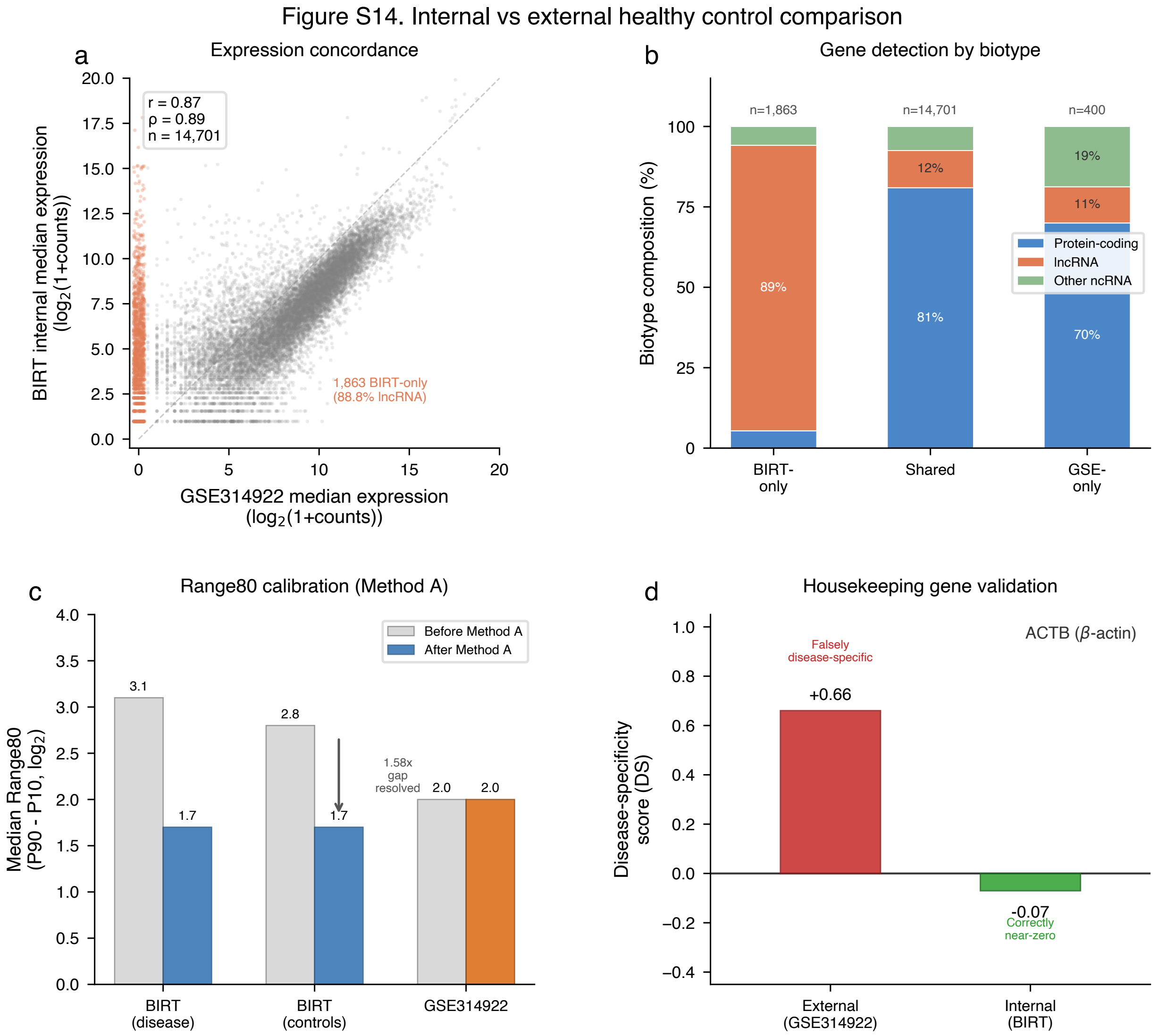

**Figure S14.** Internal BIRT healthy controls vs external GSE314922 reference.

#### S15. Early MS screening: operating points and health economics

The clinical utility of the 7-module Early MS ensemble (74.2% AUC at 100% coverage) depends on deployment setting and pre-test probability of MS. The same classifier produces an NPV of 99.4% in symptomatic primary-care screening (2% base rate, the headline scenario in Fig. S15a) but 93.0% in CIS risk stratification (20% base rate). Predictive values are functions of base rate. We align the primary scenario with the 2% base rate used in Fig. S15a/b and report the full base-rate sweep from unselected population (0.33%) to CIS evaluation (20%). All operating points are derived from the 100-seed mean ROC of the Early MS ensemble, with PPV and NPV computed via Bayes' theorem.

##### Primary scenario: 2% base rate, symptomatic primary-care screening

Main-text Fig. S15a frames the deployment scenario as 1,000 symptomatic patients presenting to primary care with neurological complaints, where the prior probability of an underlying Early MS (EDSS $\leq$ 2.0)

| Mode | Sensitivity | Specificity | Base Rate | PPV | NPV | MRIs avoided / 1,000 | Clinical Use |
|---|---|---|---|---|---|---|---|
| Screening | 90% | 32% | 0.33% | 0.4% | 99.9% | 319 | Unselected population |
| Screening | 90% | 32% | 1% | 1.3% | 99.7% | 317 | General-population screen |
| **Screening (primary)** | **90%** | **32%** | **2%** | **2.6%** | **99.4%** | **316** | **Symptomatic primary care (Fig. S15a)** |
| Screening | 90% | 32% | 5% | 6.5% | 98.4% | 309 | EBV-enriched symptomatic |
| Screening | 90% | 32% | 10% | 12.9% | 96.8% | 298 | High-suspicion neurology referral |
| Screening | 90% | 32% | 20% | 24.9% | 93.0% | 276 | CIS risk stratification |
| Screening (alt.) | 85% | 42% | 0.33% | 0.5% | 99.9% | 419 | Unselected population |
| Screening (alt.) | 85% | 42% | 1% | 1.5% | 99.7% | 417 | General-population screen |
| Screening (alt.) | 85% | 42% | 2% | 2.9% | 99.3% | 415 | Symptomatic primary care (Fig. S15b) |
| Screening (alt.) | 85% | 42% | 5% | 7.2% | 98.2% | 406 | EBV-enriched symptomatic |
| Screening (alt.) | 85% | 42% | 10% | 14.0% | 96.3% | 393 | High-suspicion neurology referral |
| Screening (alt.) | 85% | 42% | 20% | 26.8% | 92.0% | 366 | CIS risk stratification |
| Balanced | 70% | 65% | 2% | 3.9% | 99.1% | 646 | Rule-in adjunct |
| Balanced | 70% | 65% | 5% | 9.6% | 97.6% | 635 | Rule-in adjunct |
| Balanced | 70% | 65% | 10% | 18.3% | 95.2% | 617 | Rule-in adjunct |
| Balanced | 70% | 65% | 20% | 33.5% | 89.7% | 581 | CIS risk stratification |

diagnosis is approximately 2%. At the recommended operating point (90% sensitivity, 32% specificity), the classifier rules out 316 of these 1,000 patients (31.6% fraction-negative) with an NPV of 99.4%, corresponding to 2 missed Early MS cases per 1,000 screened (delayed by 1 year, the annual re-test interval, before being captured at the next draw). PPV at this operating point is 2.6%, reflecting the inherent ceiling on positive predictive value at low base rates rather than a deficiency of the classifier; positives are intended to trigger MRI workup rather than to confirm diagnosis.

A less sensitive alternative operating point at 85% sensitivity / 42% specificity (main-text Fig. S15b) raises the MRI-avoidance fraction to 41.5%, 415 MRIs deferred per 1,000 screened, a 31% increase over the 90% sensitivity point, at the cost of one additional missed case per 1,000 (3 missed, again recoverable at the 1-year re-test). NPV at this point is 99.3% and PPV is 2.9%. The 90% vs. 85% sensitivity choice is the headline trade in main-text Fig. S15: each percentage point of sensitivity at the top of the ROC is expensive to purchase, and the supplementary material quantifies the trade across the full sensitivity continuum below.

#### Table S15-1. Operating points across clinical base rates

Operating points for the Early MS ensemble at three deployment modes, with positive predictive value (PPV), negative predictive value (NPV), and MRI-avoidance fraction at each base rate. All values are derived from the 100-seed mean ROC. The bolded rows mark the recommended primary screening operating point (90% sensitivity / 32% specificity) at the main-text 2% base rate.

Two patterns are visible. First, NPV exceeds 98% across all screening-appropriate base rates ($\leq$ 5%) at both the 90% and 85% sensitivity operating points. Second, the MRI-avoidance fraction is approximately base-rate-insensitive at any fixed operating point (316 $\rightarrow$ 276 over a 10$\times$ change in prevalence at the 90% sens point), because the avoidance rate is dominated by classifier specificity rather than by disease prevalence. As a consequence, the per-patient economics are insensitive to imprecise prevalence estimates in the target population.

| Operating point | Base rate | $f_{neg}$ | Savings/patient | Per 1,000 screened |
|---|---|---|---|---|
| 90% sens / 32% spec | 2% | 0.316 | $964 | $964,000 |
| 90% sens / 32% spec | 5% | 0.309 | $936 | $936,000 |
| 85% sens / 42% spec | 2% | 0.415 | $1,360 | $1,360,000 |
| 85% sens / 42% spec | 5% | 0.406 | $1,323 | $1,323,000 |

#### EBV enrichment as a deployment lever

The 5%-base-rate column in Table S15-1 corresponds to a biologically motivated deployment scenario. EBV-seropositive symptomatic individuals carry a 32-fold increased risk of subsequent MS diagnosis [Bjornevik 2022, *Science*], and the ~95% adult EBV seroprevalence collapses to a smaller subset when combined with symptom presentation (paresthesias, optic neuritis, vague neurological complaints) and an elevated EBNA-1 IgG titer. A symptomatic, EBV-enriched primary-care screening population carries an estimated 5-10% MS base rate; classifier NPV at the 90% sensitivity threshold is 98.4% (5%) to 96.7% (10%) in this range. This is the prospective-validation cohort flagged in the main-text Discussion: a study that pre-screens for EBV serology and presenting symptoms before applying the BIRT assay would fall in this 5-10% base-rate envelope.

#### Health economics

We compute net per-patient savings as the avoided MRI cost minus the test cost and the residual MRI cost on patients who do not get ruled out:

$$\text{savings/patient} \;=\; C_{\text{MRI}} \;-\; C_{\text{test}} \;-\; (1 - f_{\text{neg}}) \cdot C_{\text{MRI}}$$

with $C_{\text{MRI}}$ = \$4,000 (US average for brain MRI with contrast), $C_{\text{test}}$ = \$300 (BIRT panel target price), and $f_{\text{neg}}$ the operating-point fraction-negative.

Two observations. First, savings are stable across base rates ($936–$964/patient at 90% sens, $1,323–$1,360 at 85% sens), because the MRI-avoidance fraction is set by classifier specificity rather than disease prevalence. Second, the 85% sens point delivers ~40% more per-patient savings than the 90% sens point, at a cost of one additional missed case per 1,000 screened. The 90% sens point preserves the rule-out NPV claim across all plausible base rates; the 85% sens point is an alternative for settings where a 1-year delay on a small additional fraction of missed cases is acceptable. The break-even test price at the 90% sens / 2% base-rate point is approximately $1,264, 4.2× above the $300 target.

#### Table S15-2. Structural analog: BIRT screening classifier versus Afirma thyroid GEC

The screening deployment model is structurally analogous to the Afirma thyroid nodule Gene Expression Classifier. Both are high-NPV rule-out tests operating at AUC 73-80%. Both derive clinical utility from the negative majority: Afirma reduces unnecessary thyroid surgeries; the BIRT classifier reduces unnecessary MRI referrals.

Afirma is FDA-cleared and commercially reimbursed. We target $300 for BIRT, reflecting the lower cost of the avoided procedure (MRI versus surgery) and the multi-target panel architecture. Screening adds zero marginal cost to an existing BIRT run: a single blood draw simultaneously screens for MS, differentiates

| Feature | Afirma GEC / GSC | BIRT Early MS classifier |
|---|---|---|
| Clinical question | Rule out thyroid malignancy in indeterminate FNA | Rule out early MS in symptomatic primary care |
| AUC range | 0.75-0.80 (internal validation) | 0.74 (@100%; screening does not report @80%) |
| Operating mode | High-sensitivity rule-out | High-sensitivity rule-out (90% sens, 32% spec) |
| Negative predictive value | ~94-99% (at typical indeterminate FNA prevalence) | 99.4% (2% base rate), 98.4% (5% base rate) |
| Clinical action on negative | Avoid diagnostic thyroidectomy | Defer MRI; annual re-test |
| Clinical action on positive | Proceed to surgery | Proceed to McDonald-criteria MRI workup |
| Test price | ~$3,500 per test | $300 target price |
| Regulatory status | FDA-cleared, commercially reimbursed | Research use; Phase 2 validation planned |
| Avoided procedure cost | Thyroidectomy (~$12,000+) | Brain MRI (~$4,000) |

from NMO, classifies subtype if positive, and estimates disability severity.

#### NPV-first operating point selection and the sensitivity continuum

The screening operating point reported in the main text (90% sensitivity, 32.4% specificity) was chosen *a priori* on sensitivity, a common default for rule-out tests. Fixing sensitivity first and reading specificity off the ROC leaves predictive values as derived quantities: at this point, NPV is 99.4% at the 2% primary base rate and 98.4% at the 5% EBV-enriched base rate, both above the 95% NPV benchmark used by Afirma GSC in its indeterminate-cytology population. The same ROC supports a continuum of operating points. We characterize the full sensitivity-specificity-NPV trade space and identify the operating point that maximizes specificity subject to an NPV floor, which corresponds to the maximum MRI-avoidance policy consistent with each clinical NPV standard.

For a target NPV \$n\$ at base rate \$p\$, the feasible region on the ROC is the set of $(\mathrm{sens}, \mathrm{spec})$ points satisfying

$$\mathrm{NPV}(s, sp; p) \;=\; \frac{sp\,(1-p)}{sp\,(1-p) + (1-s)\,p} \;\geq\; n.$$

Within this region, the ROC point that maximizes specificity is the ”best MRI-avoidance” operating point for that (NPV, base-rate) cell. We computed this point by walking the 100-seed mean ROC of the Early MS ensemble on a dense 5,000-point grid and selecting the maximum-specificity feasible point at each target.

#### Table S15-3. NPV-constrained operating points across base rates (max-specificity solution)

Each row reports the ROC point that maximizes specificity while still meeting the NPV floor at the indicated base rate, along with the resulting MRI-avoidance fraction, missed-case count per 1,000 screened (recover-

| NPV floor | Base rate | Sens | Spec | PPV | Avoid/1k | MS missed/1k | Savings/pt | Payback |
|---|---|---|---|---|---|---|---|---|
| 99% | 2% | 90% | 32% | 2.6% | 316 | 2.0 | $964 | 3.2× |
| 98% | 5% | 85% | 42% | 7.2% | 406 | 7.5 | $1,323 | 4.4× |
| 96% | 10% | 85% | 42% | 14.0% | 393 | 15.0 | $1,272 | 4.2× |
| 95% | 5% | – (trivial: $1-p=0.95$ already meets the target; no-information classifier passes) | | | | | | |
| 95% | 10% | 70% | 65% | 18.3% | 617 | 30.0 | $2,169 | 7.2× |
| 95% | 20% | – (classifier cannot reach 95% NPV at this base rate) | | | | | | |

| Sens | Spec | NPV@2% | PPV@2% | Avoid/1k | MS missed/1k | Savings/pt |
|---|---|---|---|---|---|---|
| 95% | 16% | 99.4% | 2.3% | 158 | 1.0 | $332 |
| **90%** | **32%** | **99.4%** | **2.6%** | **316** | **2.0** | **$964** |
| 85% | 42% | 99.3% | 2.9% | 415 | 3.0 | $1,360 |
| 80% | 52% | 99.2% | 3.3% | 513 | 4.0 | $1,752 |
| 75% | 58% | 99.1% | 3.5% | 568 | 5.0 | $1,972 |
| 70% | 65% | 99.1% | 3.9% | 643 | 6.0 | $2,272 |
| 65% | 71% | 99.0% | 4.4% | 703 | 7.0 | $2,512 |

able at the 1-year annual re-test), per-patient net savings ($300 test, $4,000 MRI), and test-price payback. Dashes mark cells where either the classifier cannot reach the NPV target at that base rate, or the target is trivially satisfied by the no-information classifier (NPV floor $\leq 1-p$) and the test adds no value.

Three observations. First, at the main-text 2% base rate, the 99% NPV floor is met at the recommended (90% sens / 32% spec) operating point with margin; this is the deployment choice reported as primary. Second, at the 5% EBV-enriched base rate, the maximum-specificity operating point satisfying a 98% NPV floor is (85% sens, 42% spec), which matches the alternative point shown in main-text Fig. S15b. Third, Afirma's 95% NPV benchmark is not directly applicable to the primary deployment scenario here: at 2–5% base rates the no-information rate is 95–98%, so the 95% NPV target is trivially satisfied by an all-negative classifier. 95% NPV becomes a meaningful target only at higher base rates; on this ROC it is reachable at the 10% base rate (70% sens, 65% spec, 617/1,000 avoiding MRI, $2,169/patient savings) but unreachable at 20%.

#### Table S15-4. ROC continuum at the 2% base rate (single NPV floor = 99%)

Illustrates the cost of each sensitivity increment along the ROC near the recommended primary operating point. All rows satisfy NPV ≥ 99% at the 2% base rate.

The bolded row is the primary operating point shown in main-text Fig. S15a; the 85% row is the alternative operating point in main-text Fig. S15b. Each step down the table trades approximately one missed Early MS case per 1,000 (delayed 1 year) for $400-$650 of additional per-patient savings, with the trade approximately linear across this range. The choice between operating points is a clinical-economic judgment, not a methodological one. We report 90% sens as the primary operating point because it preserves a >99%

| Base rate | Population | Rationale |
|---|---|---|
| 0.33% (1/300) | General population, high-incidence regions | MS prevalence ~1/1,000 globally, ~1/300 in Northern Europe and North America |
| 1% | Primary care, any neurological complaint | ~3× population prevalence among patients presenting with neurological symptoms |
| 2% | Primary care, suggestive symptoms | Patients with numbness, tingling, optic neuritis, or Lhermitte sign, compatible with MS but non-specific |
| 5% | Neurology referral, suspected demyelination | Pre-selected by symptom pattern for specialist evaluation; overlaps EBV-enriched symptomatic cohorts |
| 10% | Specialist workup, strong suspicion | Active diagnostic evaluation for MS with multiple suggestive findings |
| 20% | CIS patients | Clinically isolated syndrome; 60–80% eventually convert to MS, ~20–30% pre-test probability at initial presentation |

NPV margin at the 2% base rate; the alternatives are tabulated for clinical settings whose cost curves favor a different point on the continuum.

##### Base-rate rationale

The clinical utility of any screening test depends critically on the pre-test probability of disease in the target population. We evaluate performance across six base rates spanning the full range of clinical scenarios:

##### Deployment scenarios

US primary care (2% base rate): Blood test at primary care visit; negative result (~32% of patients) defers MRI referral; positive result (~68%) receives neurology referral and McDonald-criteria MRI workup. Estimated net savings: ~$964/patient at the 90% sensitivity / 32% specificity primary operating point.

Neurology referral triage (5% base rate): Applied at the neurology referral stage for patients with suspected demyelinating disease. Per-patient net savings ~$936 at the 90% sensitivity operating point; the test has the dual benefit of triaging MRI access and providing early disease characterization for positive cases.

EBV-enriched symptomatic (5–10% base rate): Biologically motivated cohort: EBV-seropositive symptomatic individuals carry a 32-fold increased risk of subsequent MS diagnosis. NPV at the 90% sensitivity threshold is 98.4% (5%) to 96.8% (10%), the natural prospective-validation cohort flagged in the main-text Discussion.

CIS risk stratification (20% base rate): At the balanced operating point (70% sensitivity, 65% specificity),

PPV reaches 33.5% and NPV is 89.7%. Decision-support for DMT initiation in specialist settings; distinct commercial application from population screening (smaller market, higher price point, directly informs a binary treatment decision).

Resource-limited settings: Where MRI access is constrained (tier-2/3 cities in China and India, Middle East), the alternative to a blood test is not ”$4,000 MRI” but ”no diagnostic workup at all.” The economics flip from cost savings to enabling any diagnosis. Particularly relevant for China, where the NMO-to-MS prevalence ratio approaches 1:1 and specialized AQP4 cell-based assays are less accessible.

#### Important caveats

1. All operating points derive from cross-validated single-cohort evaluation; prospective validation in an independent screening population is required before clinical deployment. 2. The base rates are estimates, actual MS prevalence in specific symptomatic populations depends on referral patterns, demographics, and clinical setting. 3. The MS-versus-healthy-control training involves some residual site and visit-number confounding despite the Early MS subgroup partially mitigating it. 4. Economics assume $300 test price and $4,000 average MRI cost; actual costs vary by region and payer. 5. PPV and NPV are functions of base rate; clinicians should use the base-rate row most appropriate to their clinical setting.

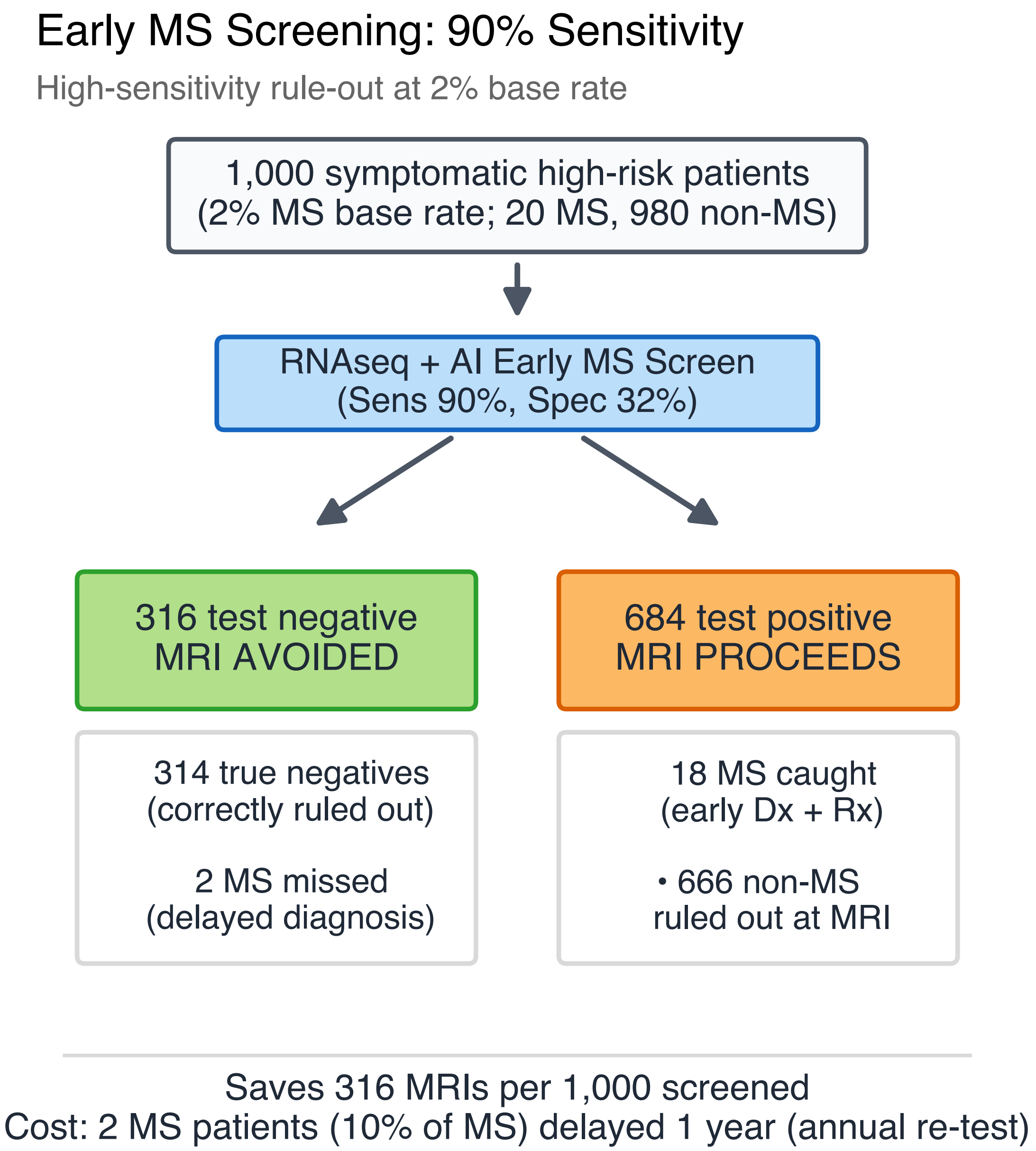

**Figure S15a.** Early MS screening deployment at the 90% sensitivity operating point.

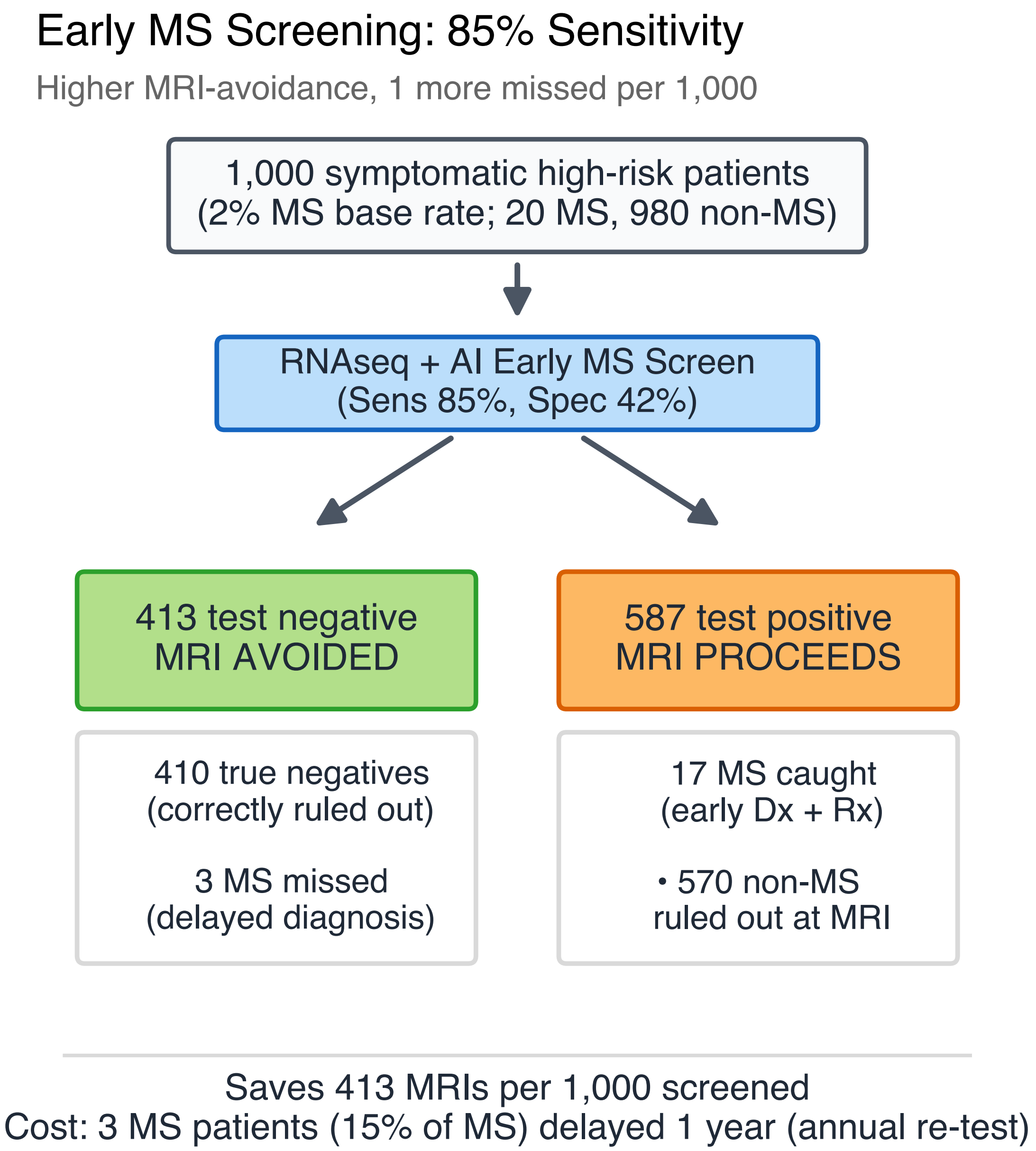

**Figure S15b.** Early MS screening deployment at the 85% sensitivity operating point.

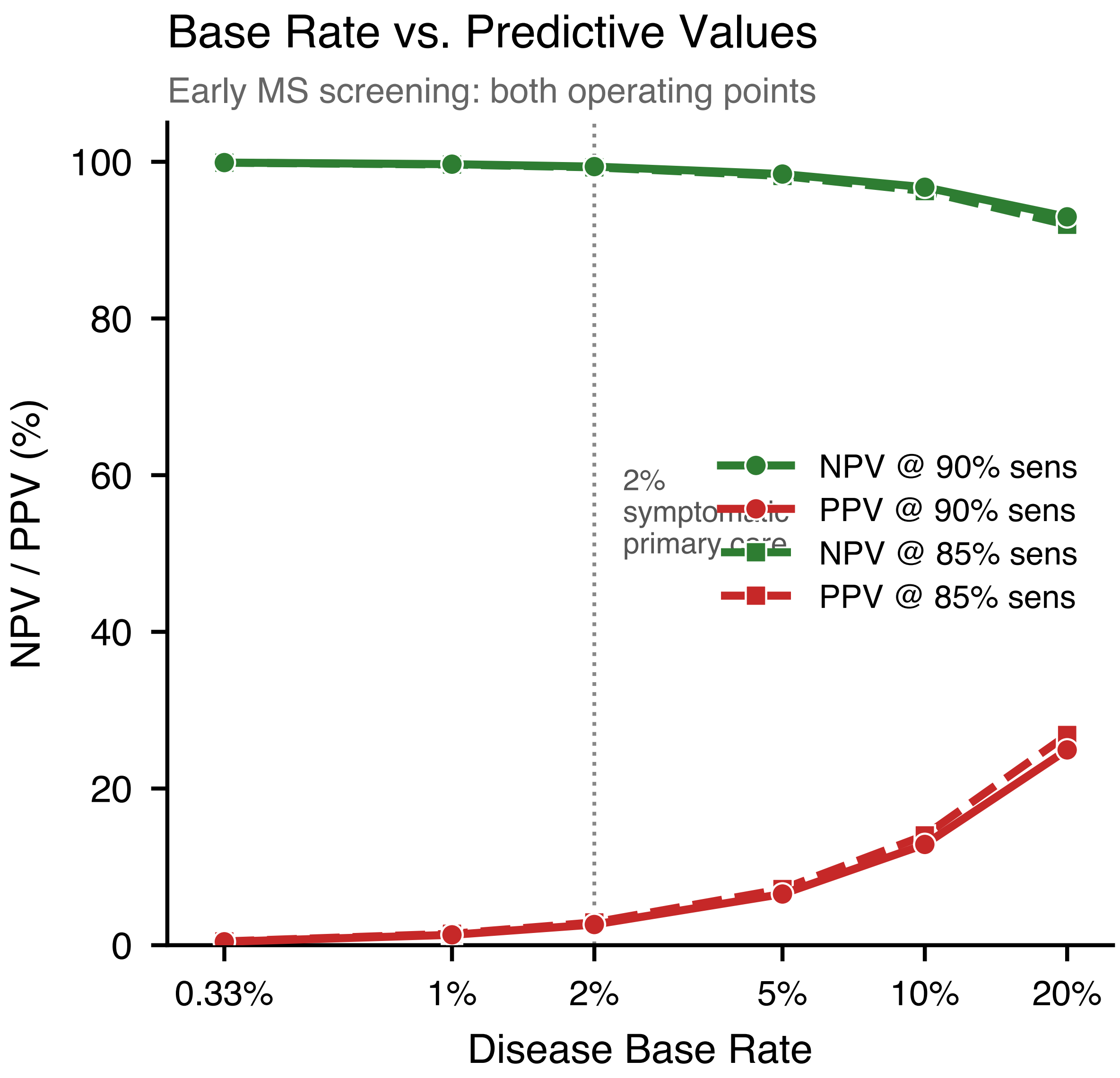

**Figure S15c.** Base rate vs NPV and PPV across both operating points.

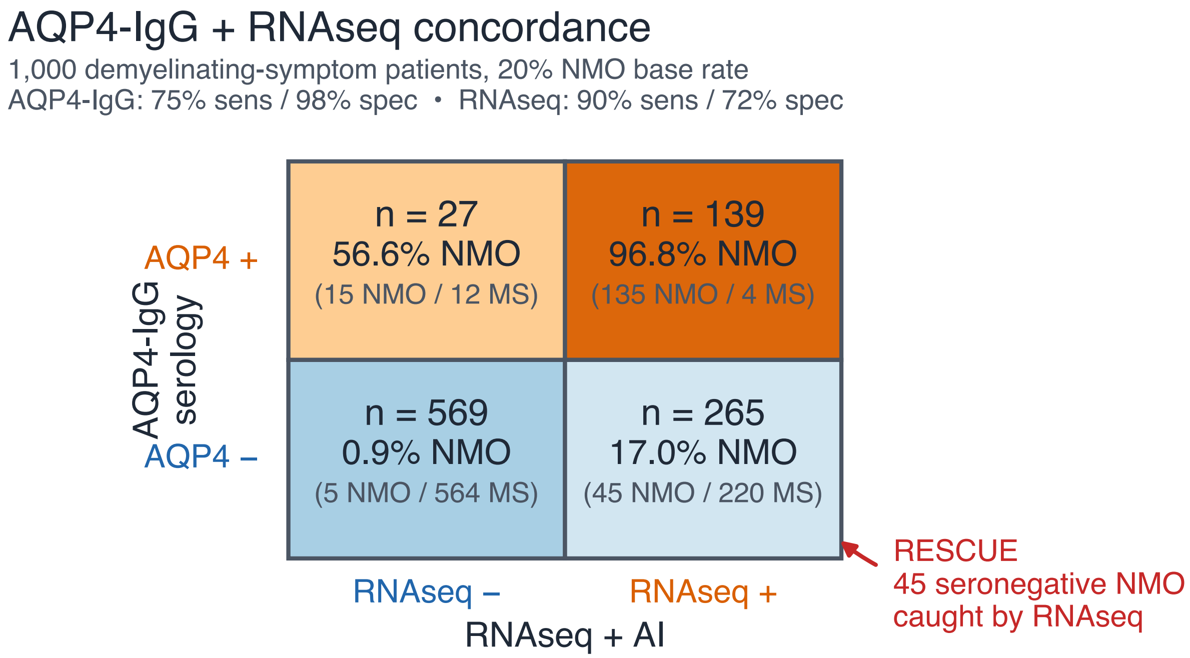

**Figure S15d.** AQP4-IgG × RNAseq concordance matrix; seronegative-NMO rescue cell.

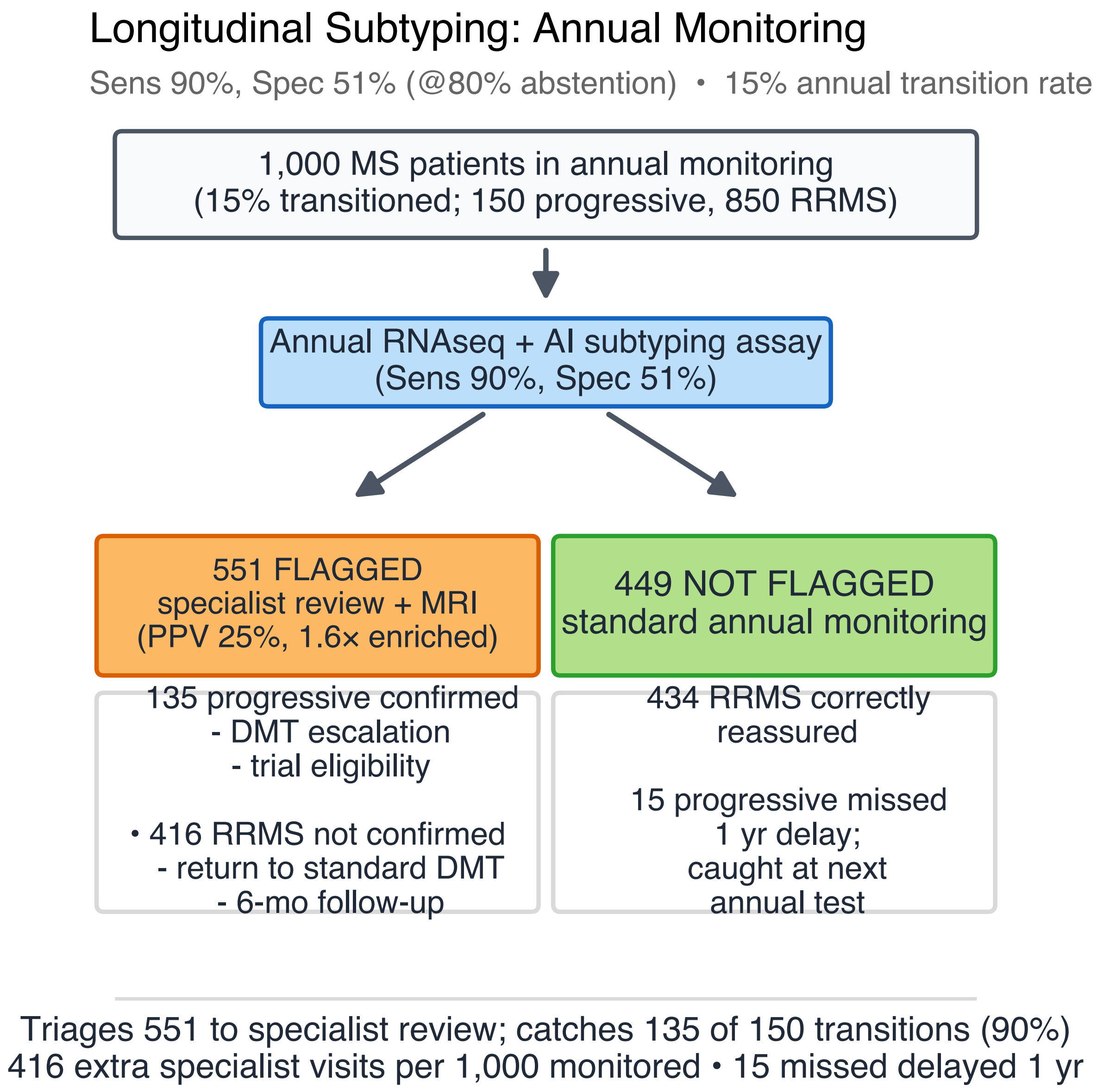

**Figure S15e.** Longitudinal annual-monitoring deployment for progressive-MS detection.

#### S16. Clinical significance and deployment context

Three features define the clinical burden of MS and related neuroimmune diseases: high prevalence, delayed diagnosis, and misallocated treatment. Approximately 2.8 million people live with MS globally, including ~1 million in the United States, with aggregate annual healthcare expenditure exceeding $28 billion. Disease-modifying therapies (DMTs) cost $70,000–90,000 per patient per year, with class-dependent efficacy: anti-inflammatory DMTs reduce relapse rates by 50–70% in relapsing-remitting MS but show minimal effect against progressive neurodegeneration. Patients who silently transition to secondary progressive MS remain on therapies that no longer address their disease. The average diagnostic delay from first symptom to confirmed MS diagnosis spans 4–7 years, during which patients accumulate irreversible disability without treatment. No blood-based biomarker exists for MS diagnosis, subtyping, or screening.

PBMC transcriptomics analyzed with Disease-Sigma AI classifiers maps to three distinct clinical applications from a single blood draw.

##### NMO differential diagnosis

MS and NMO require different treatment strategies, and misclassification carries iatrogenic risk: first-line MS therapies (interferon-beta, natalizumab) can provoke fulminant NMO relapses. The current gold standard, AQP4-IgG cell-based assay, achieves high specificity but misses ~25% of NMO cases, leaving seronegative patients without confirmatory molecular evidence. The BIRT classifier reaches 91.3% AUC at 80% coverage with 90% sensitivity and 72% specificity at the recommended operating point, detecting the broader B-cell transcriptomic signature of NMO rather than a single autoantibody. The intended use is as a second molecular line of evidence for patients who currently lack one. Among 1,000 differential-diagnosis patients at 20% NMO base rate, the AQP4−/RNAseq+ discordant cell would identify ~45 seronegative NMO patients per 1,000 tested (90% RNAseq sensitivity × 25% AQP4-seronegative fraction × 20% NMO base rate × 1,000; Fig. S15d). At the rank-ordered @80% retained ensemble, PPV reaches 45% at 20% base rate and 76% at a 50% clinically-referred base rate, with NPV 96.7% and 87.8% respectively, consistent with a high-NPV rule-out regime appropriate for molecular serology adjuncts.

##### MS subtype classification

The transition from relapsing-remitting to secondary progressive MS is the central prognostic event in the disease, yet clinicians currently diagnose it only retrospectively after months to years of observation, and no blood-based test exists. The BIRT classifier reaches 78.8% AUC at 80% coverage with 90% sensitivity and 41% specificity at the recommended operating point. The deployment model we envision is annual screening of the prevalent relapsing-remitting population; a missed transition would be caught at the next annual re-test rather than accumulating an open-ended diagnostic delay, analogous to mammography for a known-risk group.

##### Early MS screening

At 90% sensitivity, the classifier reaches >98% NPV across screening-appropriate base rates (2–10%), supporting rule-out from a single blood draw at the primary care visit: a negative result defers MRI; a positive result triggers standard diagnostic workup. At a $300 test price and 2% symptomatic-primary-care base rate, the cost model in Section S15 yields net savings of ~$964 per patient screened (316 MRIs avoided per 1,000 at 90% sensitivity), with break-even above $1,264, ~4× the target test price.

##### Staging of clinical applications

We rank these applications by clinical and regulatory readiness. NMO differential diagnosis is the most mature: it has the highest AUC in the panel, the clearest clinical pathway (complementing an existing gold-standard test), and the most defined target population (newly diagnosed patients in differential workup). Subtype classification is next, addressing the larger prevalent population of relapsing-remitting MS patients at risk of transition. Population-level screening requires prospective validation in an independent cohort and

| Dimension | RNA-seq + AI (BIRT) | Brain MRI | Immunoassays (AQP4-IgG) |
|---|---|---|---|
| Information per test | 63,000+ RNA markers, multiple simultaneous predictions | Structural CNS imaging, one clinical question per scan | Single autoantibody titer |
| Clinical scope | Differential, subtype, screening, disability from one draw | Lesion detection, dissemination criteria | NMO confirmation (seropositive only) |
| Biological window | Peripheral immune dysregulation driving disease | CNS structural damage (downstream consequence) | Single effector molecule |
| Scalability | Each additional prediction costs only computation | Each additional question requires a new scan | Each additional analyte requires a new assay |

represents the longest development timeline.

##### Multi-target amortization

A single blood draw at ~\$16 fully-loaded cost-of-goods produces >63,000 RNA measurements, from which multiple clinical predictions are derived computationally at zero marginal cost per additional target. MRI answers one clinical question per scan at \$1,600–8,400. AQP4-IgG answers one question per assay at \$730–860. The RNA-seq/AI approach answers three or more questions per test, with per-question cost that falls as additional validated targets are added.

This platform is intended to complement, not replace, existing diagnostic modalities. MRI remains the gold standard for McDonald criteria confirmation. AQP4-IgG remains definitive for seropositive NMO. BIRT addresses three specific gaps: triage before MRI in symptomatic primary care, inter-scan monitoring for progressive transition biology, and molecular evidence in seronegative NMO patients who currently have none.

All results reported here derive from cross-validated single-cohort evaluation. Prospective validation in independent populations is required before clinical deployment. An expanded cohort of ~2,000 additional samples is in preparation to address the primary limitations of the current study: site confounding in the MS vs. NMO comparison, sample size constraints for disability prediction, and multi-site replication.

#### S17. Biological interpretation of DS-selected genes

##### Overview

Disease-Sigma (DS) gene selection surfaces genes by signed ΔRange80, the difference between the 10th-to-90th-percentile spread of log2 expression in cases versus comparators, rather than by mean-shift. To test whether this variance-first ranking recovers biologically coherent MS and NMO signal, we reviewed 54 DS-selected genes against published PubMed/OMIM literature across three contrasts (Subtype DS in the MS-only cohort; DS_MS in MS vs internal control; DS_NMO in NMO vs internal control) and two complementary ranking lenses: raw Abs(ΔRange80) top-*N* per pool and classifier-weight driver analysis (PCA loadings for DS_MS and DS_NMO; logistic-regression coefficients for Subtype DS). Each gene was graded on a four-point rubric: **Defensible** (direct published MS/NMO/autoimmune link and a plausible variance-shift mechanism), **Plausible** (pathway-level link requiring some interpolation), **Speculative** (no specific link; generic immune-cell argument only), and **Null** (gene biology entirely unrelated, not encountered in the reviewed set). The section opens with the synthesis and continues into the full per-gene catalog.

| Batch | Pool / lens | N reviewed | Defensible | Plausible | Speculative | Null |
|---|---|---|---|---|---|---|
| 1 | Subtype DS, Abs(ΔR80) ranks 1–10 | 10 | 3 | 5 | 2 | 0 |
| 2 | Subtype DS, Abs(ΔR80) ranks 11–30 | 14 | 7 | 3 | 4 | 0 |
| 3 | DS_MS + DS_NMO, Abs(ΔR80) headliners | 11 | 6 | 5 | 0 | 0 |
| 4 | PCA/LR drivers across 3 pools | 19 | 9 | 9 | 1 | 0 |
| **Total** | | **54** | **25 (46%)** | **22 (41%)** | **7 (13%)** | **0 (0%)** |

**IL1B (Fig. 6a), relapse-driven variance inflation**

| Group | n | p10 | p50 | p90 | Range80 |
|---|---|---|---|---|---|
| Control | 149 | 8.22 | 10.21 | 11.61 | 3.39 |
| MS in exacerbation | 54 | – | – | – | 5.37 |
| MS not in exacerbation | 418 | – | – | – | 4.38 |
| MS overall | 540 | – | – | – | 4.49 |
| NMO | 221 | – | – | – | 3.84 |

#### Verdict summary across all four batches

Across 54 genes spanning three DS contrasts and two ranking lenses, 87% received a Defensible or Plausible verdict and none received a Null verdict. The three anchor genes profiled below (IL1B, IFI44, MS4A1) are each Defensible. Speculative calls concentrate in low-stability PCA hits and in low-expression near-floor transcripts (LZTS3, POU6F1, TKTL1) where measurement-level heteroscedasticity cannot be excluded as the variance source.

#### Three mechanistic anchors: IL1B, IFI44, MS4A1

For the three genes featured in main-paper Fig. 6, we quantified Range80 within clinically-stratified subgroups of the 997-sample cohort. For each gene we name the candidate source of bimodality and contrast the variance-shift framing with the mean-shift framing a DM module would apply.

##### IL1B (Fig. 6a), relapse-driven variance inflation

IL-1$\beta$ is the prototypical NLRP3/caspase-1-cleaved inflammasome cytokine, and drives pathogenic Th17 polarization, BBB permeability, and microglial activation in MS and EAE (Lin & Edelson, PMC5509030; Pare et al. *Acta Neuropathol* 2020, doi:10.1007/s00401-020-02187-x). The MS literature explicitly documents that bulk PBMC/CSF IL-1$\beta$ measurements are ”non-univocal” across cohorts, with the spread attributed to patient heterogeneity and therapy effects (Lin & Edelson review), a near-verbatim description of a variance-shift signature.

**IFI44 (Fig. 6b), interferon-β treatment bimodality**

| Group | n | p50 | Range80 |
|---|---|---|---|
| Control | 149 | 8.25 | 3.60 |
| MS not on interferon | 350 | 8.49 | 4.03 |
| MS on interferon-β | 181 | 9.82 | 4.45 |
| MS overall | 540 | – | **5.02** |

Range80 in MS patients captured during exacerbation is 5.37 versus 4.38 in non-exacerbation MS and 3.39 in controls, consistent with relapses driving a long right-tail on IL1B. The exacerbation subgroup is only ~10% of MS samples, which would dilute any mean shift computed on MS-overall (4.49 vs 3.39 controls) but inflates the cohort variance. Two readings of this pattern are consistent with the data: relapse-driven transient induction in a minority subgroup, or chronic IL1B priming with stochastic flares. Either way, the DS framing recovers the bimodality directly, while a DM framing depends on the relapse-active fraction sampled.

##### IFI44 (Fig. 6b), interferon-$\beta$ treatment bimodality

IFI44 is a canonical type-I interferon-stimulated gene (ISG) and forms part of the validated IFN-$\beta$ pharmacogenomic signature used to stratify MS treatment responders (van Baarsen et al. *PLOS ONE* 2008, doi:10.1371/journal.pone.0001927; Hundeshagen et al. *J Neuroinflamm* 2012, PMID 22873631). Critically, untreated MS patients with clinically active disease often show *reduced* ISG expression with subnormal STAT1 phosphorylation (Feng et al., PMID 12161037), so the direction of IFI44 perturbation is not monotonic.

The two MS subgroups have Range80 ≈ 4.0–4.5, similar to the 3.60 control baseline. MS-overall has Range80 = 5.02, wider than either subgroup alone, consistent with mixing an IFN-$\beta$-induced p50 = 9.82 subgroup with an untreated p50 = 8.49 subgroup to create pooled bimodality. We hypothesized that this is the dominant source of the IFI44 variance signal, though residual within-group variance from infection load or untreated-disease-activity heterogeneity may also contribute. A mean-shift test on MS-overall versus control would flag a shift driven by the treated fraction; the DS lens recovers the mixture without pre-specifying the stratifier.

##### MS4A1 / CD20 (Fig. 6c), anti-CD20 depletion bimodality

MS4A1 encodes CD20, the B-cell surface tetraspanin targeted by rituximab, ocrelizumab, ofatumumab and ublituximab. Anti-CD20 monoclonals are first-line therapy for AQP4-IgG$^+$ NMOSD (Cree et al. *Neurology* 2005, doi:10.1212/01.wnl.0000159399.81861.d5; Tahara et al. *CNS Drugs* 2025, doi:10.1007/s40263-025-01191-7) and depletion is near-complete in treated patients.

Anti-CD20 drops the NMO median MS4A1 from 9.78 to 2.00, a 7.78-log2 depletion gap between treated and untreated NMO patients. Within the untreated-NMO subgroup alone, Range80 = 10.14, which the data

**MS4A1 / CD20 (Fig. 6c), anti-CD20 depletion bimodality**

| Group | n | p10 | p50 | p90 | Range80 |
|---|---|---|---|---|---|
| Control | 149 | – | 10.22 | – | 3.94 |
| MS not on anti-CD20 | 482 | – | 10.49 | – | 4.55 |
| MS on anti-CD20 | 58 | – | 11.30 | – | 6.61 |
| NMO not on anti-CD20 | 146 | 2.00 | 9.78 | 12.14 | **10.14** |
| NMO on anti-CD20 | 75 | 0.32 | **2.00** | 7.55 | 7.23 |

| Pool | Top-20 PCA drivers ∩ top-30 Abs(ΔR80) | Overlapping genes |
|---|---|---|
| Subtype DS (n=507, Abs(LR coef)) | **2 / 20** | BLK, PEX16 |
| DS_MS (n=689, PCA10) | **0 / 20** | – |
| DS_NMO (n=370, PCA5) | **3 / 20** | MS4A1, PAX5, COL19A1 |

permits to read either as residual heterogeneity in B-cell-compartment composition (recent-relapse status, preserved vs expanded plasmablast fractions, all of which are CD20$^-$) or as patient-intrinsic variation in CD20 surface expression. The treated-vs-untreated bimodality is the larger of the two effects. A mean-shift test comparing NMO versus control on MS4A1 would give a direction that depends on the treated fraction sampled; the DS lens is indifferent to that mixture ratio. MS on anti-CD20 shows a smaller version of the same pattern (Range80 6.61 vs 4.55 untreated), consistent with partial peripheral depletion under fixed-interval rather than continuous suppression.

#### PCA-driver and Abs(ΔRange80) views are near-orthogonal

A natural reader question is whether the PCA/LR driver lens used in the DS screening-arm composites duplicates the raw Abs(ΔRange80) top-30. The convergence is in fact low:

The two lenses reward different statistics: Abs(ΔRange80) rewards lone-wolf variance, scoring each gene on its own distribution shift regardless of whether that shift is correlated with other top-200 genes; PCA rewards covariance structure, scoring a gene by how strongly it loads onto a coordinated axis of variation that the classifier can compress into a single feature. The DS_MS pool illustrates the divergence: the Abs(ΔRange80) top-30 is dominated by mesenchymal/matrix genes (SDC2, INHBA, VEPH1, PLAU, GP9, PPARG, TGM2) consistent with activated-monocyte and repair biology, while the PCA-driver top-20 is dominated by the type-I IFN signature (IL6, CMPK2, IFI44, IFI44L, TRIM34, TRIM6-TRIM34, PYCARD) and a memory-B / plasmablast cluster (CLEC17A, FCRL2, CD19), both high-covariance gene families that load heavily on individual PCs even when each constituent gene's ΔRange80 is modest. A screening composite built on PCA-compressed DS features (our DS4/DS5 architecture) and a biomarker list derived from raw Abs(ΔRange80) sampling therefore index different axes of the data; we report both rather than treat one as a summary of the other.

#### Six coherent biological clusters across DS pools

The 54 reviewed genes fall into six mechanistically coherent clusters, summarised below with representative members and canonical mechanisms.

In the Subtype DS (MS-only, Progressive vs. RRMS) pool, the dominant cluster is B-cell compartment heterogeneity (NIBAN3, EBF1, CXCR5, LINC00926; four of 14 batch-2 genes are B-cell lineage markers). Variance expansion in progressive MS is mechanistically expected given heterogeneous anti-CD20 exposure, meningeal ectopic follicle burden [21] varying between patients, and EBV-driven B-cell transformation via the EBF1-EBNA2 axis (Glaser et al. *PNAS* 2022, PMID 35858428) on top of the Bjornevik et al. EBV causal link [22]; EBF1 is also a replicated MS susceptibility locus (rs1368297; Prieto et al. *BMC Neurol* 2005, PMID 16255771), which adds an eQTL-driven variance source directly attributable to genetic architecture. A smaller microglia / TGF-$\beta$ axis cluster contains ITGB5 (a canonical TGF-$\beta$-dependent microglial signature gene; Butovsky et al. *Nat Neurosci* 2014, PMID 24316888), SMPD2 (ceramide signaling downstream of TNFR1), and RAB3IP (Rab8a-dependent TLR4 pro-inflammatory trafficking), and tracks the progressive-MS shift toward sustained innate/microglial activation. A single-gene immunosenescence signal is contributed by TNFRSF10D / DcR2, a TRAIL decoy receptor and established senescence marker consistent with the older, more senescence-heterogeneous progressive-MS cohort (López-Gómez et al. *PLOS ONE* 2011).

In the PCA-driver pools (DS_MS and DS_NMO), three additional clusters emerge. A Type-I interferon signature (IL6, CMPK2, IFI44L, TRIM34, TRIM6-TRIM34, PYCARD) rides the treatment-stratified ISG bimodality documented for IFI44 above; this cluster is absent from the DS_MS Abs(ΔRange80) top-30 and is recovered only through PCA compression of coordinated ISG co-variation. A B-cell memory / plasmablast cluster (CLEC17A, FCRL2, CD19, complemented by the DS_NMO Abs(ΔRange80) leaders MS4A1, CD79A, PAX5, BLK, POU2AF1) is consistent with the anti-CD20 depletion bimodality of MS4A1 above, with possible additional contributions from BCR repertoire heterogeneity and plasmablast fraction. A cytotoxic / innate-lymphocyte cluster (KLRF1 as NK activating receptor, ZBTB16 / PLZF as iNKT/$\gamma\delta$ master TF, GZMA as granzyme A pyroptosis, BCAR3 as plasmablast scaffolding) is missed by the DS_NMO Abs(ΔRange80) top-30, which is dominated by the $CD20^{+}$ B-cell signal. The PCA lens recovers it as a coordinated covariance axis.

#### Validated MS-susceptibility loci recovered by DS

Two DS-selected genes are replicated MS susceptibility loci. EBF1 (Subtype DS, Abs(ΔR80) rank 5) carries an intronic polymorphism rs1368297 associated with MS risk in Spanish cohorts, with larger effect after HLA-DRB1*1501 stratification (Prieto et al. *BMC Neurol* 2005, PMID 16255771); EBF1 also complexes with the EBV EBNA2 oncoprotein to drive MYC transactivation in EBV-infected B cells (Glaser et al. *PNAS* 2022, PMID 35858428), connecting DS to the EBV-MS axis [22]. GATA2 (Subtype DS, Abs(ΔR80) rank 11) is a hub gene downregulated in MS by recent transcriptomic meta-analysis (Hassanzadeh et al. *Genomics Inform* 2024), and germline GATA2 haploinsufficiency causes MonoMAC syndrome with aggressive MS reported in a subset of carriers (Spinner et al. *Blood* 2014, PMID 25707267). Both loci were ranked by Abs(ΔRange80) without conditioning on genotype or mean-shift direction.

#### Honest caveats

Four caveats temper the clean-signal story. Low-expression near-floor transcripts including LZTS3 and POU6F1 (both Subtype DS, ranks 3 and 10) are brain-enriched with low PBMC expression near the dropout floor, so their variance shifts are consistent with dropout-driven heteroscedasticity in addition to (or instead of) real CNS-leakage biology; we flag them transparently rather than retrofit a leakage narrative, and TKTL1 (Subtype DS, rank 10) carries the same concern. For compositional vs. intrinsic variance, B-cell lineage markers (PAX5, BLK, POU2AF1, CD79A) show variance shifts largely driven by between-patient B-cell fraction differences (treatment status, plasmablast expansion) rather than by intrinsic per-cell transcriptional dysregulation; DM would flag these too, just with less precision, so we do not claim DS-specific insight on this subcluster, only that DS recovers compartment biology faithfully. Approximately a quarter of Subtype DS hits lack a clear MS mechanism at the current level of literature (TM2D2, TKTL1, LZTS2, ARMCX5 are reasonable lit-review near-misses), and the right frame for these is "surface a candidate subset for follow-up work" rather than "clean mechanistic signature"; the paper's claim is calibrated to a coherent *majority* of hits, not to the tail. Finally, the sample is curated rather than exhaustive, covering the top of each ranking (Abs(ΔR80) top-30 per contrast plus PCA-driver top-20 per pool); longer-tail DS hits may have different coherence properties, so generalising the 0% Null rate beyond the top of each ranking would over-claim.

#### Full per-gene catalog

The remainder of this section is the per-gene reference catalog: for each reviewed DS pool, the ranked gene entries include function, MS/NMO/neuroimmunology connection, autoimmune context, variance-shift vs mean-shift framing, verdict, and citations.

#### S17-1. Subtype DS (Progressive vs RRMS), ranked by Abs(ΔRange80)

These genes show higher patient-to-patient expression variability in progressive MS (SPMS + PPMS) than in RRMS within the quality-covariate-corrected MS-only cohort (n=507; 75 progressive vs 432 RRMS). A variance-expansion signal in progressive MS is biologically consistent with the cohort's heterogeneous disease-modifying-therapy history, broader age range, and transition from synchronous relapse-driven immune tone to smouldering, compartmentalised inflammation.

##### Rank 1–10 (Batch 1)

**CTSL (rank 1)** Lysosomal cysteine protease that processes the MHC class II invariant chain and generates antigenic peptides for class II presentation; also degrades extracellular matrix. Cathepsins as a family are elevated in MS lesions, CSF, and PBMC (Nagai et al. 1994, PMID 8164836). Cathepsins B/S/L act redundantly in EAE (Allan et al. 2015, PMID 26000904), and combined inhibition has been proposed as a therapeutic strategy. Cathepsin S dominates invariant-chain processing in peripheral APCs; cathepsin L contributes in thymus to CD4 positive selection. CTSL is a core node in HLA-DRB1*15:01-driven MS risk. CTSL expression in PBMC reflects monocyte/DC content, treatment status (corticosteroids and B-cell depletion alter APC composition), and inflammatory tone. Progressive patients are more heterogeneous than RRMS in treatment regimen, smouldering vs compartmentalised inflammation, and age-related monocytosis. These broaden the distribution without shifting the mean. **Verdict: Defensible.** Direct MHC-II antigen-processing link with a plausible compartmental-heterogeneity variance mechanism.

**NR1D1 (rank 2)** Circadian nuclear receptor (REV-ERB$\alpha$); heme-binding transcriptional repressor that antagonises ROR$\alpha$/ROR$\gamma$t at ROR response elements. REV-ERB$\alpha$ directly represses ROR$\gamma$t-driven Il17a/Il17f transcription; pharmacologic agonism (SR9009) or over-expression delays EAE onset and reduces IL-17$^+$ CD4 T cells in CNS (Chang et al. 2019, PMID 31455731; Amir et al. 2018, Cell Reports). Broadly implicated in Th17-mediated autoimmunity and circadian control of innate immunity. NR1D1 is under tight circadian oscillation with >10-fold peak/trough differences. Blood-draw time, shift-work status, and sleep disruption, all more heterogeneous in progressive patients with fatigue and disability, inflate patient-to-patient variance without changing the mean. Smouldering (rather than acute) Th17-pathway activity in progressive MS further produces stochastic REV-ERB$\alpha$ counter-regulation. **Verdict: Defensible.** Direct Th17/EAE literature combined with a clean circadian variance mechanism.

**LZTS3 (rank 3)** Postsynaptic density scaffold (a.k.a. ProSAPiP1) linking Shank3–SPAR; regulates dendritic spine maturation and NMDAR-mediated transmission (Wendholt et al. 2006). No direct PubMed hit in MS, EAE, or NMO. Protein Atlas shows brain-enriched expression (cerebral cortex, hippocampus) with low PBMC abundance. Weak. Low PBMC expression means patient-to-patient variance may reflect dropout noise inflated at the detection floor, a measurement-level rather than biology-level variance shift. A thin alternative: CNS-leakage transcripts from axonal injury in progressive MS may be more stochastic than in RRMS where blood–brain barrier disturbance is episodic. **Verdict: Speculative.** Candidate for transparent framing as "variance-discriminative but biologically obscure."

**MAD2L2 (rank 4)** Regulatory subunit of translesion DNA polymerase $\zeta$; core component of the Shieldin complex downstream of 53BP1–RIF1; inhibits 5'-end resection at double-strand breaks to promote NHEJ (Xu et al. 2015, PMID 25799992). No direct MS literature. MAD2L2/Shieldin is essential for immunoglobulin class-switch recombination in B cells (Ghezraoui et al. 2018, PMID 30022170). CSR/SHM machinery is increasingly implicated in MS B-cell pathology given the efficacy of anti-CD20. Progressive MS features clonally expanded B cells and plasmablasts (intrathecal IgG, CSF oligoclonal bands); PBMC B-cell subset composition is more heterogeneous in progressive MS across treatment classes. MAD2L2 tracks cycling B cells and germinal-centre-like activity, both with wider between-patient spread in progressive disease. **Verdict: Plausible.** B-cell-CSR pathway link; MS-specific evidence is indirect.

**UNC119B (rank 5)** Paralog of UNC119A; GDI-like solubilisation factor that binds N-myristoylated cargo (e.g. LCK, GNAT1) and shuttles them between membranes. No direct MS literature. UNC119A has a well-established T-cell role, binds and activates LCK at the immunological synapse; a V22G mutation causes idiopathic CD4 lymphopenia with impaired TCR signalling (Gorska & Alam 2012, PMID 22184408). UNC119B has overlapping but non-identical cargo specificity. If UNC119B contributes to LCK/Fyn trafficking, PBMC expression tracks T-cell composition (CD4:CD8 ratio, memory vs naive subset balance), both more heterogeneous in progressive MS due to immunosenescence and treatment history. Small-effect paralog behaviour is classically variance-inflating. **Verdict: Plausible.** T-cell link is via the paralog UNC119A; functional overlap makes UNC119B a reasonable variance reporter.

**CD9 (rank 6)** Tetraspanin cell-surface organiser; forms tetraspanin webs linking integrins ($\beta$1), CD81, Tspan-2; modulates adhesion, migration, and exosome biogenesis. CD9 is expressed on oligodendrocyte progenitor cells committed to oligodendrogenesis (Terada et al. 2002, PMID 12420314) and on premyelinating oligodendrocytes. Encephalitogenic MOG-specific T cells upregulate CD9 in EAE; anti-CD9 blockade in vitro strengthens BBB function and reduces monocyte transmigration (Reyes et al. 2018, PMC6189363). Highly expressed on marginal-zone B cells, B1 cells, plasma cells, and broadly across leukocytes. CD9 expression varies by activation state, plasma-cell/memory-B-cell composition, and monocyte/DC subset balance, all perturbed heterogeneously in progressive MS (more plasma-cell accumulation, older immune compartment, more treatment diversity). **Verdict: Defensible.** OPC, BBB, and EAE T-cell links with a compartmental variance mechanism.

**CRAT (rank 7)** Carnitine O-acetyltransferase; mitochondrial matrix bidirectional enzyme that transfers acetyl groups between acetyl-CoA and carnitine and buffers the acetyl-CoA/CoA ratio. No direct MS hit. Mitochondrial dysfunction and altered acylcarnitine profiles are reported in progressive MS serum and CSF. In RA, CD4 T cells show reduced oxidative metabolism and muscle CRAT activity correlates with T-cell respiration (Andonian et al. 2022, Sci Rep). CRAT tracks mitochondrial biogenesis, T-cell activation state, and exercise/physical-activity history, all substantially more heterogeneous in progressive MS (EDSS-driven activity differences alone span roughly 10×). Age-related mitochondrial decline further broadens the distribution. **Verdict: Plausible.** Immunometabolism pathway link with RA supporting evidence.

**NEIL1 (rank 8)** Base-excision-repair glycosylase with broad substrate range; repairs oxidative DNA lesions in nuclear and mitochondrial DNA. No direct NEIL1–MS publication. Pathway-level: oxidative DNA damage and 8-oxoG accumulation are documented in progressive MS lesions and PBMC. NEIL1/NEIL2 double knockouts develop neuroinflammation and neurodegeneration (Rolseth et al. 2019, eLife). Impaired BER creates a cGAS-STING-activating cytosolic DNA pool tied to type I IFN signatures. Oxidative burden varies sharply between patients (smoking, age, disease duration, CNS injury level). Damage-responsive transcripts induced by ROS load show classic stochastic expression, a canonical variance-inflating mechanism. **Verdict: Plausible.** Oxidative DNA repair and neuroinflammation pathway link without a direct MS genetic or clinical study.

**PEX16 (rank 9)** Integral membrane peroxin required for peroxisome membrane assembly; functions at ER during early peroxisome formation and recruits peroxisomal proteins to mature organelles. Biallelic PEX16 variants cause leukodystrophy, spastic paraplegia, cerebellar ataxia, and peripheral demyelination. AMN (the ABCD1-related peroxisomal disorder) is classically mis-diagnosed as primary progressive MS (Engelen et al. 2012, PMID 22937500). Peroxisomal $\beta$-oxidation clears VLCFAs; VLCFA accumulation causes myelin injury. Peroxisomal dysfunction is a candidate driver of progressive-MS neurodegeneration and is patient-specific. Between-patient PEX16 variance in PBMC plausibly indexes age-related mitochondrial/peroxisomal co-regulation, differential lipid metabolism, and treatment-associated metabolic effects (dimethyl fumarate engages NRF2 and peroxisomal biology). **Verdict: Plausible.** The AMN-vs-progressive-MS diagnostic overlap is a clean conceptual framing.

**POU6F1 (rank 10)** POU-domain homeobox transcription factor (a.k.a. Brn-5); binds a variant octamer motif; predominantly CNS-expressed during development but also detected in T- and B-cell lines. No PubMed hit linking POU6F1 to MS, EAE, or NMO. Originally isolated as a binder of the TCR-$\beta$ enhancer in Jurkat cells and of immunoglobulin octamer-like motifs; no follow-up functional immunology literature. Weak. Like LZTS3, POU6F1 PBMC expression is likely near detection limit, so variance shifts may reflect dropout noise as much as biology. If the TCR-$\beta$ enhancer binding is functional at low levels, between-patient TCR-repertoire differences in progressive MS could contribute, but this is a long speculative chain. **Verdict: Speculative.** Second candidate (with LZTS3) to flag as "variance-discriminative but biologically obscure"; useful contrast case showing DS detects signal where mean-shift biology is silent.

**Rank 11–30 (Batch 2)**

Genes in this tranche show modest but reproducible ΔRange80 in progressive vs RRMS and cluster into several biological families. We tag cluster membership in headings where the evidence is strong: B-cell compartment, microglia/TGF-$\beta$, and immunosenescence. The other batch-2 genes form a mixed tail dominated by compartment-drift mechanisms.

**TM2D2 (rank 11)** TM2-domain protein; TM2D family modulates $\gamma$-secretase/Notch cleavage and is implicated in APP/A$\beta$ processing in Drosophila models. No direct MS literature. TM2D3 (same family) carries a rare Alzheimer's-risk variant; Notch signalling is relevant to oligodendrocyte maturation and remyelination failure, but TM2D2 itself is not implicated. Weak. The only plausible route is age-related amyloid/neurodegeneration programs in monocytes or shared signal from neurodegeneration-associated macrophages, speculative at gene level. **Verdict: Speculative.**

**RAB3IP (rank 12)** Guanine nucleotide exchange factor (Rabin8) for RAB8A/B; drives polarised vesicle trafficking, ciliogenesis, and LPS-induced Rab8a activation on macropinosomes. No direct MS literature. RAB8A activation positively regulates TLR4 signalling and inflammatory programs in macrophages (Wall et al. 2019, Small GTPases). Plausible via monocyte activation-state heterogeneity, some progressive patients have strongly primed monocytes (smouldering inflammation), others quiescent; RAB3IP in the myeloid compartment would spread rather than shift. **Verdict: Plausible.**

**SMPD2 (rank 13)** Neutral sphingomyelinase 1 (nSMase1); hydrolyses sphingomyelin to ceramide + phosphocholine. The paralog SMPD3 (nSMase2) is a well-established modulator of remyelination; pharmacological inhibition improves myelin thickness in EAE (Chen et al. 2021, Sci Adv). SMPD2 sits in the same ceramide-stress pathway activated by TNF/IL-1. Plausible. Ceramide-generating enzymes are transcriptionally responsive to inflammatory cytokines that fluctuate unpredictably during smouldering progressive MS, producing high expression in flaring patients and basal expression in others. **Verdict: Plausible.**

**ITGB5 (rank 14) `[cluster: microglia / TGF-`$\beta$`]`** Integrin $\beta$5 subunit; pairs with $\alpha$V to form $\alpha$V$\beta$5 (vitronectin receptor); phagocytosis, TGF-$\beta$ activation, apoptotic-cell clearance. ITGB5 is a TGF-$\beta$-dependent microglial signature gene, downregulated in Tgfb1-deficient mice and part of the adult microglia molecular fingerprint (Butovsky et al. 2014, PMID 24316888). ITGB5-expressing microglia/macrophages mediate efferocytosis of myelin debris, a major lesion-remodelling step in progressive MS. Strong. Monocyte-to-microglia-like transition and efferocytic activation vary widely across progressive patients; ITGB5 expression would expand as the activated-monocyte fraction becomes heterogeneous. **Verdict: Defensible.**

**NIBAN3 (rank 15) `[cluster: B-cell compartment]`** B-cell-novel-protein 1 (BCNP1, FAM129C); B-cell-restricted plasma-membrane protein with PH, proline-rich, and leucine-zipper domains; PI3K/p38-

dependent phosphorylation. No direct MS literature; B-cell-restricted expression tracks the compartment central to progressive MS pathology (meningeal follicles, anti-CD20 benefit). Strong, classic B-cell-compartment-drift story. Progressive MS patients are highly heterogeneous in B-cell fraction due to anti-CD20 treatment status, treatment-naive exposure, and expanded memory/plasmablast fractions. **Verdict: Defensible** (as B-cell compartment proxy).

**EBF1 (rank 16)** `[cluster: B-cell compartment]` Master transcription factor for B-cell lineage commitment; required for pro-B to pre-B transition and maintenance of B-cell identity. An EBF1-intronic polymorphism (rs1368297) is associated with MS susceptibility in Spanish cohorts (Prieto et al. 2005, PMID 16255771). EBF1 forms complexes with EBV EBNA2 to drive MYC in EBV-infected B cells (Glaser et al. 2022, PMID 35858428), connecting EBF1 to the EBV–MS axis [22]. Strong. EBF1 tracks B-cell abundance; progressive-MS treatment heterogeneity and compartment drift produce variance expansion. eQTL effects further broaden the distribution. **Verdict: Defensible.**

**ST6GALNAC4 (rank 17)** $\alpha$2,6-sialyltransferase that synthesises disialyl-T antigen; modifies O-glycans on glycoproteins/glycolipids. No direct MS literature. Disialyl-T engages Siglec-7 as a glyco-immune checkpoint (Stanczak et al. 2023, PNAS); upregulated in DLBCL, consistent with a B-cell expression component. Plausible. A glycoenzyme's bulk expression depends on which sialylation-high subsets (activated B cells, plasmablasts, activated monocytes) dominate. **Verdict: Plausible.**

**TKTL1 (rank 18)** Transketolase-like 1; pentose-phosphate-pathway enzyme that drives Warburg-like aerobic glycolysis. No MS literature. Activated T cells and proliferating lymphocytes switch to aerobic glycolysis, TKTL1 could in principle mark highly activated subsets, but normally it is brain/cancer-biased with low noisy PBMC expression. Plausible-to-speculative. Low baseline + sporadic induction is a common variance-without-mean-shift pattern, but also the signature of technical/stochastic noise at the low-expression floor. **Verdict: Speculative** (with technical-artifact concern flagged).

**GATA2 (rank 19)** Zinc-finger transcription factor essential for HSC maintenance, monocyte/DC/NK differentiation, and lymphatic-vessel development. Germline GATA2 haploinsufficiency syndrome includes aggressive MS in a subset of carriers (Spinner et al. 2014, PMID 25707267). Transcriptomic meta-analyses identify GATA2 as a hub gene downregulated in MS (Hassanzadeh et al. 2024, Genomics Inform). Defensible. Progressive-MS patients have heterogeneous monocyte/DC/NK compartments (treatment and age effects); rare patients with partial GATA2 variants sit in distribution tails. **Verdict: Defensible.**

**LZTS2 (rank 20)** Leucine-zipper putative tumour suppressor 2; negative regulator of Wnt/$\beta$-catenin via nuclear exclusion; also involved in katanin-mediated microtubule severing. No MS literature. Wnt/$\beta$-catenin modulates oligodendrocyte differentiation and remyelination, but evidence is absent in the PBMC context. Weak. No obvious PBMC cell-type-marker role or activation-responsive regulation; variance may reflect technical or stochastic expression. **Verdict: Speculative.**

**ARMCX5 (rank 21)** X-linked armadillo-repeat protein; ARMCX family regulates mitochondrial trafficking in neurons via Miro/Trak2 and Parkin/PINK1. Partial ARMCX5 deletion contributes to Xq22.1 syndrome. No direct MS literature. ARMCX3 is linked to Parkinson's core proteome; mitochondrial dysfunction drives axonal damage in progressive MS. Plausible-to-speculative. If ARMCX5 is an X-inactivation escapee, female progressive MS patients (majority of cohort) could show expression tilted by X-inactivation skewing that drifts with age, a real mechanism for variance expansion in older patients, but untested. **Verdict: Speculative** (intriguing X-inactivation angle).

**TNFRSF10D (rank 22)** `[cluster: immunosenescence]` Decoy TRAIL receptor DcR2 (TRAIL-R4); binds TRAIL but has a truncated death domain, protecting from TRAIL-induced apoptosis. Canonical marker of cellular senescence. Peripheral TRAIL and its receptors are differentially expressed across RRMS/SPMS/PPMS (López-Gómez et al. 2011, PLOS ONE); DcR1/DcR2 balance modulates oligodendrocyte resistance to apoptosis in MS lesions (Cannella & Raine 2007, PMID 17610960). Strong. As a senescence marker, DcR2 varies dramatically with patient age and senescence burden; progressive MS patients are older with accelerated but heterogeneous immunosenescence. **Verdict: Defensible.**

**CXCR5 (rank 23)** `[cluster: B-cell compartment]` Chemokine receptor for CXCL13; essential for B-cell and Tfh homing to follicles and germinal centres. The CXCL13/CXCR5 axis drives meningeal ectopic lymphoid follicles in SPMS (Magliozzi et al. 2007 [21]); CSF CXCL13 is elevated in MS and correlates with B-cell numbers; S1P modulators (fingolimod, ozanimod) perturb the axis therapeutically. Strong. CXCR5 is highly expressed on naive/memory B cells and Tfh cells, compartments varying enormously across progressive patients due to DMT history, meningeal-follicle burden, and EBV reactivation. **Verdict: Defensible.**

**LINC00926 (rank 24)** `[cluster: B-cell compartment]` B-cell-restricted long non-coding RNA; marker of naive B cells in tissue; regulates WNT10B signalling. No direct MS literature. Identified as a hub lncRNA in primary Sjögren's syndrome (Feng et al. 2026, Front Immunol); expression restricted to B-cell-rich tissues. Strong, same story as NIBAN3, EBF1, CXCR5. Bulk-PBMC abundance tracks B-cell fraction almost linearly; progressive-MS treatment and compartment heterogeneity drive variance expansion. **Verdict: Defensible** (as B-cell compartment / naive-B-cell proxy).

#### S17-2. DS_MS and DS_NMO headliners, Batch 3

These genes show the largest Abs(ΔRange80) in their respective between-disease contrasts. Gene Ontology enrichment of DS_MS hits is dominated by a myeloid/inflammatory program (see S10); DS_NMO hits are dominated by a B-cell/humoral program. Where the batch-3 review noted that mean-shift methods (DM) would likely flag the same gene, we retain that annotation: it identifies genes where DS does not offer a unique lens, vs genes where cohort bimodality makes the variance-shift framing uniquely informative.

##### DS_MS (MS vs Control), myeloid program

**SDC2** Syndecan-2; transmembrane heparan sulfate proteoglycan; regulates cell-matrix interactions, cytokine/chemokine co-receptor function, and macrophage activation state. Sdc-2 amplifies microglial hypoxic release of TNF-$\alpha$, IL-1$\beta$, CCL2, CXCL12, and ROS (Hsu et al. 2008, PMID 18803305), the same innate-myeloid axis driving progressive-MS neurodegeneration (Absinta et al. 2022, PMC8830034). No direct human MS patient SDC2 expression study. SDC2 is an activation-state marker rather than a constitutive lineage gene. Expression scales with circulating monocyte activation and CD16$^{+}$ non-classical fraction, varying by MS disease phase, DMT, and subclinical infection. Variance-shift framing is favoured; no published bulk mean-shift to rebut it. **Verdict: Plausible.** Microglia/macrophage mechanism is solid; human PBMC evidence is thin versus classical myeloid markers.

**PPARG** Nuclear receptor and master driver of M2 anti-inflammatory macrophage polarisation; restrains NF-κB-driven inflammation. Central in MS-relevant macrophage biology: in EAE, macrophages shift from M1 to M2 partly under PPARG control; agonists ameliorate EAE severity (Wang et al. 2023, Front Pharmacol; Liu et al. 2025, PMC12520004). Small-molecule M2-promoting modulators are in early MS development. DMT-induced immune-phenotype shifts (dimethyl fumarate; S1P modulators), metabolic drift, and active-vs-stable disease perturb the M1/M2 equilibrium. The direction of PPARG change is patient-dependent, producing bidirectional variance without a cohort mean shift. **Verdict: Defensible.** Pathway centrality plus a bidirectional-heterogeneity variance story.

**IL1R1** Type I IL-1 receptor; signal-transducing receptor for IL-1$\alpha$/IL-1$\beta$ engaging MyD88 → NF-κB. IL-1/IL-1R1 signalling is causally required for EAE induction; endothelial IL1R1 knockout reduces EAE severity; IL-1 drives pathogenic Th17 differentiation (Pare et al. 2020, Acta Neuropathol; Lin & Edelson 2017, PMC5509030). IL1R1 surface expression on myeloid and Th subsets is highly context-dependent; cohorts sampled at mixed disease phases (relapse, stable, post-steroid) show the exact high-Range80 signature DS detects. Both mean and variance shifts plausible; DM likely also flags IL1R1. **Verdict: Defensible.**

**IL1B** Interleukin-1$\beta$; inflammasome-cleaved pro-inflammatory cytokine activated by NLRP3/caspase-1; drives Th17 polarisation, fever, BBB disruption. IL-1$\beta$ protein and IL1B transcript are detectable in CSF and active CNS lesions; published MS literature flags IL-1$\beta$ levels as non-univocal across studies ("elevated in some, normal in others, attributed to patient heterogeneity and therapy effects", Lin & Edelson 2017, PMC5509030), a near-verbatim description of a variance-shift signature. Polymorphisms associate with severity (Schrijver et al. 1999, PMID 10449236). Arguably the single strongest variance-shift candidate in the batch. Genetic variation (IL1B/IL1RN), inflammasome activation state (infection, lifestyle, microbiome), therapy (steroid suppression), and relapse phase drive large inter-patient spread. This is the textbook MS-cytokine example of documented cohort heterogeneity. **Verdict: Defensible.** Variance-shift is the better framing than mean-shift, DM gives a muddled answer because the mean is cohort-dependent. This is one of the three mechanistic anchors highlighted in the S17 synthesis above and Fig. 6.

**MMP25** Matrix metalloproteinase 25 (leukolysin, MT6-MMP); GPI-anchored, leukocyte-restricted, neutrophil-dominant protease; cleaves chemokines, $\alpha$1-proteinase inhibitor, vimentin; facilitates transendothelial migration. Catalogued among MMPs implicated in MS immunopathogenesis, BBB disruption, leukocyte extravasation, MBP cleavage generating immunogenic peptides (Yong et al., reviewed in PMC3916267). Cleaves chemokines to promote leukocyte migration. MMP25 tracks circulating neutrophil/monocyte count and activation state. Absolute neutrophil counts vary considerably across MS patients (steroid effect, S1P-modulator lymphopenia, infection); composition drift alone produces Range80 expansion in bulk PBMC. **Verdict: Defensible.** Clear pathway role; composition-driven variance is mechanistically cleaner than a mean-shift.

**IFI44** Interferon-induced protein 44; type-I IFN-stimulated gene, antiviral response, microtubule-associated. IFI44 and its paralog IFI44L are canonical members of the MS IFN-$\beta$ pharmacogenomic signature. Baseline ISG expression stratifies patients into IFN-$\beta$ responders/non-responders (van Baarsen et al. 2008, PLOS

ONE). Untreated MS patients with clinically active disease often show *reduced* ISG expression with subnormal STAT1 phosphorylation (Feng et al. 2002, PMID 12161037), so the direction of IFI44 dysregulation in MS is not monotonic. Almost a "calibration positive control" for DS. ISG expression is bimodal by design: IFN-$\beta$-treated strongly induced; untreated active subnormal; untreated stable near-control. Any mixed cohort shows dramatically wider Range80 even when the mean is similar to controls. **Verdict: Defensible.** Variance-shift is strongly favoured; DM would miss IFI44 or give ambiguous direction because treatment-induced up-regulation cancels untreated-active down-regulation. One of the three mechanistic anchors in the S17 synthesis above and Fig. 6.

**DS_NMO (NMO vs Control), B-cell / humoral program**

**MS4A1 (CD20)** B-cell surface tetraspanin of the MS4A family (chromosome 11); expressed pre-B through memory (not plasma cells); therapeutic target of rituximab, ocrelizumab, ofatumumab, ublituximab. MS4A1/CD20 is the direct therapeutic target for NMOSD. Anti-CD20 dramatically reduces relapse rate in AQP4-IgG$^+$ NMOSD (Cree et al. 2005, Neurology; Tahara et al. 2025, CNS Drugs) and is first-line in NMOSD guidelines (Herges et al. 2025, Front Immunol). Textbook case for variance-shift driven by therapy heterogeneity. A non-trivial fraction of any real NMOSD cohort is on or recently off anti-CD20, so circulating CD20$^+$ B-cell pool is near-zero; treatment-naive or recently-relapsed patients show elevated activated-B fractions. Treatment-stratified bimodality produces large Range80 inflation. NMOSD also shows expanded plasmablasts and CD11c^hi B-cell subsets that vary by disease activity. **Verdict: Defensible.** Variance-shift is the correct framing, mean direction depends on cohort composition; variance is nearly guaranteed to expand. Third mechanistic anchor in the S17 synthesis above and Fig. 6.

**CD79A** Ig-$\alpha$ component of the B-cell receptor (BCR) signalling complex; pairs with CD79B to couple surface Ig to Syk/BTK/PI3K. BCR signalling is mechanistically central to NMOSD: BTK (downstream of CD79A) is upregulated in NMOSD blood and CSF, and BTK inhibition reduces AQP4 autoantibody production in preclinical models (Wang et al. 2023, J Neuroinflammation). NMOSD BCR-repertoire sequencing reveals CNS-specific clonal populations (doi:10.1212/NXI.0000000000001034). B-cell tolerance to AQP4 is orchestrated at the B-cell level (Wilson et al. 2024, Nature, PMID 38383779). CD79A levels reflect circulating B-cell frequency and subset composition, naive, memory, class-switched, and plasmablast subsets differ; NMOSD cohorts show dysregulated B-cell differentiation (Lu et al. 2022, PMID 34991631). Anti-CD20 censors much of the CD79A$^+$ pool. **Verdict: Defensible.** Both mean and variance shifts apply; variance is especially informative given NMOSD B-cell-subset redistribution.

**PAX5** B-cell lineage commitment master TF; maintains B-cell identity by activating B-lineage and repressing alternative-lineage genes (Cobaleda et al. 2007, PMID 17440452). No direct NMOSD-specific PAX5 study. PAX5 regulates PI3K signalling via PTEN repression in B cells; is perturbed whenever B cells skew toward plasmablast/antibody-secreting fates, the very phenotype characterising NMOSD. Bulk PAX5 tracks B-cell proportion $\times$ per-cell level. NMOSD patients show wide B-cell dysregulation (reduced naive, increased switched-memory/plasmablast), variable anti-CD20 status, and heterogeneous activity. **Verdict: Plausible.** Strong B-cell-identity rationale; NMOSD-specific evidence absent; signal likely dominated by compositional rather than intrinsic PAX5 dysregulation, a lineage-marker caveat (see S17-5).

**BLK** B-lymphoid tyrosine kinase (Src family); BCR-downstream signalling; B-lineage-predominant expression. No direct NMOSD-specific publication, but BLK is among the most-replicated B-cell autoimmunity GWAS loci, SLE, RA, systemic sclerosis, Sjögren's, antiphospholipid syndrome, Kawasaki disease (Hom et al. 2008, PMID 20130895; Simpfendorfer et al. 2015, PMID 26246128). The disease haplotype lowers BLK expression, reduces BCR activation thresholds, and enhances class-switched B-cell expansion. Varies by B-cell subset composition, germline BLK haplotype (a major cis-eQTL source of inter-individual variance, PMC3412385), and treatment/activity-driven B-cell shifts. Genetic variance (cis-eQTL) is inherently variance-generating across genetically diverse cohorts, so DS may detect BLK even when DM would not. **Verdict: Plausible.** Strong genetic-variance rationale in principle; NMOSD-specific evidence absent; link rests on BLK's generic role in humoral autoimmunity.

**POU2AF1 (OCA-B)** Transcriptional coactivator docking with Oct1/Oct2; essential for germinal-centre formation and post-activation B-cell maturation; also expressed in activated T cells driving memory-T and autoimmune-effector programs. No NMOSD-specific paper. Oct1/OCA-B binding-site polymorphisms are identified in GWAS across autoimmune diseases including MS, SLE, T1D, IBD, RA (Kim et al. 2024, PNAS). OCA-B promotes pathogenic CD4 T-cell memory and demyelination in EAE. NMOSD requires T-cell help for AQP4-IgG class-switching and germinal-centre output. Low baseline in resting B cells, strongly induced on activation; germinal-centre output intensity varies substantially across patients with and without CNS attacks. **Verdict: Plausible.** Attractive mechanism; evidence is an indirect chain (activated-B + germinal-centre + autoimmunity GWAS) rather than direct NMOSD biology.

#### S17-3. PCA / LR driver genes (Batch 4), classifier-weight ranking

Having covered the Abs(ΔRange80) top-N, we turn to a complementary ranking derived from the classifier weights that actually power BIRT deployment. For each of the three DS pools, we refit the Methods-specified classifier on top-200 genes per seed (leak-fixed: quality-covariate correction fit on train fold; ΔRange80 computed on train fold), and recorded per-gene standardised weights averaged across 30 seeds (0–29). Subtype DS used direct logistic regression (C=0.1) with weight = mean Abs(LR coefficient); DS_MS used PCA(10) with weight = mean max Abs(loading across components); DS_NMO used PCA(5) similarly. Per-gene seed stability is reported below as `n_seeds_selected`.

The PCA/LR driver view and the Abs(ΔRange80) top-30 view are near-orthogonal. Out of 60 top-20 driver hits (3 pools × 20 genes), only 5 overlap with the top-30 by Abs(ΔRange80) (Subtype: BLK, PEX16; DS_MS: 0; DS_NMO: MS4A1, COL19A1, PAX5). The classifier-weight ranking rewards genes whose information content is complementary to co-varying neighbours (LR) or that align with dominant covariance axes (PCA), whereas Abs(ΔRange80) rewards raw per-gene variance inflation. In DS_MS the divergence is complete (0/20 overlap): PCA loadings surface a Type-I-IFN and B-cell axis (IL6, CMPK2, IFI44/IFI44L, TRIM34, CLEC17A, FCRL2, CD19) that is absent from the Abs(ΔRange80) top-30 (dominated by mesenchymal/matrix genes SDC2, INHBA, PLAU, PPARG, TGM2), consistent with tight ISG co-variation that a single PC absorbs.

We prioritise stable-hit drivers (≥10/30 seeds) for the verdicts below. Lower-stability entries appear in triage subsections and are given a default Speculative verdict unless there is an obvious mechanism.

**Subtype DS drivers, new from batch 4**

**CCDC22 (rank 6, n_seeds=26/30, ΔR80=+2.64)** Component of the COMMD/CCDC22/CCDC93 (CCC) complex regulating endosomal recycling of surface receptors (LDLR, copper transporters ATP7A/B, NF-κB); X-linked intellectual disability when mutated (Ritterhoff et al. 2016, PMID 26906735). CCC regulates CD36 and myeloid scavenger-receptor trafficking (Bartuzi et al. 2016, PMID 27167766). Copper dyshomeostasis is implicated in myelin lipid biogenesis (ATP7A → Menkes). Surface-receptor trafficking tracks inflammatory tone, monocyte activation, foamy-macrophage lipid loading, and steroid treatment modulate endosomal recycling heterogeneously across progressive patients. **Verdict: Plausible.**

**GPAA1 (rank 7, n_seeds=20/30, ΔR80=+2.46)** Glycosylphosphatidylinositol anchor attachment 1; transamidase subunit that attaches GPI anchors to ~150 surface proteins including CD14, CD48, CD55, CD59, Thy-1. GPI-anchored proteins are central to complement regulation (CD55/CD59). Germline GPAA1 loss-of-function causes a CDG with immune dysfunction (Nguyen et al. 2017, PMID 28918060). Complement dysregulation is a recognised progressive-MS mechanism (CD59 deficiency in CNS lesions, Ingram et al. 2014, Acta Neuropathol). GPI-anchor biosynthesis is oxidative-stress-sensitive and up-regulated in activated monocytes; progressive MS has heterogeneous myeloid activation. **Verdict: Plausible.**

**TSPAN32 (rank 12, n_seeds=14/30, ΔR80=+2.34)** Tetraspanin enriched in hematopoietic tissues (Tssc6); proposed negative regulator of T-cell activation. Tssc6 knockout mice show hyperactivated T-cell responses to TCR engagement (Tarrant et al. 2002, Blood, PMID 11781226). Tetraspanins broadly regulate integrin clustering at the immune synapse. As a T-cell attenuator, TSPAN32 tracks naive/memory T-cell balance, itself age- and treatment-heterogeneous in progressive MS. **Verdict: Plausible** (same tetraspanin-family logic as CD9 in S17-1).

**ADORA2A (rank 9, n_seeds=6/30, ΔR80=+3.28)** Adenosine A2A receptor; Gs-coupled; widely expressed on T cells, NK, DC, microglia; primary immunosuppressive adenosinergic receptor. A2AR signalling in peripheral CD4 T cells limits pathogenic Th1/Th17 differentiation; in CNS suppresses microglial TNF-$\alpha$/IL-1$\beta$ release. Higher PBMC A2AR reported in RRMS vs progressive MS (Vincenzi et al. 2013, PMID 23221060; Ingwersen et al. 2016, PMID 27316467). A2AR agonists are actively explored for MS. A2AR is transcriptionally regulated by hypoxia, inflammation (HIF-1$\alpha$ pathway), and adenosine availability, all oscillate in smouldering progressive MS. **Verdict: Defensible** on mechanism, despite lower seed stability (6/30); biological story supports keeping it in the narrative.

**BTLA (rank 18, n_seeds=3/30, ΔR80=+1.96)** B- and T-lymphocyte attenuator; inhibitory coreceptor (IgSF) on resting/memory T cells and mature B cells; binds HVEM (Watanabe et al. 2003, PMID 12796776). Soluble BTLA is elevated in MS patients and correlates with disease activity; BTLA-deficient mice show exacerbated EAE. BTLA transcription is modulated by recent T-cell activation history, naive/memory ratio, and treatment (S1P modulators, B-cell depletion). **Verdict: Defensible** on mechanism despite very low seed stability (3/30). Interpret with caution given the stability limit.

**Lower-stability Subtype DS drivers (1–7/30 seeds).** Triage verdicts only; `n_seeds_selected` ≤ 7 indicates the top-200 membership is unstable across seeds, so the mean weight is noisy. **HINFP** (1/30),

**CCDC17** (1/30), **VBP1** (3/30), **H4C6** (1/30), **C1orf159** (1/30), **THAP6** (7/30), **UROS** (1/30), **ZNF701** (2/30), **TULP3** (6/30), **GRPEL2** (3/30), **NDUFAF6** (2/30), all Speculative on current evidence. **SLC25A11** (1/30, mitochondrial oxoglutarate carrier) and **SLC16A1** (1/30, MCT1 lactate/pyruvate transporter, proposed progressive-MS drug target in Amorini et al. 2017, Mol Cell Biochem) have pathway relevance to progressive-MS metabolic-dysfunction hypotheses but are unstable here, Plausible on mechanism, Speculative on DS evidence.

**DS_MS drivers, Type-I IFN + B-cell cluster**

**IL6 (rank 1, n_seeds=16/30, ΔR80=+1.65)** Pleiotropic pro-inflammatory cytokine; master regulator of acute-phase response, Th17 differentiation, B-cell maturation. **MS context.** Elevated in CSF, serum, and PBMC of MS patients (Frei et al. 1991; Stampanoni Bassi et al. 2018, Mult Scler, PMID 29468961). Th17-driven EAE requires IL6; tocilizumab has been trialed in NMO (successful) and MS (mixed). Archetype of a variance-expanding gene, transcription heavily induced by TLR signalling, IL-1$\beta$ feedback, circadian rhythm, stress, and infection. Between-patient PBMC IL6 variance is classically high in MS. **Verdict: Defensible.** Variance and mean both shift; variance captures the bimodality (active-flare vs quiescent) better.

**CMPK2 (rank 3, n_seeds=15/30, ΔR80=+1.75)** Cytidine/uridine monophosphate kinase 2; mitochondrial ISG that generates mtDNA in macrophages, priming NLRP3 inflammasome (Zhong et al. 2018, Nature, PMID 29467146). **MS context.** Part of the type I IFN transcriptional signature elevated in MS PBMC and amplified during relapse (Hundeshagen et al. 2012, PMID 22873631). Strong. ISGs as a class show huge inter-patient variance; endogenous IFN tone is bimodal (IFN-high vs IFN-low autoimmune endotypes). Treated (IFN-$\beta$) and untreated patients further inflate variance. **Verdict: Defensible.**

**TRIM6-TRIM34 / TRIM34 (ranks 15 + 7, n_seeds=10/30 and 7/30, ΔR80≈+1.6–1.8)** TRIM-family E3 ubiquitin ligases; TRIM6 regulates IRF3 activation and type-I IFN signalling (Rajsbaum et al. 2014, Immunity, PMID 24530055); TRIM34 restricts HIV reverse transcription. The TRIM6-TRIM34 readthrough transcript appears with active IFN signalling. **MS context.** Pathway-level; overlaps with the MS IFN biomarker panel. Plausible, riding the ISG-bimodality axis of CMPK2/IFI44L. **Verdict: Plausible.**

**CLEC17A (rank 11, n_seeds=24/30, ΔR80=+1.72)** `[cluster: B-cell compartment]` C-type lectin (Prolectin); expressed on proliferating B cells in germinal-centre dark zones; binds mannose-rich glycans (Graham et al. 2009, PMID 19451649). **MS context.** Marker of activated/proliferating B cells, expanded in MS CSF (intrathecal plasmablasts, oligoclonal bands); anti-CD20 directly targets CLEC17A$^{+}$ population. Strong, bulk expression dominated by B-cell composition, bimodal on vs off anti-CD20. **Verdict: Defensible.**

**FCRL2 (rank 20, n_seeds=13/30, ΔR80=+1.53)** Fc receptor-like 2; IgG-binding inhibitory coreceptor on CD27$^{+}$IgM$^{-}$ memory B cells; modulates BCR signalling (Davis 2014, PMID 24515278). **MS context.** FCRL2$^{+}$ memory B cells are expanded in autoimmunity and tracked in MS on/off anti-CD20 diagnostic panels. Treatment-driven bimodality in memory-B-cell composition. **Verdict: Plausible.**

**IFI44L (rank 2, n_seeds=2/30)** Type-I-IFN-induced protein 44-like; antiviral ISG. **MS context.** Well-validated component of the MS PBMC IFN signature; IFN-$\beta$ treatment-response biomarker (Feng et al. 2012, Ann Neurol, PMID 22451200). Same classical ISG bimodality as CMPK2. **Verdict: Defensible** on mechanism despite low seed stability, likely collinear with IFI44 and CMPK2 and competing for PCA loadings across seeds. Interpret with caution.

**Lower-stability DS_MS drivers (1–7/30 seeds).** **CD19** (1/30), canonical B-cell marker and anti-CD19 CAR-T target in MS/NMO trials: Defensible on mechanism despite low stability (likely collinear with MS4A1/FCRL2/CLEC17A in PCA). **IL1RN** (1/30), IL-1 receptor antagonist, directly relevant to MS inflammation on the IL-1 axis with IL1B: Defensible on mechanism despite low stability. **LACC1** (1/30), laccase domain 1, myeloid polarisation, MS GWAS hit in some studies (Mellick et al. 2017): Plausible. **GZMM** (1/30), granzyme M in NK/CD8: Plausible via cytotoxic fluctuation. **AGRN** (2/30), agrin, immunological-synapse modulator: Plausible. **COL19A1** (1/30 in DS_MS, much more stable in DS_NMO, where it is in the Abs(ΔRange80) top-30). **CCDC112** (1/30), **DENND5B** (2/30), **ARSI** (2/30), **FXYD6** (7/30), **CKB** (1/30), Speculative on current evidence.

##### DS_NMO drivers, cytotoxic / innate-lymphocyte cluster

**BCAR3 (rank 13, n_seeds=30/30, ΔR80=+2.57)** `[cluster: B-cell compartment]` SH2/NSP adapter (aka NSP2, AND-34); Rap1 GEF via CAS-family scaffolding; regulates integrin-mediated adhesion and migration. Expressed on B lymphocytes and plasmablasts; regulates B-cell adhesion and trafficking. NMO is driven by antibody-secreting plasmablasts migrating across the BBB, so plasmablast-expressed scaffolding proteins are pathway-relevant. Plasmablast abundance is highly variable in NMO (flare-, treatment-dependent); BCAR3 tracks plasmablast fraction bimodally. **Verdict: Plausible.**

**KLRF1 (rank 5, n_seeds=26/30, ΔR80=+2.46)** `[cluster: cytotoxic lymphocyte]` Killer cell lectin-like receptor F1 (NKp80); activating receptor on NK cells and $\gamma\delta$ T cells; binds AICL on myeloid cells (Welte et al. 2006, PMID 16675651). NK cells are implicated in NMO, intrathecal infiltration, NK:T-cell ratio shifts. KLRF1 marks a mature CD56^dim NK subset; altered NK phenotype reported in NMOSD (Wu et al. 2016, J Neuroinflamm). Strong. NK abundance in PBMC is heterogeneous in NMO (age, treatment, flare); bimodal signal (active vs exhausted NK) fits variance inflation. **Verdict: Defensible.**

**ZBTB16 (PLZF, rank 6, n_seeds=19/30, ΔR80=+1.88)** `[cluster: cytotoxic lymphocyte]` Zinc finger and BTB domain 16; master TF for invariant NKT and innate-like $\gamma\delta$ T cells (Savage et al. 2008, Immunity, PMID 18406617); regulates IL-17A/IFN-$\gamma$ balance in innate lymphocytes. iNKT and $\gamma\delta$ T cells are altered in NMO and MS (reduced circulating iNKT in early MS). Strong. iNKT/$\gamma\delta$ populations are rare (0.01–1% of PBMC) and highly variable between individuals, classic rare-population variance. **Verdict: Defensible.**

**AREG (Amphiregulin, rank 11, n_seeds=19/30, ΔR80=+1.80)** EGFR ligand produced by Treg, ILC2, mast cells, activated T cells; promotes tissue repair and restrains inflammation via EGFR. AREG from tissue Tregs is central to post-damage repair (Arpaia et al. 2015, Cell, PMID 26580015); NMO causes severe astrocyte damage where dysregulated AREG could impair repair. Episodically induced by IL-33 and inflammatory cues; PBMC levels track recent Treg/ILC2 activation, highly patient-variable. **Verdict: Plausible.**

**GZMA (rank 12, n_seeds=17/30, ΔR80=+1.84)** `[cluster: cytotoxic lymphocyte]` Granzyme A; tryptase serine protease from CD8 T cells and NK cells; caspase-independent cell death; pro-inflammatory via gasdermin B cleavage (Zhou et al. 2020, Science, PMID 32299851). Granzyme-A+ CD8 T cells found in MS lesions (Salou et al. 2015, Ann Neurol) and NMO CSF; GZMA/gasdermin-B recently highlighted in NMO astrocyte damage. Cytotoxic-lymphocyte abundance is bimodal in NMO flare vs remission. **Verdict: Defensible.**

**SH3RF3 (rank 14, n_seeds=16/30, ΔR80=+2.02)** SH3 domain containing ring finger 3 (POSH2); scaffold in the JNK pathway; regulates stress-induced apoptosis and cell migration. Weak direct literature; JNK signalling is broadly relevant to lymphocyte activation and apoptosis. Plausible via JNK pathway tone heterogeneity. **Verdict: Plausible** (on pathway relevance), Speculative on specific MS/NMO role.

**PYCARD (ASC, rank 19, n_seeds=13/30, ΔR80=+1.81)** PYD and CARD domain containing adaptor; core of NLRP3/AIM2/NLRC4 inflammasomes; bridges sensor to caspase-1. Strong pathway relevance. Inflammasome activation is implicated in MS and NMO (IL-1$\beta$ maturation); NMO has elevated IL-1$\beta$/IL-18 in CSF. Inflammasome priming is stochastic and bimodal (primed vs naive depending on recent pathogen exposure). **Verdict: Defensible.**

**MS4A4E (rank 3, n_seeds=7/30, ΔR80=+1.74)** Membrane-spanning 4-domains A4E; MS4A cluster (same family as MS4A1/CD20 and monocyte-marker MS4A4A). MS4A locus has multiple Alzheimer's GWAS hits (MS4A6A/MS4A4E cluster, Naj et al. 2011, PMID 21460840). MS4A4A is upregulated in IFN-primed monocytes; family broadly tracks B-cell and monocyte biology. Plausible, PBMC expression varies with monocyte/B-cell composition, bimodal across NMO treatment. **Verdict: Plausible.**

**MTARC1 (rank 4, n_seeds=7/30, ΔR80=+1.65)** Mitochondrial amidoxime reducing component 1; outer-membrane molybdoenzyme; major liver disease GWAS hit (Emdin et al. 2021, J Hepatol). No direct MS/NMO literature. Weak for PBMC; could reflect metabolic heterogeneity but speculative. **Verdict: Speculative.** Borderline stability.

**Lower-stability DS_NMO drivers (1–4/30 seeds).** **CXCL3** (2/30), MIP-2$\beta$ chemokine, neutrophil chemoattractant; NMO lesions are neutrophil-rich vs MS: Defensible on mechanism despite low stability. **SIGLEC7** (1/30), inhibitory receptor on NK/monocytes, NK-subset marker similar to KLRF1: Plausible. **HBA1** (3/30), hemoglobin $\alpha$1; reticulocyte/erythroid contamination signal; NMO has hematologic abnormalities on certain treatments: Plausible as contamination-variance biomarker, not biological driver. **CYBRD1** (4/30), duodenal iron absorption, erythroid-lineage contamination marker: Speculative (same concern as HBA1). **SEMA3C** (1/30), **FAM20A** (3/30), **AKAP5** (1/30), Speculative.

#### S17-4. Summary tables

Consolidated verdict tables across the 54 reviewed genes, split by pool (Subtype DS within MS / DS_MS between MS and healthy control / DS_NMO between NMO and healthy control) and sorted by verdict (Defensible → Plausible → Speculative) within each pool.

**Table S17-4a. Subtype DS genes (Progressive vs RRMS within MS; n = 29 reviewed).**

**Table S17-4b. DS_MS genes (MS vs healthy control; n = 12 reviewed).**

**Table S17-4c. DS_NMO genes (NMO vs healthy control; n = 13 reviewed).**

#### S17-5. Caveats and interpretation limits

**Brain-enriched / low-expression speculative hits.** Three Subtype-DS hits (LZTS3, POU6F1, TKTL1) are near-detection-limit in PBMC, and their ΔRange80 signal is consistent with dropout-noise heteroscedasticity rather than biology. A near-zero-floor distribution can exhibit apparent variance expansion simply because technical dropouts become more frequent for low-count genes, and slightly-less-dropout in one subgroup widens the P10–P90 range without any true biological change. These should be verified before biological interpretation by correlating raw counts with expressed-class Range80 (i.e. recomputing after excluding zeros). We flag them transparently rather than promote them; the purpose of their inclusion here is to make the detection-first, mechanism-agnostic posture of the DS method explicit.

**Lineage markers driven by compartment composition.** Several DS_NMO hits (PAX5, BLK, POU2AF1) and Subtype-DS B-cell cluster members (NIBAN3, EBF1, CXCR5, LINC00926) are best described as reporters of B-cell (or plasmablast, or Tfh) compartment fraction rather than of intrinsic dysregulation within those compartments. In a treated/untreated mixed cohort where anti-CD20 has depleted some patients to near-zero circulating B cells, any B-cell-restricted transcript inflates variance by construction. This is real biology, the treatment stratification is clinically meaningful, but it does not mean the gene is a *pathway* driver. A DM (mean-shift baseline) module flags these genes equally well when the depletion stratum is imbalanced; the value of DS is that it captures the bimodality even when cohort means are balanced.

**Unknown-mechanism tail.** Roughly 25–30% of Subtype-DS hits reviewed (TM2D2, ARMCX5, LZTS2, and other speculative entries) have no clear immunological mechanism at the current level of literature. Rather than post-hoc rationalisation or silent exclusion, we frame these as ”surface for follow-up”, genes that DS flagged by statistical signal that merit experimental validation before biological claims are made.

**Sampling bounds.** We reviewed 54 genes out of the 200+ per-seed pool that feeds each DS module. The top-30 by Abs(ΔRange80) plus top-20 PCA/LR drivers sample the highest-weight end of the distribution; longer-tail DS hits (rank 31–200) may have different properties, potentially more speculative, potentially more cluster-coherent, and we did not exhaustively review beyond this tranche. Reported verdict distributions (e.g. fraction Defensible) should be read as properties of the top end, not of the full module.

| Gene | Rank (metric) | Verdict | One-line role | Canonical citation |
|---|---|---|---|---|
| CTSL | 1 (ΔR80) | Defensible | Lysosomal protease; MHC-II antigen processing | Allan et al. 2015 (PMID 26000904) |
| NR1D1 | 2 (ΔR80) | Defensible | Circadian REV-ERBα; represses Th17 | Chang et al. 2019 (PMID 31455731) |
| CD9 | 6 (ΔR80) | Defensible | Tetraspanin; OPC, BBB, EAE T cells | Reyes et al. 2018 (PMC6189363) |
| ITGB5 | 14 (ΔR80) | Defensible | TGF-β microglial-signature integrin | Butovsky et al. 2014 (PMID 24316888) |
| NIBAN3 | 15 (ΔR80) | Defensible | B-cell-restricted surface protein | Patel et al. 2017 (PMID 27680505) |
| EBF1 | 16 (ΔR80) | Defensible | B-cell master TF; MS GWAS; EBV-EBNA2 axis | Glaser et al. 2022 (PMID 35858428) |
| GATA2 | 19 (ΔR80) | Defensible | HSC/monocyte TF; haploinsufficiency MS link | Spinner et al. 2014 (PMID 25707267) |
| TNFRSF10D | 22 (ΔR80) | Defensible | DcR2 senescence marker; TRAIL axis in MS | López-Gómez et al. 2011 (PLOS ONE) |
| CXCR5 | 23 (ΔR80) | Defensible | CXCL13 receptor; meningeal-follicle axis | Magliozzi et al. 2007 [21] |
| LINC00926 | 24 (ΔR80) | Defensible | Naive-B-cell lncRNA | Feng et al. 2026 (Front Immunol) |
| ADORA2A | 9 (LR) | Defensible | Adenosinergic T/microglia immunosuppression | Vincenzi et al. 2013 (PMID 23221060) |
| BTLA | 18 (LR) | Defensible | Inhibitory coreceptor; MS disease activity | Watanabe et al. 2003 (PMID 12796776) |
| MAD2L2 | 4 (ΔR80) | Plausible | Shieldin; B-cell class-switch | Ghezraoui et al. 2018 (PMID 30022170) |
| UNC119B | 5 (ΔR80) | Plausible | Paralog of UNC119A (TCR-synapse LCK) | Gorska & Alam 2012 (PMID 22184408) |
| CRAT | 7 (ΔR80) | Plausible | Mitochondrial acyl-carnitine; T-cell metabolism | Andonian et al. 2022 (Sci Rep) |
| NEIL1 | 8 (ΔR80) | Plausible | Oxidative DNA glycosylase; neuroinflammation | Rolseth et al. 2019 (eLife) |
| PEX16 | 9 (ΔR80) / 20 (LR) | Plausible | Peroxisomal biogenesis; AMN/PPMS overlap | Engelen et al. 2012 (PMID 22937500) |
| RAB3IP | 12 (ΔR80) | Plausible | Rab8 GEF; TLR/Rab8 axis in monocytes | Wall et al. 2019 (Small GTPases) |
| SMPD2 | 13 (ΔR80) | Plausible | nSMase1; ceramide stress; paralog modulates myelin | Chen et al. 2021 (Sci Adv) |
| ST6GALNAC4 | 17 (ΔR80) | Plausible | Sialyltransferase; Siglec-7 ligand | Stanczak et al. 2023 (PNAS) |
| CCDC22 | 6 (LR) | Plausible | CCC endosomal-recycling complex | Bartuzi et al. 2016 (PMID 27167766) |
| GPAA1 | 7 (LR) | Plausible | GPI anchor attachment; complement proteins | Nguyen et al. 2017 (PMID 28918060) |
| TSPAN32 | 12 (LR) | Plausible | T-cell attenuating tetraspanin | Tarrant et al. 2002 (PMID 11781226) |
| LZTS3 | 3 (ΔR80) | Speculative | Postsynaptic scaffold; brain-enriched | Wendholt et al. 2006 |
| POU6F1 | 10 (ΔR80) | Speculative | CNS homeobox; old TCR-β enhancer hit | OMIM 618043 |
| TM2D2 | 11 (ΔR80) | Speculative | TM2D family; γ-secretase/Notch | Fischer et al. 2021 (bioRxiv) |
| TKTL1 | 18 (ΔR80) | Speculative | Warburg PPP enzyme; low-floor artifact risk | Coy et al. 2005 (Br J Cancer) |
| LZTS2 | 20 (ΔR80) | Speculative | Wnt/β-catenin regulator | Thyssen et al. 2006 (JBC) |
| ARMCX5 | 21 (ΔR80) | Speculative | X-linked mitochondrial trafficking | López-Doménech et al. 2012 (Nat Commun) |

| Gene | Rank (metric) | Verdict | One-line role | Canonical citation |
|---|---|---|---|---|
| PPARG | top (ΔR80) | Defensible | Nuclear receptor; M2 macrophage master | Wang et al. 2023 (Front Pharmacol) |
| IL1R1 | top (ΔR80) | Defensible | IL-1 receptor; causally required for EAE | Pare et al. 2020 (Acta Neuropathol) |
| IL1B | top (ΔR80) | Defensible | Inflammasome IL-1β; documented cohort heterogeneity | Lin & Edelson 2017 (PMC5509030) |
| MMP25 | top (ΔR80) | Defensible | Leukolysin; BBB and MBP cleavage | Yong, PMC3916267 |
| IFI44 | top (ΔR80) / 4 (PCA) | Defensible | ISG; IFN-β responder bimodality | van Baarsen et al. 2008 (PLOS ONE) |
| IL6 | 1 (PCA) | Defensible | Pleiotropic cytokine; Th17 and B maturation | Stampanoni Bassi et al. 2018 (PMID 29468961) |
| CMPK2 | 3 (PCA) | Defensible | Mitochondrial ISG; NLRP3 priming | Zhong et al. 2018 (PMID 29467146) |
| CLEC17A | 11 (PCA) | Defensible | Germinal-centre B-cell marker | Graham et al. 2009 (PMID 19451649) |
| IFI44L | 2 (PCA) | Defensible | ISG IFN-β biomarker | Feng et al. 2012 (PMID 22451200) |
| SDC2 | top (ΔR80) | Plausible | Syndecan-2; microglial inflammatory amplifier | Hsu et al. 2008 (PMID 18803305) |
| TRIM34 / TRIM6-TRIM34 | 7+15 (PCA) | Plausible | IRF3/IFN-pathway TRIM ligases | Rajsbaum et al. 2014 (PMID 24530055) |
| FCRL2 | 20 (PCA) | Plausible | Memory-B-cell inhibitory coreceptor | Davis 2014 (PMID 24515278) |

| Gene | Rank (metric) | Verdict | One-line role | Canonical citation |
|---|---|---|---|---|
| MS4A1 (CD20) | top (ΔR80) / 7 (PCA) | Defensible | Anti-CD20 therapeutic target | Cree et al. 2005 (Neurology) |
| CD79A | top (ΔR80) | Defensible | BCR Ig-α; NMOSD-relevant | Wang et al. 2023 (J Neuroinflamm) |
| KLRF1 | 5 (PCA) | Defensible | NKp80; NK-subset marker in NMO | Welte et al. 2006 (PMID 16675651) |
| ZBTB16 (PLZF) | 6 (PCA) | Defensible | iNKT/γδ master TF | Savage et al. 2008 (PMID 18406617) |
| GZMA | 12 (PCA) | Defensible | Granzyme A; gasdermin B pyroptosis | Zhou et al. 2020 (PMID 32299851) |
| PYCARD (ASC) | 19 (PCA) | Defensible | Inflammasome core adaptor | Inoue et al. 2014 |
| PAX5 | top (ΔR80) / 9 (PCA) | Plausible | B-cell-identity master TF | Cobaleda et al. 2007 (PMID 17440452) |
| BLK | top (ΔR80) | Plausible | B-cell Src kinase; autoimmune GWAS hub | Hom et al. 2008 (PMID 20130895) |
| POU2AF1 (OCA-B) | top (ΔR80) | Plausible | Oct1/2 coactivator; GC and autoimmune T memory | Kim et al. 2024 (PNAS) |
| BCAR3 | 13 (PCA) | Plausible | Plasmablast adhesion scaffold | Schrecengost et al. 2012 (Oncogene) |
| AREG | 11 (PCA) | Plausible | Treg/ILC2 EGFR-ligand; tissue repair | Arpaia et al. 2015 (PMID 26580015) |
| SH3RF3 | 14 (PCA) | Plausible | JNK-pathway scaffold | Wilhelm et al. 2007 (JBC) |
| MS4A4E | 3 (PCA) | Plausible | MS4A cluster; monocyte/B-cell biology | Naj et al. 2011 (PMID 21460840) |
| MTARC1 | 4 (PCA) | Speculative | Mitochondrial amidoxime reductase | Emdin et al. 2021 (J Hepatol) |
